# The effect of maternal pre-/early-pregnancy BMI and pregnancy smoking and alcohol on congenital heart diseases: a parental negative control study

**DOI:** 10.1101/2020.09.29.20203786

**Authors:** Kurt Taylor, Ahmed Elhakeem, Johanna Lucia Thorbjørnsrud Nader, Tiffany Yang, Elena Isaevska, Lorenzo Richiardi, Tanja Vrijkotte, Angela Pinot de Moira, Deirdre M Murray, Daragh Finn, Dan Mason, John Wright, Sam Oddie, Nel Roeleveld, Jennifer R Harris, Anne-Marie Nybo Andersen, Massimo Caputo, Deborah A. Lawlor

**Affiliations:** Population Health Science, Bristol Medical School, Bristol BS8 2BN, UK; MRC Integrative Epidemiology Unit at the University of Bristol, Bristol BS8 2PS, UK; Department of Genetics and Bioinformatics, Division of Health Data and Digitalisation, Norwegian Institute of Public Health, Oslo, Norway; Bradford Institute for Health Research, Bradford Teaching Hospitals NHS Foundation Trust, Bradford BD9 6RJ, UK; Cancer Epidemiology Unit, Department of Medical Sciences, University of Turin and CPO Piemonte, Turin, Italy; Amsterdam UMC, University of Amsterdam, Department of Public and Occupational Health, Amsterdam Public Health research institute, Amsterdam, the Netherlands; Section for Epidemiology, Department of Public Health, University of Copenhagen, Denmark; The Irish Centre for Fetal and Neonatal Translational Research (INFANT), University College Cork, Cork, Ireland; Department of Paediatrics and Child Health, University College Cork, Cork, Ireland; Centre for Reviews and Dissemination, University of York, Heslington, York, UK; Department for Health Evidence, Radboud Institute for Health Sciences, Radboud university medical center, Nijmegen 6500 HB, The Netherlands; Division of Health Data and Digitalisation, and Centre for Fertility and Health, Norwegian Institute of Public Health, Oslo, Norway; Translational Science, Bristol Medical School, Bristol BS2 8DZ, UK; Bristol NIHR Biomedical Research Center, Bristol BS1 2NT, UK

**Author notes:** Corresponding author – Oakfield House, MRC Integrative Epidemiology Unit Bristol, Population Health Science Institute, University of Bristol, BS8 2BN.

**Keywords:** congenital heart disease, risk factors, negative control

## Abstract

**Background:** Congenital heart diseases (CHDs) are the most common congenital anomaly. The causes of CHDs are largely unknown. Higher prenatal body mass index (BMI), smoking and alcohol consumption are associated with increased risk of CHDs. Whether these are causal is unclear.

**Methods and Results:** Seven European birth cohorts including 232,390 offspring (2,469 CHD cases [1.1%]) were included. We applied negative exposure paternal control analyses to explore the intrauterine effects of maternal BMI, smoking and alcohol consumption during pregnancy, on offspring CHDs and CHD severity. We used logistic regression and combined estimates using a fixed-effects meta-analysis. Analyses of BMI categories resulted in similar increased odds of CHD in overweight (mothers OR: 1.15 (1.01, 1.31) and fathers 1.10 (0.96, 1.27)) and obesity (mothers OR: 1.12 (0.93, 1.36) and fathers 1.16 (0.90, 1.50)). The association of mean BMI with CHD was null. Maternal smoking was associated with increased odds of CHD (OR: 1.11 (0.97, 1.25)) but paternal smoking was not (OR: 0.96 (0.85, 1.07)). The difference increased when removing offspring with genetic/chromosomal defects (mothers OR: 1.15 (1.01, 1.32) and fathers 0.93 (0.83, 1.05)). The positive association with maternal pregnancy smoking appeared to be driven by non-severe CHD cases (OR: 1.22 (1.04, 1.44)). Associations with maternal (OR: 1.16 (0.52, 2.58)) and paternal (OR: 1.23 (0.74, 2.06)) moderate/heavy pregnancy alcohol consumption were similar.

**Conclusions:** We found evidence of an intrauterine effect for maternal smoking on offspring CHDs, but no evidence for higher maternal BMI or alcohol consumption. Our findings provide further support for why smoking cessation is important during pregnancy.

## Introduction

Congenital heart diseases (CHDs) are the most common congenital anomaly (CA), affecting 6-8 per 1000 live births and 10% of stillbirths, and are the leading cause of death from CAs ^1^. Many CHD patients present with sequela from surgical intervention and late complications related to the anomaly, resulting in health problems that persist into adulthood ^2,3^. The causes of CHDs are largely unknown, but intrauterine mechanisms may play a role in their underlying pathophysiology ^4^. Identifying modifiable risk factors for CHDs is important for improving etiological understanding and developing preventive interventions.

Several modifiable maternal characteristics have been found to be associated with increased risk of CHDs, including maternal pre/early pregnancy body mass index (BMI) ^5–7^, smoking ^8^ and alcohol ^9^ consumption in pregnancy. Whether these are causal is unclear. A recent systematic review and meta-analysis of the association of BMI with CHDs found that risk of CHDs was higher in those whose mothers were overweight or obese at the start of pregnancy, compared with those who were normal weight. Results for underweight mothers were not reported ^5^, but a large cohort study consisting of >2,000,000 singletons found no clear association for maternal underweight status and CHDs ^6^. These results from conventional multivariable approaches may be explained by residual confounding due to incomplete identification or adjustment for confounders. Maternal active smoking ^8^ and maternal exposure to alcohol ^9^ were both associated with offspring CHDs in recent meta-analyses. However, 68% and 69% of the studies within the meta-analyses (for maternal smoking and alcohol, respectively) did not adjust for confounders. Therefore, those studies showing associations for smoking and alcohol cannot determine whether these reflect the magnitude of causal effect or are biased by confounding.

Negative control studies are widely used in laboratory science and in recent years have become increasingly used to explore causal effects in epidemiology ^10^. The idea behind negative control studies is that either the exposure or the outcome in the real experiment is substituted for a negative control exposure (or outcome) that is not a plausible risk factor but would have similar sources of bias or confounding as in the main experiment. In epidemiology this approach has been primarily used for determining the extent to which hypothesized intrauterine and early life exposures might be associated with outcomes as a result of residual confounding ^10,11^. Negative parental exposure control studies are used for this purpose. This involves comparing the confounder adjusted associations of maternal pregnancy exposures with the offspring outcome of interest to similarly adjusted associations of the same characteristics (negative controls) in the father. The assumptions of this approach are that: (i) measured and unmeasured confounders influence the exposures in the same direction and with a similar magnitude in mothers and fathers and (ii) there is no plausible reason why the exposure in the father would affect the offspring outcome (or at a minimum the paternal association would be much weaker than in the mother). In the present study we are assuming that paternal BMI, smoking and alcohol cannot causally influence offspring CHDs through intrauterine mechanisms. Under these assumptions, if there is a causal intrauterine effect of any of the maternal pregnancy exposures, we would expect to see a maternal-specific association, with no (or a much weaker) association with the equivalent paternal exposure. Similar associations in mothers and fathers would suggest that these are largely driven by residual confounding.

We aimed to explore the causal intrauterine effects of maternal pregnancy BMI, smoking and alcohol on CHDs using data from the Horizon 2020 LifeCycle project ^12^. As well as the negative parental control study providing scope to explore residual confounding, the use of a large existing collaboration of birth cohorts has considerable benefit. First, both offspring with and without CHDs are from the same underlying population and have been selected for inclusion and assessed in identical ways. Related to this most studies of risk factors for CHDs are case control studies which dominate meta-analysis results. These have advantages in that they have large numbers of CHD cases and hence greater statistical power than most cohorts, but they are prone to selection bias as response rates in controls are commonly low and some controls are selected from hospitals or clinics. Furthermore, case control studies are susceptible to information bias due to differential recall and reporting of the exposure between cases and controls. Second, we have harmonized data on all exposures, confounders and outcomes. Third, we have large numbers, with 232,390 participants in total and 2,469 CHD cases. Lastly, the ethos of the LifeCycle collaboration is that all studies contribute to each research question unless they do not have data on either exposure or outcome, meaning publication bias is minimized.

## Methods

### Inclusion criteria and participating cohorts

This study was part of the Horizon2020 LifeCycle Project. LifeCycle is a collaboration of largely European birth cohorts that aims to determine the impact of early-life stressors on risk of developing adverse cardio-vascular/-metabolic, respiratory, cognitive and mental health outcomes (http://lifecycle-project.eu) ^12^. A LifeCycle cohort was eligible for inclusion if it had information on CHD in the offspring ascertained by any method and data on at least one of the following: i) mother’s pre-/early-pregnancy BMI, ii) maternal smoking during pregnancy iii) maternal alcohol consumption during pregnancy, iv) exposures i-iii above measured in the father at a similar time to their pregnant partners. Eligible LifeCycle cohorts could be from any geographical area and with participants from any ethnic background. In total, seven cohorts were eligible and all participated: The Amsterdam Born Children and their Development Study (ABCD) ^13^, Avon Longitudinal Study of Parents and Children (ALSPAC) ^14,15^, Cork SCOPE BASELINE Study (BASELINE) ^16^, Born in Bradford (BiB) ^17^, Danish National Birth Cohort (DNBC) ^18^, Norwegian Mother, Father and Child Cohort Study (MoBa) ^19,20^ and Nascita e INFanzia: gli Effetti dell’Ambiente (NINFEA) ^21,22^. Individual cohort descriptions can be found in the Supplementary Material (Text S1). We excluded multiple births from the study population since they differ from single births for CA outcomes ^23,24^. Some previous studies have excluded infants with any known chromosomal/genetic/teratogenic defects on the assumption that modifiable risk factors are unlikely to contribute in the presence of known (genetic) causes. We have not made these exclusions in our main analyses since they are often presented as complex syndromes with variations in phenotype and severity which may be influenced by the modifiable exposures we explore here. In additional analyses we explore whether their removal alters our main results.

### BMI, smoking and alcohol measurements

We used harmonized LifeCycle data for exposure and confounder data, with the exclusion of paternal alcohol consumption which had not been harmonized by LifeCycle when we started this project. ABCD and BASELINE are additional LifeCycle cohorts (all others were core). This means that at the time of this study they were not part of the (phase 1) LifeCycle data harmonization. We harmonized the data for these cohorts to resemble the harmonized LifeCycle variables. Cohort-specific information on methods of data collection can be found in Supplementary Material (Text S2).

LifeCycle harmonized maternal BMI used measured or self-reported pre-/early-pregnancy weight and height. Pre-pregnancy weight was prioritized and if not available the earliest pregnancy measures were used. Paternal BMI was similarly reported (by the father or their pregnant partner) or measured and we prioritized the timing to be pre- or as early as possible in their partners pregnancy. BMI was used as a continuous variable for the main analyses. In cohorts that had >100 CHD cases, we also categorized BMI as underweight (BMI <18.5 kg/m^2^), normal weight (BMI 18.5 to <25 kg/m^2^), overweight (BMI 25 to <30 kg/m^2^) and obese (BMI ≥30 kg/m^2^). ALSPAC, BiB, DNBC and MoBa contributed to these analyses.

We used two LifeCycle smoking variables for maternal and paternal smoking at the time of pregnancy: (i) smoking in the first trimester (yes/no) where this was available, otherwise any smoking during pregnancy (yes/no) and (ii) categorized into non-smokers, light (< 10 cigarettes smoked per day) and heavy (≥ 10 cigarettes per day) throughout the entire pregnancy. Paternal smoking was categorised as ‘any smoking (yes/no)’ at the time of partners pregnancy.

We used two LifeCycle variables for maternal alcohol consumption: (i) binary (yes/no), which like smoking prioritized the first trimester if available but was otherwise any alcohol intake during pregnancy and (ii) categorized into non-drinkers (none), light (>0 and <3 units per week) and moderate/heavy (≥3 units per week) drinkers during pregnancy. Two studies (ALSPAC and MoBa) had data on paternal alcohol consumption in pregnancy and thus were able to harmonize variables relating to paternal alcohol for this project. We generated one variable, categorized as: non-drinkers, light (>0 and <7units per week) or moderate/heavy (≥7 units per week) drinkers (Text S3).

The rationale for prioritizing maternal pregnancy smoking and alcohol during the first trimester is because fetal cardiac development starts early in pregnancy and much of the development occurs in the first trimester ^25^. 47% and 96% of mothers had measures specifically in the first trimester for smoking and alcohol, respectively.

### Congenital heart disease outcomes

Information on CHDs was retrieved from a variety of sources depending on the cohort. ALSPAC, BiB, DNBC and NINFEA had International Classification of Diseases v10 (ICD-10) coded data. BASELINE had individual CHD diagnoses assigned by a cardiologist based on echocardiography. For ABCD and MoBa, we had a non-specific CHD diagnosis (yes/no). Data in ABCD, BASELINE, DNBC, and NINFEA were restricted to liveborn infants, whereas other cohorts included stillbirths.

In the ABCD cohort, data on CHDs in liveborn children were obtained from three different sources: (i) the infant questionnaire, which was filled out by the mother at an average infant age of 12.9 weeks, (ii) the questionnaire filled out by the mother at an average infant age of 5.1 years, and (iii) clinical data of the Youth Health Care Registration. In the ALSPAC cohort, cases were obtained from a range of data sources, including health record linkage and questionnaire data up until age 25 following European Surveillance of Congenital Anomalies (EUROCAT) guidelines ^26^. In BASELINE, at 2 months, mothers were asked of any medical problems and/or referrals. If a baby had been referred to a specialist, it was checked by a cardiologist to see if they had results from an echocardiogram with exact diagnoses reported. Further diagnoses up until age 12 were identified through records from the echocardiogram. In the BiB cohort, there were two separate sources to identify CAs. Both sources were used in this study: (i) CAs up to 5 years of age, identified in GP records by Bishop et al ^27^ following EUROCAT guidelines. ICD-10 codes were mapped to clinical term (CT)-V3 codes prior to extraction from GP records. (ii) Data extracted from the Yorkshire and Humber CAs register database. Data were ICD-10 coded. All of these were confirmed postnatally. In the DNBC, all diagnoses of congenital anomalies (according to EUROCAT guide 1.4 section 3.2 and 3.3) up until the age of 15 years were extracted from the Danish National Patient Register (DNPR) which is linked to the cohort data^28,29^. Diagnoses were ICD-coded. These data were restricted to children born alive. In MoBa, information on whether a child had a CHD or not was obtained though linkage to the Medical Birth Registry of Norway (MBRN). All maternity units in Norway must notify births to the MBRN. In the NINFEA cohort, CHDs were reported in the second questionnaire compiled 6 months after birth. Mothers compiled a checklist that included pre-specified anomalies. If the child died or had any surgery performed in the first 6 months, the cause of death and type of surgery were also checked to see if any CA was reported. Data were coded using ICD-10 codes by an experienced pediatrician and were reassessed by an independent MD. Further details of the sources of data for CHDs in each cohort are provided in the Supplementary Material (Text S4).

In all studies, our main outcome was any CHD. Where data allowed (e.g. when we had full ICD-codes), any CHD was defined according to EUROCAT, which excludes isolated patent ductus arteriosus (PDA) and peripheral pulmonary artery stenosis in preterm births (gestational age <37 weeks) (Table S2). We also categorized cases into severe CHD (heterotaxia, conotruncal defect, atrioventricular septal defect, anomalous pulmonary venous return, left ventricle outflow tract obstruction, right ventricle outflow tract obstruction, other complex defects) and the remainder as non-severe CHD (PDA [in full term infants], valvular pulmonary stenosis, ventricular septal defect [VSD], atrial septum defects [ASD], unspecified septal defects, isolated valve defects, other specified heart defects, unspecified heart defects) ^30,31^ (Table S2).

### Confounders

Analyses were adjusted for a number of confounders based on their known or plausible influence on one or more of the maternal pregnancy exposures and on CHD: Maternal age (all exposures), parity (all exposures), ethnicity (all exposures), socioeconomic position (SEP; all exposures), smoking (for BMI and alcohol analyses), alcohol use (for BMI and smoking analyses). In the paternal negative control analyses confounders were similar: fathers’ age (all exposures), number of children (all exposures), ethnicity (all exposures), SEP (all exposures) smoking (for BMI and alcohol), alcohol use (for BMI and smoking). We also adjusted for offspring sex in all adjusted analyses. We used educational attainment for both parents’ measures of SEP. Full details of our selection and harmonization of confounders is provided in the Supplementary Material (Text S5).

### Statistical analysis

Analyses were conducted in either R (version 3.6.1) or Stata (version 16). An analysis plan was written and published in October 2019, with any subsequent changes and their rationale documented in the publication ^32^. All associations between exposures and CHDs were performed within participating studies using logistic regression (binary for main analyses and multinomial for CHD severity analyses). In the two largest cohorts (DNBC and MoBa), we assessed deviation from linearity in our models in the BMI analyses by running our main confounder adjusted model with BMI split into fifths. We ran regression models with these fifths as four indicator variables (non-linear) and compared this model with one in which the fifths were treated as a continuous (score) variable. We used a likelihood ratio comparison to compare these two models. All analyses were run unadjusted and adjusted for maternal/paternal age, SEP, parity (maternal) or number of children (paternal), ethnicity, smoking and/or alcohol (depending on exposure) and offspring sex. In the adjusted models, studies were asked to adjust for as many of the confounders as possible. All analyses were performed with maximal numbers (i.e. numbers included in each model will vary due to missing data on exposure/outcome or confounders). In a sensitivity analysis, we repeated our main analyses using complete-case data to assess whether missing data were influencing the results.

For the main negative control analyses – i.e. where we directly compared maternal to paternal exposure-CHD associations – we used multivariable logistic regression in which both maternal and paternal exposures were adjusted for the other parent’s exposure. This produces a maternal association that adjusts for maternal confounders as well as the paternal exposure, and similarly a paternal association adjusting for paternal confounders and the maternal exposure. The rationale for mutually adjusting for the other parent’s exposure is that parental BMI, smoking and alcohol may relate to each other through assortative mating and/or convergence of behaviours that occurs overtime in couples ^33^. Causal structural graphs together with simulated data show failure to undertake this mutual adjustment will bias the negative control analysis results ^34^. Also, paternal exposures may have some intrauterine impact, for example via passive smoking or paternal support for the mother to reduce alcohol and have a normal BMI during her pre-conceptual period or in pregnancy ^35^. Mutual adjustment for maternal and paternal confounders was necessary for ensuring both parental results were fully adjusted. Comparisons between maternal and paternal associations from this model were assessed by visually comparing the two results. In addition, statistical evidence of any differences was obtained by calculating differences in log odds of CHD between the fathers’ and mothers’ associations and report the corresponding P-value (P_diff_), under the null hypothesis that there is no difference between the maternal and paternal estimate.

Analyses were conducted separately in each study and then meta-analysed using the *meta* package in R ^36^. All the data used in the present study originated from European birth cohorts, with broadly similar methods and therefore, we assumed that they were each estimating an association from the same underlying populations and used a fixed-effects meta-analysis. To explore this assumption, differences between studies were assessed using I^2^ and Cochrane Q P-values for heterogeneity ^37^.

### Additional analyses

We repeated the main analyses after excluding infants with any known chromosomal/genetic/teratogenic defects. Methods of data collection and definition of these variables can be found in Supplementary Material (Table S3). Folic acid supplementation has been shown to lower risk of birth defects and adverse pregnancy outcomes ^38,39^. We repeated the adjusted maternal analyses with additional adjustment for first trimester folic acid supplementation (yes/no).

## Results

### Participant characteristics

Figures S1-S7 in the Supplementary Material show flowcharts designating the assignment of participants into analysis groups for each cohort. In total, 7 cohorts including 232,390 offspring with 2,469 CHD cases (1.1%) were included. The prevalence of CHD was close to 1% in most cohorts, with the lowest being in ABCD (0.4%) and the highest in DNBC (1.4%) (**Table 1**). **Table 1** shows the distributions of maternal and paternal characteristics for each cohort. Mean maternal age across the cohorts was broadly similar (all late 20s to early 30s). Mean BMI was also similar across the cohorts but proportions in different categories varied, with the lowest prevalence of pre-/early-pregnancy obesity seen in NINFEA (5%) and the highest in BiB (21%). There was also variation in maternal smoking and alcohol consumption across the cohorts, with notably high levels of both smoking (25% and 26%, respectively) and alcohol (55% and 45%, respectively) in ALSPAC and DNBC. Fathers were generally older than mothers and more likely to smoke and drink alcohol, with the overall patterns of between study differences being similar to those for the mothers. There were differing levels of missing data in each cohort (summarized in Table S4 and also illustrated in cohort specific flow charts (Figures S1-S7). To check whether missing data influenced any of our results, we report complete-case analysis results for our main analyses in the Supplementary Material. Overall, complete-case results from meta-analyses were comparable (Tables S5-S8).

**Table 1.**
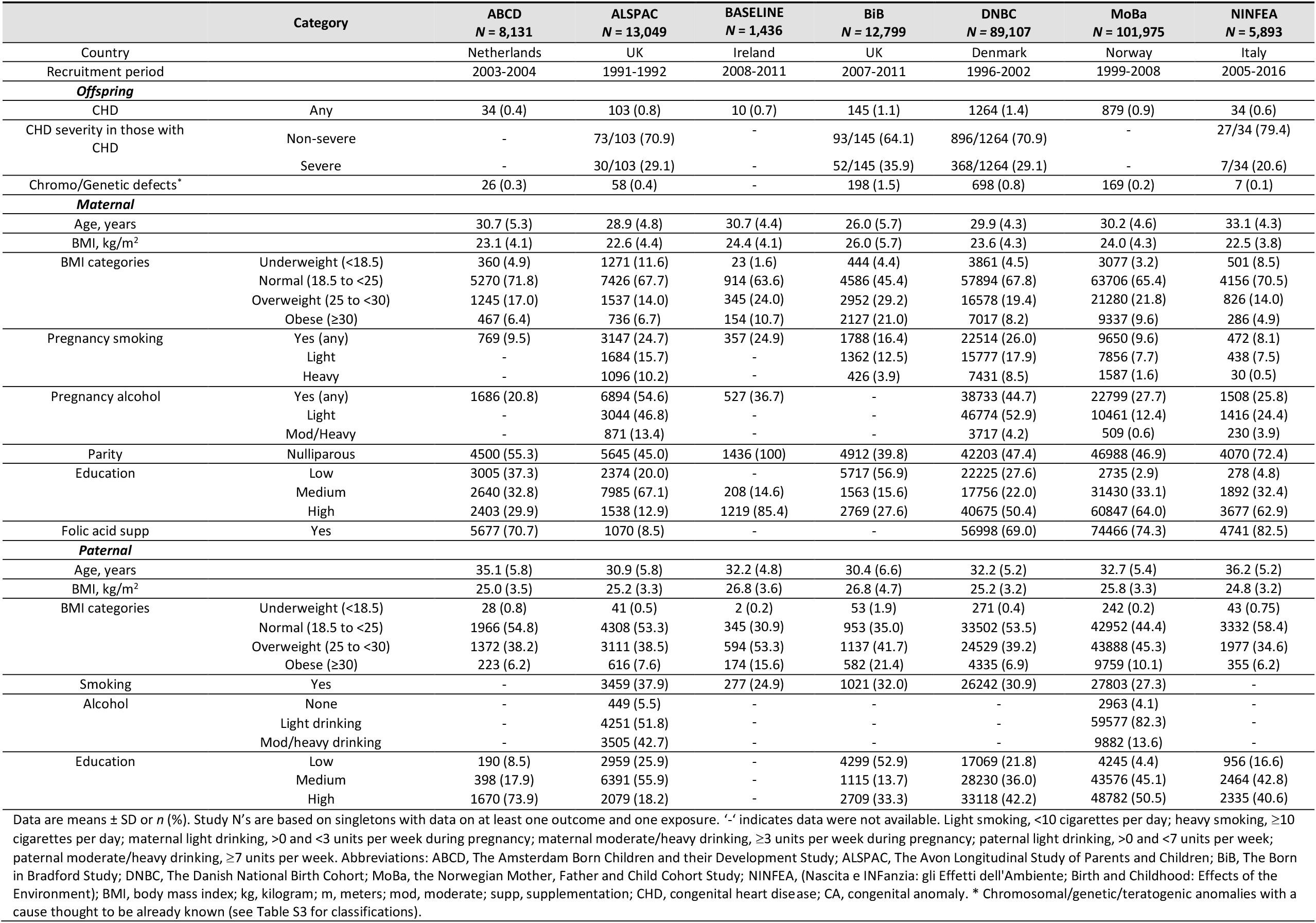
Characteristics of the participating cohorts.

### BMI and CHDs

In confounder and other parent BMI adjusted analyses, there was no difference in the odds of offspring CHD per 1kg/m^2^ difference in maternal BMI (OR: 1.00, 95%CI: 0.99, 1.02) or paternal mean BMI (OR: 1.01, 95%CI: 0.99, 1.03) (P_diff_ = 0.43), with both being close to the null (**Figure 1A**). Results were similar across studies (I^2^ = 0% & 0%, P_heterogeneity_ = 0.45 & 0.68 for maternal and paternal BMI respectively). Unadjusted and confounder only adjusted results did not differ notably from those presented in **Figure 1** (Figure S8). Figures S9 and S10 show the odds ratios of CHD by fifths of the BMI distribution for mothers and fathers in DNBC and MoBa, respectively. Whilst there was statistical evidence for a linear trend in DNBC mothers (p-value for per fifth increase = 0.05) the graph shows this was driven by increased risk only in the highest fifth, with the 2nd, 3rd and 4th fifth (compared to the first) consistent with the null. In MoBa mothers there was no clear pattern with some evidence that the 4th compared to the 1st fifth was associated with lower risk with both other categories being consistent with the null (p-value for linear trend in MoBa = 0.22). Whilst the p-values for the likelihood ratio comparing the linear model with the category model (0.03 and 0.09, for DNBC and MoBa mothers, respectfully) provide statistical support for the category model in each, this is based on just one of the fifths. Results for the fathers are broadly consistent with those for the mothers, and overall these results are consistent with no association of maternal or paternal mean BMI with offspring CHD risk.

**Figure 1.**
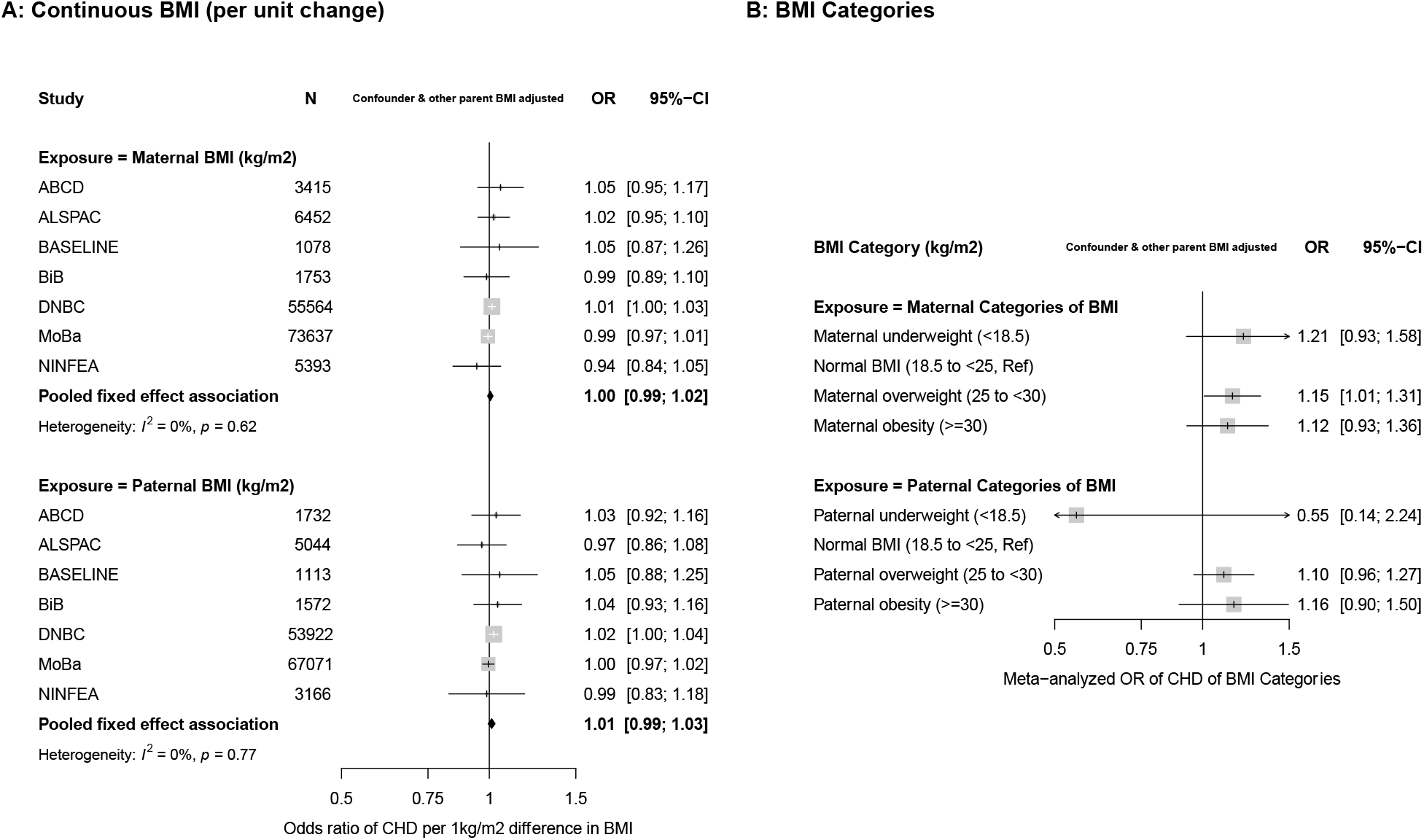
Associations between maternal and paternal pre/early pregnancy body mass index (BMI) and offspring congenital heart disease (CHD). Figure 1A shows odds ratios of CHD for a one-unit (1kg/m^2^) difference in maternal BMI (top graph) and paternal BMI (bottom graph) in each study and pooled across studies. Figure 1B shows the pooled (across ALSPAC, BiB, DNBC, MoBa) results for maternal (top) and paternal (bottom) BMI categories. Results are odds ratios of CHD in comparison to normal BMI. The study specific results for BMI categories are shown in supplementary material Figures S11-S13. 147,292 mothers (1,430 with an offspring with CHD) and 133,620 fathers (1,325 with an offspring with CHD) were included in the analyses presented in this figure. All results are adjusted for confounders (depending on cohort: maternal and paternal age, education, ethnicity, smoking, alcohol, maternal parity and offspring sex) as well as the other parents BMI.

In analyses of BMI categories, there were increased odds of offspring CHD in overweight and obese mothers and fathers compared with those of a normal BMI, with similar magnitudes of association in both parents (P_diff_ overweight = 0.65 & P_diff_ obese = 0.83) (**Figure 1B**). Underweight mothers had an increased odds of offspring CHD, whereas underweight fathers had a decreased odds of offspring CHD. Because of very small numbers of underweight parents, particularly fathers, however, results were imprecise with wide confidence intervals and there was no statistical evidence for between parental differences for underweight (P_diff_ underweight = 0.27). Individual study results for BMI categories are shown in Figures S11-S13; there was no statistical evidence of heterogeneity across studies in the results shown in Figure 1B (I^2^ = 0%, Figure S13).

Analyses of continuously measured BMI with CHD cases separated into severe and non-severe showed similar null associations for both mothers and fathers (P_diff_ = 1.00 for severe and 0.53 for non-severe) (Figure S14) as well as after adjustment for folic acid supplementation in maternal analyses (Figure S15). Results with both continuous and categories of BMI were unchanged when offspring with chromosomal/genetic defects were removed from the study population (Figure S16 and Table S9).

### Smoking and CHDs

In confounder and other parental smoking adjusted analyses any maternal smoking in pregnancy was associated with increased odds of CHD (OR: 1.11, 95%CI: 0.97, 1.25), whereas paternal smoking at the time of their partners pregnancy did not increase odds of offspring CHD (OR: 0.96, 95%CI: 0.85, 1.07) (P_diff_ = 0.09) (**Figure 2A**). There was no statistical evidence of heterogeneity across studies for maternal or paternal estimates (I^2^ = 0% & 0%, P_heterogeneity_ = 0.82 & 0.65 for maternal and paternal smoking, respectively). Results for unadjusted analyses were consistent with the confounder and mutual parent smoking adjusted result, whereas confounder only analyses were slightly attenuated for maternal smoking (Figure S17). When removing offspring with a chromosomal/genetic defect, the magnitude of the association for maternal smoking and CHDs increased slightly (OR: 1.15, 95%CI: 1.01, 1.32), and that for paternal smoking decreased slightly (OR: 0.93, 95%CI: 0.83, 1.05), (P_diff_ = 0.02) (**Figure 2B** & Figure S18). Adjusting for folic acid supplementation did not change results from main analyses (Figure S19).

**Figure 2.**
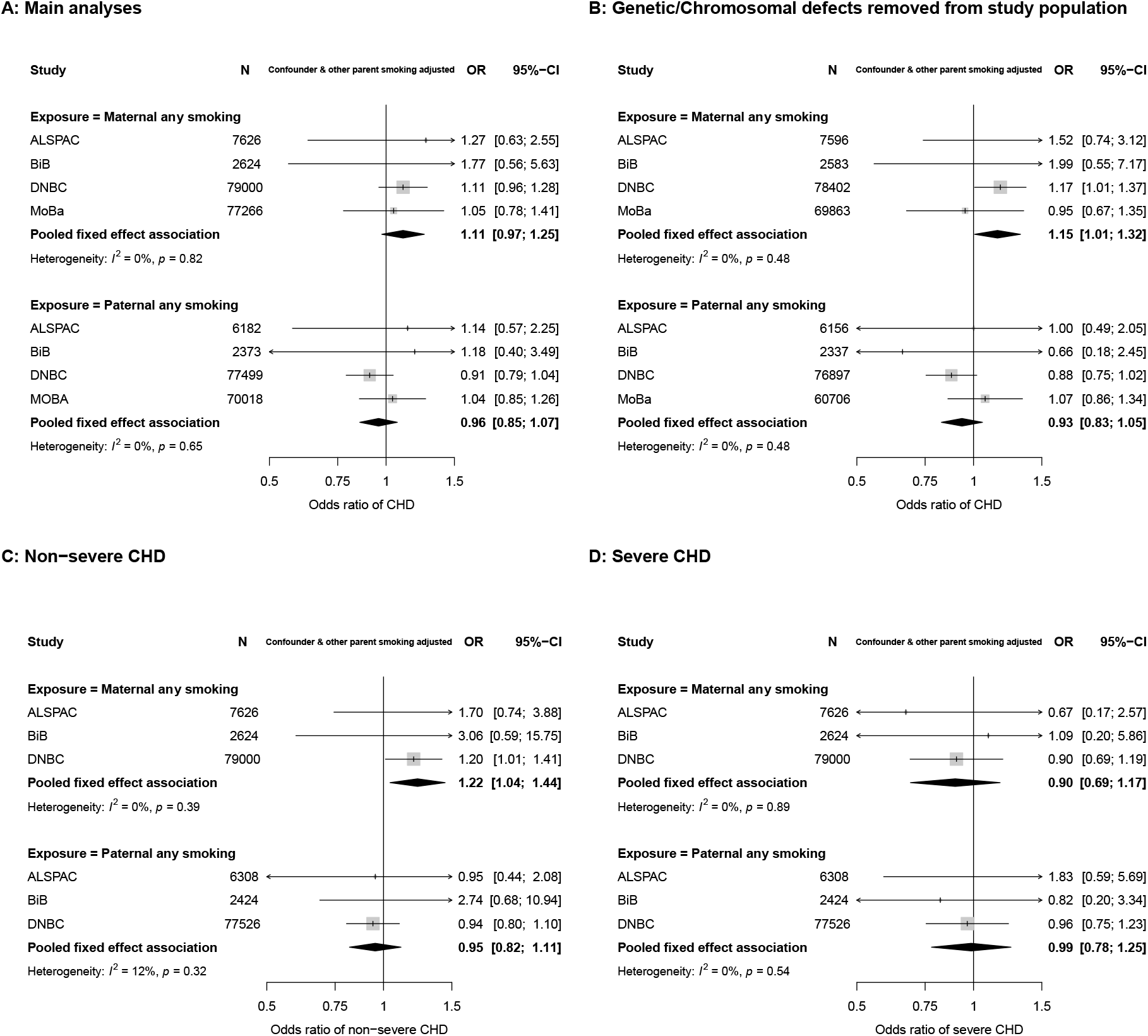
Associations in each study and pooled across studies for maternal and paternal pregnancy smoking and offspring congenital heart disease (CHD). Maternal first trimester smoking was prioritised and used where possible. Figure 2A shows odds ratios of any CHD for any maternal smoking during pregnancy (top graph) and paternal smoking (bottom graph). Figure 2B shows odds ratios of any CHD after removing those with a chromosomal/genetic defect from the study population. 166,516 & 158,444 mothers (1,802 & 1,527 with an offspring with CHD) and 156,072 & 146,096 fathers (1,734 & 1,449 with an offspring with CHD) were included in 2A and 2B respectively. Figures 2C and 2D show odds ratios of non-severe CHD and severe CHD respectively. 89,250 mothers (828 non-severe CHD & 347 severe CHD) and 86,258 fathers (813 non-severe CHD & 333 severe CHD) were included in the CHD severity analyses shown (2C & 2D). All results are adjusted for confounders (depending on cohort: maternal and paternal age, education, ethnicity, alcohol, maternal parity and offspring sex) as well as the other parents smoking.

The positive association between maternal smoking and offspring CHD appeared to be driven by an association with non-severe CHD (OR: 1.22, 95%CI: 1.04, 1.44) with no increased risk of severe CHD (OR: 0.95, 95%CI: 0.82, 1.11) (**Figures 2C & 2D**). When we analysed maternal smoking frequency categories (i.e. none, light and heavy smoking), the results did not support an effect of heaviness over and above what we saw with any (first trimester) smoking (Figure S20). Related to this, the maternal and paternal associations for these categories were statistically consistent (P_diff_ = 0.25 & 0.38 for light and heavy smoking, respectively) (Figure S20).

### Alcohol and CHDs

Due to lack of relevant paternal data, we were unable to undertake negative control analyses for any first trimester alcohol consumption. Maternal only associations for that exposure are presented here followed by the negative control analyses for levels of alcohol intake at any time in pregnancy. With adjustment for all confounders, any maternal first trimester alcohol consumption was not associated with odds of offspring CHD in meta-analyses from 5 cohorts (OR: 1.03, 95%CI: 0.94, 1.13). Results were unchanged in unadjusted models (Figure S21) and with additional adjustment for folic acid supplementation (Figure S22). There was a small increase in risk when restricting these analyses to non-severe CHD (OR: 1.07, 95%CI: 0.93, 1.22) although confidence intervals included the null. Associations for severe CHD were null (OR: 0.91, 95%CI: 0.73, 1.12) (Figure S23).

In confounder and other parental alcohol adjusted analyses, there was weak evidence of an association between maternal light alcohol intake and CHDs (OR: 1.15, 95%CI: 0.90, 1.48), although this did not statistically differ from paternal light intake (OR: 1.01, 95%CI: 0.63, 1.62) (P_diff_ = 0.63). Associations for moderate/heavy intake were consistent for maternal and paternal alcohol use (P_diff_ = 0.75) with point estimates showing weak positive associations, but with wide confidence intervals that included the null (**Figure 3A and 3B**). We did not test associations between levels of alcohol intake and CHD severity due to small numbers. Results for alcohol analyses were materially unchanged when removing offspring with a chromosomal/genetic defect from the study population (Table S10). Due to the small number of cohorts having paternal alcohol data, we also show confounder adjusted models (without mutual paternal adjustment) for maternal alcohol intake (**Figure 3C**). The point estimate for maternal light drinking was very close to the null and that for heavy drinking suggested it resulted in increased risk of offspring CHD. However, both of these estimates had wide confidence intervals due to relatively few women reporting drinking (particularly heavily) during pregnancy. Results in unadjusted analyses were unchanged (Figure S24).

**Figure 3.**
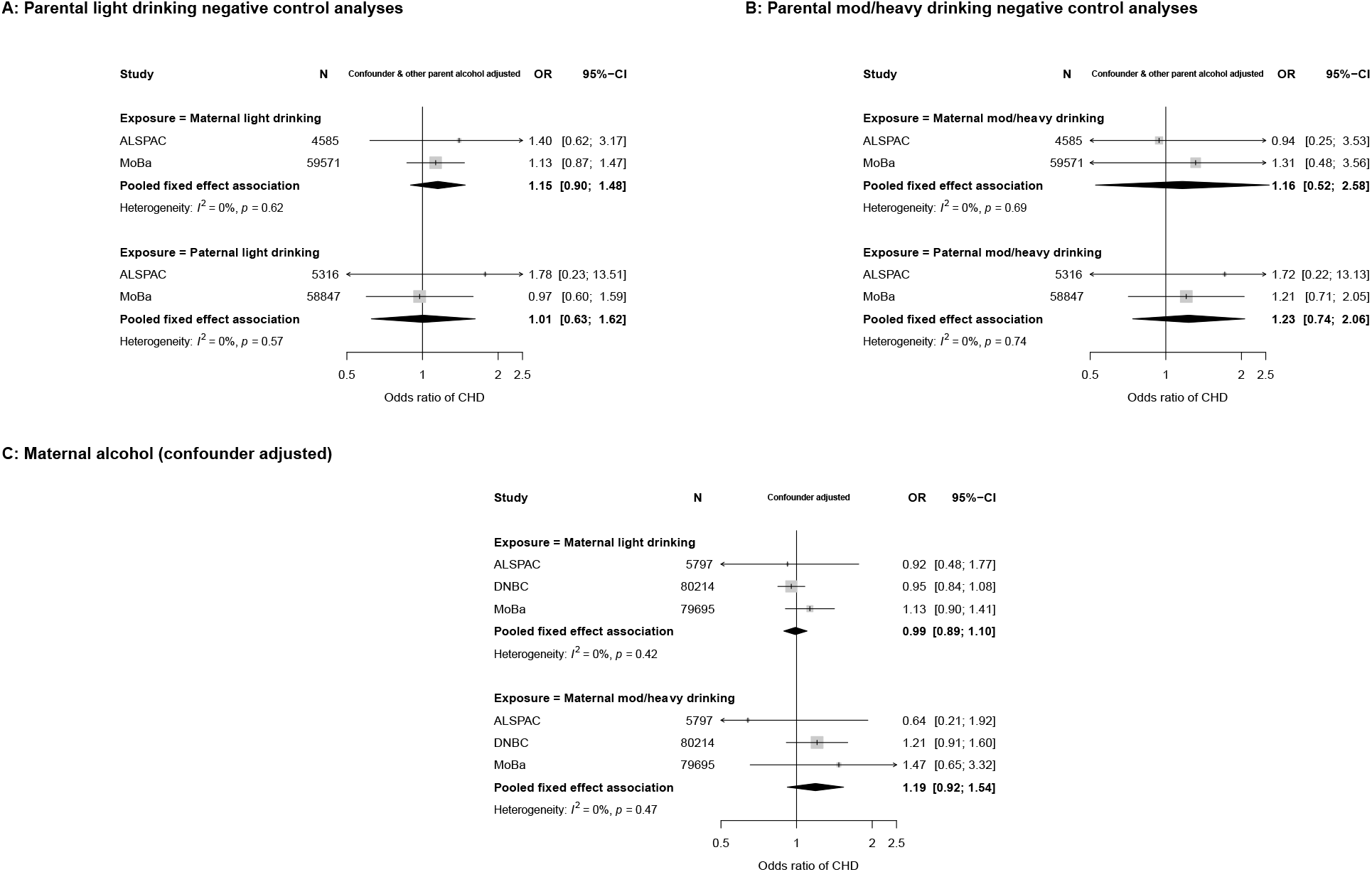
Associations in each study and pooled across studies for maternal and paternal pregnancy alcohol intake and offspring CHDs. Figure 3A shows confounder and other parent’s alcohol adjusted odds ratios of any CHD for maternal light drinking during pregnancy (top graph) and paternal light drinking (bottom graph). Figure 3B shows confounder and other parent’s alcohol adjusted odds ratios of any CHD for maternal moderate/heavy drinking during pregnancy (top graph) and paternal moderate/heavy drinking (bottom graph). 64,156 mothers with 524 CHD cases and 64,163 fathers with 529 CHD cases were included in the alcohol negative control analyses shown (3A & 3B). Figure 3C shows confounder adjusted odds ratios of any CHD for maternal light drinking during pregnancy (top graph) and maternal mod/heavy drinking (bottom graph) (165,706 mothers with 1,823 CHD cases). Confounders (depending on cohort): maternal and paternal age, education, ethnicity, smoking, maternal parity, offspring sex (and other parental alcohol intake in panels A & B).

## Discussion

In this large multi-cohort study, we found evidence that maternal pregnancy smoking increased offspring CHD risk via intrauterine mechanisms and that this appeared to be driven by a specific effect on non-severe CHDs. We did not find robust evidence to suggest a causal intrauterine effect of higher maternal pre-/early-pregnancy mean BMI or overweight or obesity on offspring CHD risk. Nor did we find evidence of an intrauterine effect of alcohol consumption on offspring CHD risk, although we acknowledge that for alcohol, we had less data and limited statistical power. To our knowledge, this is the first study to use a parental negative control method to explore whether maternal exposures have a causal intrauterine effect on offspring CHDs or whether associations are explained by residual confounding, which would generate a similar association for parental exposures.

We found increased odds of offspring CHD in mothers who were overweight and obese. This is consistent with the most recent systematic review and meta-analysis, which included 2,416,546 participants (57,172 with offspring CHD), from 19 studies and reported increased risk of any offspring CHD in women who were overweight or obese during pregnancy ^5^. However, adjustment for confounders was poor, with 10 of the 19 included studies not providing information on confounder adjustment or not adjusting for any confounders. With more stringent confounder adjustment and the findings from a negative control study, our results suggest that the increased risk of offspring CHD in overweight and obese mothers is largely the result of residual confounding. We also found that mothers who were underweight at the start of pregnancy were at increased risk of having offspring with CHD, whereas underweight in fathers appeared to be protective of offspring CHD. There were 9,537 underweight mothers (4.4%) but only 680 underweight fathers (0.4%), making the fathers analyses imprecise and our negative control analyses lacking in power to reliably identify parental differences. The recent systematic review mentioned above did not report on associations of underweight with CHDs because too few studies looked at this. A large Swedish linkage study of over 2 million singleton live born infants (born between 1992 to 2012 with 28,628 CHD cases), has explored associations with maternal underweight, as well as overweight and three grades of obesity ^7^. It is difficult to compare the results from that study with ours as they only present associations of maternal BMI with specific subtypes of CHDs, and not with any CHD as in our main analyses. Risks of offspring CHD were similar in underweight compared to normal weight women for all types of CHD except for mitral to tricuspid valve defects (14 cases), pulmonary valve defects (24 cases) and right ventricular defects (5 cases), where there was some evidence of increased prevalence with underweight. However, these estimates were imprecise, with confidence intervals including the null. Whilst our findings suggest maternal underweight might increase offspring risk of any CHD, we lacked power to rule out residual confounding in our negative control analyses and published studies have limited power to explore any effects in women. The global obesity epidemic, which is reflected in contemporary obstetric populations, might limit any potential concerns about maternal underweight. However, as the prevalence of CHD in some low- and middle-income countries is high ^40^, and these countries currently experience the double burden of under- and over-nutrition we would argue further exploration of any possible impact of maternal underweight is warranted.

Consistent with our findings, a recent meta-analysis of >8 million participants (137,575 CHD cases) from 125 studies reported positive associations between maternal pregnancy smoking and offspring CHDs ^8^. There was substantial heterogeneity (I^2^ = 89%) in their pooled results and only 68% of the included studies report adjustment for confounders. The authors also report positive associations between maternal passive smoking and paternal active smoking with offspring CHDs, both of which (somewhat unexpectedly) had stronger magnitudes of association than results from maternal active smoking. Our results, including the negative control study, add to the previous research findings by providing more robust evidence that these associations are unlikely to be explained by residual confounding and are potentially causal. Other research has shown that pregnancy smoking is a risk factor for orofacial clefts ^41^. The prevalence of CHD is around 1% in the general population, as shown in our study, yet in those with orofacial clefts, CHD prevalence rates of up to 20% have been reported ^42^. Both the heart and the palate develop during early pregnancy around weeks 5 to 9. Therefore, it is plausible that smoking in early pregnancy could disturb common biological pathways in these conditions. We found that the associations for maternal smoking were largely driven by an effect in non-severe CHDs, with the association strengthening when those with chromosomal or genetic defects were removed. Previous research has reported positive associations between maternal smoking and septal defects, in particular for ASDs ^43–45^ which are defined as non-severe according to the classification system used in our study.

In confounder adjusted analyses maternal alcohol consumption in the first trimester of pregnancy was not associated with offspring CHD. There was some evidence that maternal moderate or heavy alcohol consumption any time in pregnancy was associated with increased risk of offspring CHD. Whilst associations between mothers and fathers light, moderate and heavy alcohol consumption, compared with none, were statistically consistent, only 2 cohorts (80,627 participants, 703 with offspring CHD) had alcohol information on fathers around the time of their partners pregnancy. Associations for fathers in particular were imprecise with wide confidence intervals. Two recent meta-analyses found consistent modest increases in risk of offspring CHD in mothers reporting alcohol consumption in pregnancy (OR: 1.11 (95%CI: 0.96, 1.29) ^46^ and 1.16 (1.05, 1.27) ^9^. Although the first of these concluded ‘no association’ it can be seen that the results for the two are consistent, and the larger sample size of the second has increased precision. Of note, the second of these studies also explored paternal consumption and found increased risk of offspring CHD related to fathers’ alcohol consumption (1.44 (1.19, 1.74)) ^9^. Although the odds ratio for fathers’ consumption suggests a stronger effect, the confidence intervals are wide, and the result is statistically consistent with that for mothers’ alcohol. As in our study there were fewer studies with data on parental alcohol consumption around the time of their partners pregnancy. Taken together with our findings these suggest that positive associations of maternal alcohol consumption with offspring CHD may be due to residual confounding rather than a causal intrauterine effect.

The key strengths of this study are its large sample size, the use of a negative paternal exposures control study and the pooling of results from several cohort studies that are less prone to selection bias that can occur in case control studies and are not selected based on publication, but on being part of an existing collaboration. The latter reduces the risk of publication bias as studies were included if they had data and not on the basis of (published) results. This also allowed us to explore replication across studies and the consistency of findings between studies in our main analyses adds confidence to our conclusions.

The use of harmonized data from LifeCycle is a strength that limits between study heterogeneity. However, harmonizing data across several studies, as we have done in LifeCycle, can mean that some variables lose detail. Here that is particularly relevant for the exposure and confounding variables. For example, we were not able to explore pack weeks of smoking across the entire pregnancy. Simplified confounder measurements, such as Western versus non-Western for ethnicity could result in residual confounding if more specific ethnic groups have strong influences on exposure and outcome. Furthermore, there were other confounders that we considered, including type-1 / existing diabetes and physical activity, but had too few numbers (diabetes) across all cohorts or too few studies with data (physical activity) to include. However, we aimed to address any form or residual confounding in our paternal negative control analyses. Under the assumption that adjusted for but poorly measured (e.g. ethnicity) or unadjusted for (e.g. physical activity) influence paternal exposure in the same direction and to the same extent as in mothers, observing parental consistency of association this implies the maternal association is influenced by residual confounding.

We were not able to fully harmonize outcome data with the key differences between studies being the extent to which they only included cases that were diagnosed antenatally or at birth or whether they included cases later in life. MoBa (N = 101,975 participants and N = 879 cases) only had cases diagnosed antenatally or around the time of birth, with the remaining cohorts having diagnoses beyond antenatal care, ranging from 6 months to 25 years. Many previous studies have only included cases diagnosed at birth or early infancy. They, and the cohorts included here that only have these early life cases, may be biased by outcome misclassification (i.e. the offspring who would have been diagnosed later in life are treated as not having CHD). This is an important point for consideration because although most CHDs are identified in utero or at birth, many are diagnosed after discharge from hospital during childhood or even adulthood ^47^. Therefore, It is reassuring therefore that our main results are largely consistent across studies. In confounder and other parent adjusted smoking analyses, the weakest association was found in MoBa (**Figure 2A**). It is likely that we missed some non-severe cases in MoBa which were diagnosed later in life. Given that we demonstrate the smoking results were largely driven by non-severe CHDs, this could have biased MoBa (and therefore meta-analysis) results towards the null.

The negative control analyses assume that factors that would confound the maternal exposure-offspring CHD associations would have a similar magnitude and direction of confounding for the equivalent paternal associations, irrespective of whether the confounders are measured or if measured how accurately and precisely they are measured. This is likely to be true for paternal negative control exposure studies, as used here ^10,11^. It also assumes that there is no plausible intrauterine mechanism through which the paternal exposure could impact offspring CHD. Whilst it has been argued that paternal epigenetic preconception effects ^48^, and in the case of smoking, passive smoking could have an impact, we would expect any such paternal exposure effect to be weaker than the maternal exposure effect. Heart development occurs in utero (specifically in early pregnancy) and passive smoking would expose the fetus to a much lower dose than active maternal smoking. As proof of principle, this approach confirms the strong effect of smoking on birth weight, and fetal growth assessed by repeat ultrasound scan, with no paternal association ^49^. It is possible that potential differences in misreporting smoking and alcohol consumption between mothers and fathers could produce spurious parental differences. Pregnant women are likely to underreport whether they smoke or drink alcohol and the amount they smoke or drink, because of the social stigma of these, particularly in recent decades. As the report of alcohol and smoking in the LifeCycle cohorts was collected early in pregnancy it is likely to be random in relation to an offspring CHD as the vast majority would not have been diagnosed. Hence, this underreporting would be expected to attenuate any true effect of smoking/alcohol on CHD towards the null. This misclassification is less likely in fathers. Thus, the specific positive association of maternal smoking on CHDs and its difference to the paternal association may be underestimated.

In summary, we found evidence to support a causal intrauterine effect of maternal smoking on any CHD, particularly with non-severe CHDs, but did not find robust evidence for a causal effect of maternal BMI or alcohol on offspring CHD risk. Whilst everyone should be encouraged not to smoke, and all clinical guidelines advocate not starting smoking, and if women do smoke, to quit before becoming pregnant, there are still high rates of smoking in some groups, particularly those from deprived backgrounds. In the studies included in this paper, two contemporary cohorts, BASELINE (Ireland), with births occurring between 2008 and 2011 and BiB (UK), with births occurring between 2007 and 2011, smoking prevalence rates were 25% and 16% respectively. The prevalence in BiB masks the high rate in white British women (33%) who are from socioeconomically deprived backgrounds, as over 50% of births in that cohort are to Pakistani women who have low rates of smoking (3%) ^17^. It is possible that emphasizing the potential adverse effect on CHDs in specific groups might help in supporting women of reproductive age not to start smoking and women who are smoking at the start of pregnancy to be encouraged to quit. Furthermore, understanding the specific mechanisms that link maternal smoking to increased offspring CHD risk could identify targets for interventions for its prevention.

## Data Availability

A manuscript describing in detail the LifeCycle project can be found at: https://link.springer.com/article/10.1007/s10654-020-00662-z. The LifeCycle Project or EU Child Cohort Network do not own data, but bring data from other cohorts together via a federated data analysis platform. Ethical and legal responsibility for data management and security is maintained by the source studies or home institutions. The principal investigators or home institutions should always administer permission for external access to specific data on their server for addressing research questions. The EU Child Cohort Network cannot provide open access to researchers. The data sharing protocols and agreements will be updated regularly, according to new legal practices, such as the European General Data Protection Regulation 2016/679 (GDPR). All governance protocols will take not only the short-term, but also the long-term EU Child Cohort Network, beyond the LifeCycle Project duration, into account.

## Acknowledgements

The authors would like to acknowledge everyone who has supported and contributed to each cohort included in this study. Please see Text S6 (Supplementary Material) for a full list of acknowledgments.

