## Supplementary Material for "The effect of maternal pre-/early-pregnancy BMI and pregnancy smoking and alcohol on congenital heart diseases: a parental negative control study"

**Contents and navigation menu**

**Supplementary methods** ........................................................2-13

Text S1. Cohort descriptions ........................................................2-4

Text S2. Data description and methods ........................................................5-7

Text S3. Paternal alcohol consumption ........................................................8

Text S4. Defining congenital outcomes ........................................................9-12

Text S5. Confounder data ........................................................13-14

**Supplementary Results** ........................................................14-45

Participant flow charts for each cohort ........................................................15-21

Missing data and complete-case analyses ........................................................22-28

Supplementary results (BMI) ........................................................29-39

Supplementary results (Smoking) ........................................................40-43

Supplementary results (Alcohol) ........................................................44-48

**Text S6. Acknowledgements**........................................................49

**References** ........................................................50-51

### Supplementary methods

#### ***Text S1. Cohort descriptions***

##### *The Amsterdam Born Children and their Development Study (ABCD)*

The following text was adapted from the ABCD cohort profile where full study details are described (https://doi.org/10.1093/ije/dyq128) [1]:

Between January 2003 and March 2004, all pregnant women living in Amsterdam were asked to participate in the ABCD study during their first prenatal visit to an obstetric care provider (general practitioner, midwife or gynaecologist). Altogether, 12 373 women were approached—by estimate, ≥99% of the target population. According to Dutch law, all pregnant women, including illegal immigrants and asylum-seekers, are entitled to receive prenatal care, which is free of charge if costs are a problem. For all of the women approached, the care provider completed a registration form which included personal data such as name, address and date of birth. Based on this information, a questionnaire covering socio-demographic characteristics, obstetric history, lifestyles and psychosocial conditions was sent to the pregnant women within 2 weeks, to be filled out at home and returned to the Public Health Service by prepaid mail. A reminder was sent 2 weeks later. The questionnaire included an informed consent sheet the women could use to grant permission for follow-up of their infants at the age of 3 months and every 5 years thereafter, and for the perusal of their medical files. Approval for the ABCD study was obtained from the Central Committee on Research involving Human Subjects in the Netherlands, the Medical Ethical Committees of the participating hospitals, and from the Registration Committee of the Municipality of Amsterdam. Written informed consent was obtained from all participating mothers.

Of the 12 373 women approached, 8266 women filled out the pregnancy questionnaire (response rate: 67%). Of this group, 7050 women granted permission for follow-up (85%) and 7043 women granted permission for perusal of her and her child's medical files (85%). To enhance participation among foreign-born women, two supportive measures were taken: (i) a Turkish, Arabic or English translation was provided to women born in Turkey, Morocco or other non-Dutch-speaking countries and (ii) the possibility of completing the questionnaire orally was offered to women who were illiterate or had reading difficulties.

##### *The Avon Longitudinal Study of Parents and Children (ALSPAC)*

ALSPAC is a prospective birth cohort study which was devised to investigate the environmental and genetic factors of health and development. Detailed information about the methods and procedures of ALSPAC is available elsewhere [2–4]. 14,541 pregnant women with an expected delivery date of April 1991 and December 1992, residing in the former region of Avon, UK were eligible to take part. Additional enrolment provided a baseline sample of 14,901 participants [4]. The study website contains details of all the data that is available through a fully searchable data dictionary. Ethical approval for the study was obtained from the ALSPAC Ethics and Law Committee and the Local Research Ethics Committees (http://www.bristol.ac.uk/alspac/researchers/research-ethics/). Informed consent for the use of data collected via questionnaires and clinics was obtained from participants following the recommendations of the ALSPAC Ethics and Law Committee at the time.

***The Cork SCOPE BASELINE Birth Cohort Study (BASELINE)***

The following text was adapted from the BASELINE cohort profile where full study details are described: https://doi.org/10.1093/ije/dyu157 [5].

The study is based in Cork, Ireland. The SCOPE Ireland pregnancy cohort formed the basis of recruitment of infants to BASELINE (n = 1537). In 2007, the amalgamation of all three Cork maternity units into one centre, Cork University Maternity Hospital (CUMH), provided a unique opportunity to conduct research in pregnancy in Cork. CUMH, which is co-located with the Cork University Hospital, is the third largest maternity hospital in Ireland, with 8563 deliveries in 2012. As recruitment was regionally based, the generalizability of the data may be limited. In 2008, all primiparous women in Cork were invited to take part in the Screening for Pregnancy Endpoints (SCOPE) pregnancy cohort. The SCOPE cohort is an international collaboration of research groups interested in the study of major adverse outcomes in late pregnancy, particularly but not exclusively, pre-eclampsia, fetal growth restriction and spontaneous preterm birth8 and as a consequence strict exclusion criteria were applied.9 Detailed maternal, fetal and paternal information was obtained antenatally, as well as blood samples at 15 and 20 weeks' gestation, see Table 1. All women who participated in the SCOPE study were informed about the birth cohort, and if consent was obtained infants were registered to the Cork BASELINE birth cohort.

***The Born in Bradford Cohort (BiB)***

The Born in Bradford study is a population-based prospective birth cohort including 12,453 women who experienced 13,776 pregnancies between 2007 and 2011. The study is unique in that it has almost an equal split between White European and South Asian women, all residing in Bradford, UK. Bradford is a city in the North of England with high levels of socioeconomic deprivation, and the cohort was started due to a high prevalence of poor child health in the city [6]. Full details of the study methodology were reported previously [7]. The study website provides more information, including protocols, questionnaires and information on how researchers can access data and a full list of all available data (https://borninbradford.nhs.uk/research/documents-data/). Mothers, and their partners, recruited into the study provided detailed interview questionnaire data, measurements, and biological samples. They also consented to the linkage of theirs and their child's data.

***The Danish National Birth Cohort (DNBC)***

The DNBC is a nationwide cohort of pregnant women, recruited from 1996 through 2002 consisting of 100,415 pregnancies [8]. Informed consent was obtained from participants upon enrolment, and the study was approved by the Danish Data Protection Agency through the joint notification of the Faculty of Health and Medical Sciences at the University of Copenhagen (Sund-2017-09), according to Danish regulations. Information on lifestyle and environmental factors potentially associated with offspring health was collected through 4 prenatal and postnatal telephone interviews at target ages gestational weeks 12 and 30 and child ages 6 and 18 months. The parent-child dyads were then invited for follow-up at 7, 11, and 18 years.

***The Norwegian Mother, Father and Child Cohort Study (MoBa)***

MoBa is a nationwide, pregnancy cohort comprising family triads (mother-father-offspring) who are followed longitudinally. All pregnant women in Norway who were able to read Norwegian were eligible for participation. The first child was born in October 1999 and the last in July 2009. Invitations were sent to women in 277 702 pregnancies, the participation rate was 41%. The cohort includes more than 114 000 children, 95 000 mothers and 75 000 fathers[9,10]. Extensive longitudinal data were collected using nine questionnaires: three during pregnancy, and then follow-up questionnaires when the children were 6 months, 18 months, 36 months, 5 years, 7 years and 8 years of age. In addition, a single questionnaire was administered to fathers during gestational weeks 15-18. Data collected include general background and health information, including diet and lifestyle, a semi-quantitative food frequency questionnaire, information on birth and pregnancy outcomes, and on several aspects of child nutrition and development, as well as the physical and mental health of both mother and child. MoBa is linked to the Medical Birth Registry of Norway, which provides standardized information about the health of the mother during pregnancy, other essential medical information related to the pregnancy and birth, and standard post-natal measures of the child. The establishment of MoBa and initial data collection was based on a license from the Norwegian Data Protection Agency and approval from The Regional Committees for Medical and Health Research Ethics. The MoBa cohort is now based on regulations related to the Norwegian Health Registry Act.

***NINFEA study***

The NINFEA study is internet-based birth cohort established in 2005 in Italy (http://www.progettoninfea.it) [11–13]. The cohort consists of children born to mothers who have access to the internet and enough knowledge of Italian to complete online questionnaires. The recruitment is conducted actively, through obstetrics clinics, and passively, via internet and the media. A baseline questionnaire on general health and exposures before and during pregnancy is completed by mothers at enrolment, which may occur at any time during pregnancy. Further follow-up information is obtained with repeated questionnaires completed 6 and 18 months after delivery and when children turn 4, 7, 10 and 13 years.

#### *Text S2. Data description and methods*

**Table S1.** Study-specific methods for data collection.

| ***Measurement*** | ***Study-specific details*** |
|---|---|
| *BMI data* | |
| Maternal BMI | **ABCD:** Women filled out a questionnaire containing questions on sociodemographic characteristics, medical history, lifestyle and dietary habits (16 weeks of gestation; IQR 12–20 weeks). BMI was based on pre-pregnancy height and weight as reported in the pregnancy questionnaire.<br>**ALSPAC:** In the 2nd pregnancy questionnaire (12 weeks' gestation) women were asked to report their pre-pregnancy weight and height. No definition of pre-pregnancy was provided in the question. Subsequently for the majority of women all weight measurements from any time of pregnancy have been extracted from obstetric records (height was not routinely measured antenatally in the UK when these women were pregnant). First antenatal clinic measurements of weight correlated strongly with the women's self-report (Pearson correlation = 0.93).<br>**Baseline:** At 15 weeks' gestation sociodemographic and anthropometric measurements, including objectively measured weight and height, were collected.<br>**BiB:** Weight and height (unshod and in light clothing and following a standard protocol) were measured at the recruitment assessment. As women were recruited at the oral glucose tolerance test (26-28 weeks of gestation for the majority) this would not provide an accurate measure of pre-/early-pregnancy weight and would include fetal and amniotic weight and pregnancy related weight gain. All measurements of weight from all antenatal clinics were extracted from the obstetric records and pre-/early-pregnancy BMI was calculated using weight from the first antenatal clinic (median 12 weeks' gestation) and height at recruitment (26-28 weeks' gestation).<br>**DNBC:** Self-reported information on pre-pregnancy weight and height from the first pregnancy interview at around 16 weeks gestation.<br>**MoBa:** Pre-pregnancy weight and height were self-reported during the first interview at week 17 in pregnancy.<br>**NINFEA:** Pre-pregnancy weight and height were self-reported within the Q1 questionnaire which can be completed at any time during pregnancy. |
| Paternal BMI | **ABCD:** Paternal weight was maternally reported in questionnaire when child was aged 5-6 years (the closest timepoint available to pregnancy). Paternal height was maternally reported in the pregnancy questionnaire at around 16 weeks' gestation.<br>**ALSPAC:** Paternal weight and height were self-reported from the first partner questionnaire completed around 18 weeks' gestation.<br>**Baseline:** Paternal weight and height were measured around the time of pregnancy. |

| | |
|---|---|
| | **BiB:** Paternal weight and height were self-reported from the first partner questionnaire mostly completed at recruitment (26–28 weeks' gestation).<br>**DNBC:** Paternal weight and height were maternally reported by interview when the child 18 months.<br>**MoBa:** Paternal weight and height were maternally reported by questionnaire at around 18 weeks' gestation.<br>**NINFEA:** Paternal weight and height were maternally reported within the Q1 questionnaire which can be completed at any time during pregnancy. |
| *Smoking data* | |
| Maternal smoking | **ABCD:** Asked number of cigarettes per day during pregnancy in first questionnaire (16 weeks of gestation; IQR 12–20 weeks). Binary variable used any smoking during pregnancy.<br>**ALSPAC:** Asked number of cigarettes per day during pregnancy in questionnaire at around 18 weeks' gestation. Binary variable used any smoking during the first trimester.<br>**Baseline:** Reported in early pregnancy questionnaire around 14 weeks gestation. Binary variable used any smoking during the first trimester. Baseline smoking data only used to adjust for BMI analyses.<br>**BiB:** Asked number of cigarettes per day during pregnancy in first questionnaire (26-28 weeks' gestation). Binary variable used any smoking during pregnancy.<br>**DNBC:** Maternal smoking in the first trimester was ascertained from a computer-assisted telephone interview conducted at approximately 16 weeks' gestation. Binary variable used any smoking during the first trimester.<br>**MoBa:** Smoking habits were assessed from questionnaires sent by mail at 13-17 and 30 weeks. Binary variable used any smoking during pregnancy.<br>**NINFEA:** Smoking habits in the first two trimesters were assessed in the baseline questionnaire, compiled during pregnancy (could be completed any time during pregnancy). Binary variable used any smoking during the first trimester. |
| Paternal smoking | **ABCD:** NA<br>**ALSPAC:** Asked about smoking habits within the partner questionnaire during pregnancy at around 18 weeks' gestation.<br>**Baseline:** Maternally reported in pregnancy questionnaire around 14 weeks' gestation.<br>**BiB:** Asked about smoking habits within partner questionnaire during pregnancy (26-28 weeks' gestation).<br>**DNBC:** Maternally reported at 16 weeks' gestation.<br>**MoBa:** Self-reported within first partner questionnaire around 15 weeks' gestation.<br>**NINFEA:** NA |
| *Alcohol data* | |
| Maternal alcohol | **ABCD:** Mothers asked how many glasses of alcohol they drunk during first period of pregnancy (16 weeks of gestation; IQR 12–20 weeks). Binary variable used any alcohol intake during pregnancy. |

| | |
|---|---|
| | **ALSPAC:** Self-reported from pregnancy questionnaire at around 18 weeks' gestation. Binary variable used any alcohol intake during the first trimester.<br>**Baseline:** Reported in early pregnancy questionnaire around 14 weeks gestation. Binary variable used any alcohol intake during the first trimester. Baseline alcohol data only used to adjust for BMI analyses.<br>**BiB:** NA<br>**DNBC:** Self-reported at 16 weeks' gestation. Binary variable used alcohol intake during the first trimester.<br>**MoBa:** Assessed via questionnaire around 17 weeks' gestation. Binary variable used any alcohol intake during the first trimester.<br>**NINFEA:** Drinking habits in the first trimester were assessed in the baseline questionnaire (completed at any time during pregnancy). Binary variable used any alcohol intake during the first trimester. |
| Paternal alcohol | **ALSPAC:** Self-reported within first partner questionnaire at around 18 weeks' gestation.<br>**MoBa:** Self-reported within first partner questionnaire at around 15 weeks' gestation. |

#### ***Text S3. Paternal alcohol consumption***

***ALSPAC***

We used data from the partners questionnaire which was filled in by partners at around 18 weeks' gestation. We used data from questions B18 and B19 from the PB questionnaire (http://www.bristol.ac.uk/alspac/researchers/our-data/).

*B18b. How often have you drunk alcoholic drinks during the last 3 months: 1) Never, 2) less than once a week, 3) at least once a week, 4) 1-2 glasses every day, 5) 3-9 glasses every day, 6) at least 10 glasses every day.*

*B19b. How many days in the past month did you drink the equivalent of 2 pints of beer, 4 glasses of wine or 4 pub measures of spirit? 1) Every day, 2) more than 10 days, 3) 5-10 days, 4) 3-4 days, 5) 1-2 days, 6) none.*

We coded paternal alcohol consumption as follows: non-drinkers = If answered 1 to B18b; light drinkers = answered 5 to B19b; mod/heavy drinkers = answered 1,2,3 or 4 to B19b.

***MoBa***

Question *FF244. How often do you drink alcohol now that your partner is pregnant? Response options: 1) Approximately 6-7 times per week, 2) Approximately 4-5 times per week, 3) Approximately 2-3 times per week, 4) Approximately once per week, 5) Approximately 1-3 times per month, 6) Less than once per month, 7) Never.*

Using data from FF244**,** we coded paternal alcohol consumption as follows: non-drinkers = Answered number 7; light drinkers = Answered 4, 5 or 6; mod/heavy drinkers = Answered 1, 2 or 3

#### ***Text S4. Definition of congenital heart disease (CHD) and other congenital anomalies (CAs)***

Here we describe ascertainment of CA cases for each cohort. International Classification of Diseases (ICD; version 10) codes were used to define CA cases when possible. However, in some cohorts these data were not available. The following cohorts were used to define CA cases with ICD codes: ALSPAC, BiB, DNBC, NINFEA.

##### ***ABCD***

The ABCD cohort has previously published research involving CAs [14]. The same methods for data extraction were used for the present study. Data on CAs were obtained from three different sources: the infant questionnaire, which was filled out by the mother at an average infant age of 12.9 weeks (IQR 12.4–13.4 weeks); the questionnaire filled out by the mother at an average infant age of 5.07 years (IQR 5.04–5.13 years), and clinical data of the Youth Health Care Registration (health and development registration of all children in the Netherlands, which is mandatory under the law on medical treatment agreement). The questionnaires were screened by a researcher, and in the case of missing or unclear answers the mothers were contacted. Subsequently, the questionnaires were scanned and transferred to a database by a certified company (Scan serv, Nootdorp, the Netherlands). Missing data in the questionnaires could be supplemented by data from the Youth Health Care Registration, and in the case of any discrepancy the data from the Youth Health Care Registration prevailed. CA data in ABCD was restricted to live-born children.

CAs were categorized as follows: 0 = no defect 1 = congenital malformations of the nervous system 2 = congenital malformations of eye, ear, face, throat 3 = congenital malformations of the cardiovascular system 4 = congenital malformations of the respiratory tract 5 = split lip and/or palate 6 = congenital malformations of the digestive tract 7 = congenital malformations of the kidneys, urinary tract, genitalia 8 = congenital malformations of the musculoskeletal system 9 =neoplasms 10 = other congenital malformations 11 = chromosomal defect 12 = monogenic defect 13 = microdeletions and uniparental disomy 14 = other syndromes 15 = complex cardiovascular defects 16 = multiple defects of the extremities 17 = other multiple defects within an organ system 18 = multiple defects (in multiple organ systems) 21 = minor defect 22 = unclear/uncertain diagnosis 23 = "don't know which defect" 24 = "not applicable" 25 = missing information.

We coded CHD cases if they were “Yes” for category 3. We coded chromosomal/genetic aberrations if “Yes” for any of the following categories: 11, 12, 13, 14.

##### ***ALSPAC***

Case ascertainment of CAs in the ALSPAC cohort has been described in detail in a recently published data note [15]. Data were combined from multiple sources: NHS records (primary care, paediatric cardiology database, data on fetal deaths and local child health services), midwifery and birth records and maternal self-report via child-based questionnaires. Each source was coded using ICD-10 codes. By combining sources, there would be a greater possibility of capturing all of possible cases within the cohort. The majority of cases of CAs were identified by primary care records (79% for any CA and 68% for any CHD). We included diagnoses made at any age (from birth up until age 25/26). There were no restrictions in cases of CAs in ALSPAC, we included all cases whether live-born or not. However, it is possible that some cases that were terminated earlier in pregnancy were missed due to them never having an NHS number and thus not being identified through record linkage.

***BASELINE***

At 2 months, mothers were asked of any medical problems and/or referrals. If a baby had been referred to a specialist, it was checked to see if they had results from an echocardiogram. Echocardiograms were checked by a cardiologist. Exact CHD diagnoses were reported based on the echo. At 6 months, there was one additional baby that had cardiac surgery and added as a case. If a baby had been diagnosed after 6 months, they would have been identified through records on the Echo. Therefore, in BASELINE we obtained all CHDs up until ~age 12.

***BiB***

In the BiB cohort, there were two separate sources to identify CAs. Both sources were used in this study: (i) CAs up to 5 years of age, identified in GP records by Bishop et al [16] following EUROCAT guidelines. ICD-10 codes were mapped to clinical term (CT)-V3 codes prior to extraction from GP records. (ii) Data extracted from the Yorkshire and Humber CAs register database. Data were ICD-10 coded. All of these were confirmed postnatally. BiB includes data on the birth outcome of each child (live birth, miscarriage, still birth). Therefore, diagnoses were not necessarily restricted to live born children. However, there is the possibility that some women would have terminated the pregnancy after the 12- or 20-week scans which would lead to an under-representation of congenital anomaly cases.

***DNBC***

In the DNBC, all diagnoses of congenital anomalies (according to EUROCAT guide 1.4 section 3.2 and 3.3) up until the age of 15 years were extracted from the Danish National Patient Register (DNPR) which is linked to the cohort data[17,18]. Diagnoses were ICD-coded. These data were restricted to children born alive.

***MoBa***

Information on whether a child had a CHD or not was obtained though linkage to the Medical Birth Registry of Norway (MBRN). All maternity units in Norway must notify births to the MBRN. The notification form includes the name and personal identity number of the child and parents, as well as information about maternal health before and during pregnancy, and any complications during pregnancy or at birth, including the presence of any heart defects. The MBRN contains information on all births and pregnancies ended after the 12th week of gestation, including stillbirths and abortions after the 12th week, including on heart defects. Heart defects are registered in the MBRN through notifications from clinical staff identifying these defects at delivery or any hospital in patient treatments occurring immediately after birth until the child is discharged. The medical notification is made at discharge, which can be several months after birth. Details of the notified heart defects, such as specific diagnosis or treatment are not provided. Whilst most of the heart defects would have been diagnosed at birth it is possible that some children were admitted to hospital after delivery for non-specific reasons of for diagnoses that at the time were not considered to be related to a heart defect. Therefore, MOBA contribute only to analyses of any CHD and we considered diagnosis to have been made between birth and 6 months (few would remain in hospital after this length).

***NINFEA***

Congenital anomalies in the NINFEA cohort were reported in the second questionnaire compiled 6 months after birth. Mothers compiled a checklist that included pre-specified anomalies (namely cryptorchidism (also assessed 18 months after birth), congenital hip dysplasia, cleft palate, spina bifida and pyloric stenosis) and anomalies divided by major systems (namely cardiovascular, gastrointestinal, genitourinary, musculoskeletal, respiratory and nervous system, and genetic/chromosomal or metabolic/endocrine disease). If the mother reported an anomaly from a specific system, the exact name of the anomaly was asked. If the child died or had any surgery performed in the first 6 months, the cause of death and type of surgery were also checked to see if any congenital anomaly was reported. All congenital anomalies were coded using ICD-10 codes by an experienced pediatrician and were reassessed by an independent MD. NINFEA included live-born infants only.

***Studies with ICD coded data***

Table S2 shows how cases of CHD were defined in the studies with ICD codes (ALSPAC, BiB, DNBC, NINFEA).

**Table S2.** Subcategories of CHD.

| Category | CHDs included/excl | ICD codes |
|---|---|---|
| All | Any CHD as defined by EUROCAT*<br>Patent ductus arteriosus (PDA) with gestational age (GA) < 37 weeks not considered a CHD case.<br>Peripheral pulmonary artery stenosis with GA < 37weeks not considered as a CHD case. | Q20-Q25, Q260, Q262-Q269** |
| Severe | Heterotaxia, Conotruncal defect, Atrioventricular septal defect, Anomalous pulmonary venous return, Left ventricle outflow tract obstruction, Right ventricle outflow tract obstruction, Other complex defects | Q240, Q241, Q206, Q200, Q251, Q252, Q253, Q254, Q203, Q213, Q201, Q214, Q212, Q26, Q262, Q264, Q268, Q269, Q234, Q251, Q230, Q231, Q221, Q224, Q225, Q255, Q204 |
| Non-severe | PDA (in full term infants), valvular pulmonary stenosis, ventricular septal defect (VSD), atrial septum defects (ASD), unspecified septal defects, isolated valve defects, other specified heart defects, unspecified heart defects | Non-severe cases that are All=1 and Severe=0. |

* Definitions taken from here: https://eu-rd-platform.jrc.ec.europa.eu/sites/default/files/EUROCAT-Guide-1.4-Section-3.3.pdf
**Q250 and Q256 not a case if isolated and GA<37weeks

***Additional analysis - excluding infants with any known chromosomal/genetic/teratogenic defects***

ABCD, ALSPAC, BiB, DNBC, MoBa and NINFEA contributed to this additional analysis. In ALSPAC, BiB, DNBC and NINFEA, we used the ICD codes in Table S3 to exclude cases. In ABCD, there were specific categories (described above) which corresponded to chromosomal and genetic anomalies (11 = chromosomal defect 12 = monogenic defect 13 = microdeletions and uniparental disomy 14 = other syndromes). In MoBa, we used questionnaire data which was maternally reported at 6 months after birth: “Is your child suspected of having a syndrome?” and “Is your child suspected of having a chromosomal defect?”.

**Table S3.** Subcategories of congenital anomalies with a ‘known cause’ used in additional analyses.

| **Category** | **ICD-10 Codes** |
|---|---|
| Teratogenic/genetic syndromes, microdeletions and chromosomal abnormalities (additional analysis). | D821, P350-P352, P371, Q619, Q751, Q754, Q771-Q772, Q780, Q796, Q85, Q861-Q869, Q87, Q90-Q92, Q930-Q939, Q95-Q99 |

#### ***Text S5. Confounder data***

The maximum number of confounders used in fully adjusted models are listed below. Confounder and other parent exposure adjusted models are the same as fully adjusted but with additional adjustment for the other parent's exposure and additional adjustment for maternal parity in paternal models.

Exposure = BMI: age, education, parity (maternal), ethnicity, smoking, alcohol, offspring sex.

Exposure = Smoking: age, education, parity (maternal), ethnicity, alcohol, offspring sex.

Exposure = Alcohol: age, education, parity (maternal), ethnicity, smoking, offspring sex.

There is evidence that smoking and alcohol influence BMI [19–22]. We therefore treated those as confounders for the association of maternal/paternal BMI with CHD. Smoking and alcohol are associated with each other in most populations but whether one causes the other is unclear. It is possible that most of their association is due to socioeconomic and cultural factors. Despite being unclear about whether they could be confounders of each other's effect on CHD (e.g. alcohol a confounder for smoking and vice versa) in the final confounder adjusted model we included alcohol as a confounder for smoking and vice versa.

We used maternal/paternal age at birth in complete years. We used educational attainment for both parents' measures of socioeconomic position (SEP). In the harmonized LifeCycle data education has been defined according to the international classification (High: Short cycle tertiary, Bachelor, Masters, Doctoral or equivalent (ISCED-2011: 5-8, ISCED-97: 5-6) Medium: Upper secondary, Post-secondary non-tertiary (ISCED-2011: 3-4, ISCED-97: 3-4) Low: No education; early childhood; pre-primary; primary; lower secondary or second stage of basic education). Mothers parity was based on previous born children (previous stillbirths included, abortions excluded) (coded as 0, 1, 3, ≥ 4). For ethnicity we used the best estimate of the mother's/father's ethnic background based on the cohort's discretion (Western, Non-western, Mixed). Offspring sex was a binary variable (male/female). In additional analyses, we adjusted for folic acid supplementation in fully adjusted maternal models. This was a yes/no variable defined as intake of folic acids (folate, vitamin B9) during the period from conception to early pregnancy (12 weeks).

In NINFEA, due to the smaller sample size, maternal parity and maternal/paternal education were categorized as binary variables (parity: nulliparous and multiparous, education: low and medium combined together).

In ALSPAC, BASELINE, DNBC, MoBa and NINFEA we did not adjust for ethnicity in any analyses. 98% of women were of Western origin in ALSPAC. >98.5% of women in BASELINE were of Western origin. Ethnicity in the DNBC is said to be of >99% White European origin with a recent paper reporting their DNBC population to be 100% of White origin [23]. There were no data available on ethnicity in MoBa, however, it is believed that 99-100% are of Western origin. Ethnicity data were not available in NINFEA, although, the large majority of mothers (>98%) were born in Europe. Data on paternal country of birth was available for approximately half of the cohort and >98% of them were born in Europe.

In BiB only ~28% of mothers had harmonized data on alcohol intake during pregnancy, therefore this was not included in any models within BiB analyses as an exposure and also as a confounder in BMI and smoking models.

ABCD and BASELINE did not have harmonized LifeCycle data available. We describe methods for data harmonization here:

We used available ABCD data and tried to harmonize it as best as possible to match the LifeCycle data. BMI, sex, age, parity and folic acid supplementation were identical variables to the harmonized LifeCycle ones. Paternal height was self-reported by the mother and paternal weight was from 11 months after pregnancy (the closest timepoint available). We used any pregnancy smoking or drinking (yes/no) for the smoking and alcohol variables as there was no trimester specific exposure data. ABCD did not contribute to paternal alcohol or smoking analyses as there were no data for these exposures around the time of pregnancy. Maternal education was originally defined as a continuous variable as years of education after elementary school. We split this into 3 equal groups and defined as low, medium and high. Paternal education was from the 11-year questionnaire and split into 3 groups as this was the only data available. For ethnicity, we defined Western and non-western as appropriate from physiological ethnicity of grandmother's birth country for maternal ethnicity. Paternal ethnicity was reported by the mother and recoded to Western/Non-Western/Mixed.

All women were experiencing their first pregnancy in BASELINE; therefore we did not adjust for parity in any analyses. BMI, sex, age and smoking were coded the same as the harmonized LifeCycle data. Education in BASELINE was binary defined as medium or high. This was left unchanged and used as a measure of SEP as in other analyses.

In the analysis plan, we originally stated that we would treat type-1 diabetes (T1D) as a confounder. The rationale for this was that diabetes is a known teratogen for CHDs and could also influence pregnancy lifestyle factors through changes in behaviours. However, after exploring the data, the prevalence of T1D was low in those cohorts with data (0.2% in ALSPAC, 0.1% in BiB and 0.2% in DNBC for maternal T1D) and the other cohorts did not have data on specific diabetes diagnoses. For cohorts with T1D data, the number of CHD cases in those with a diagnosis was either zero or less than 10, making adjustment not meaningful or impossible through complete separation in the logistic model.

### Supplementary results

#### ***Participant flow charts for each cohort***

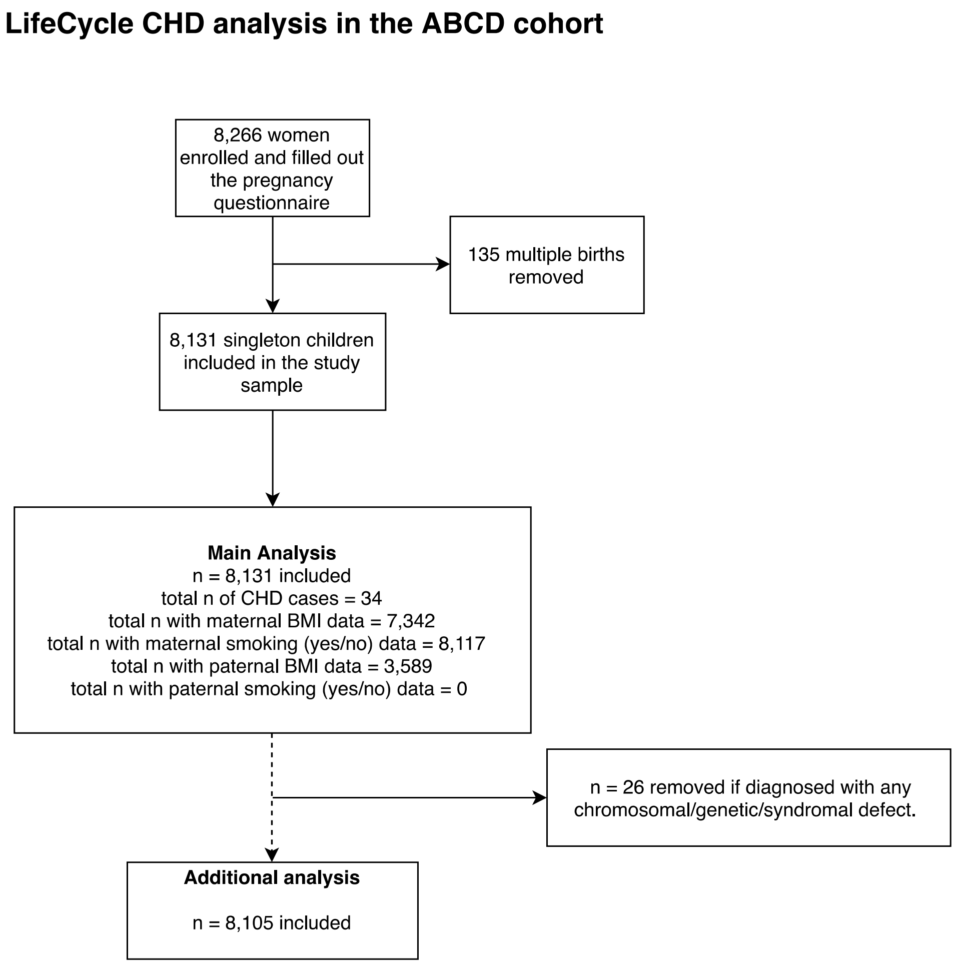

**Figure S1.** Study flow chart illustrating participant selection in the ABCD cohort.

**LifeCycle CHD analysis in the ALSPAC cohort**

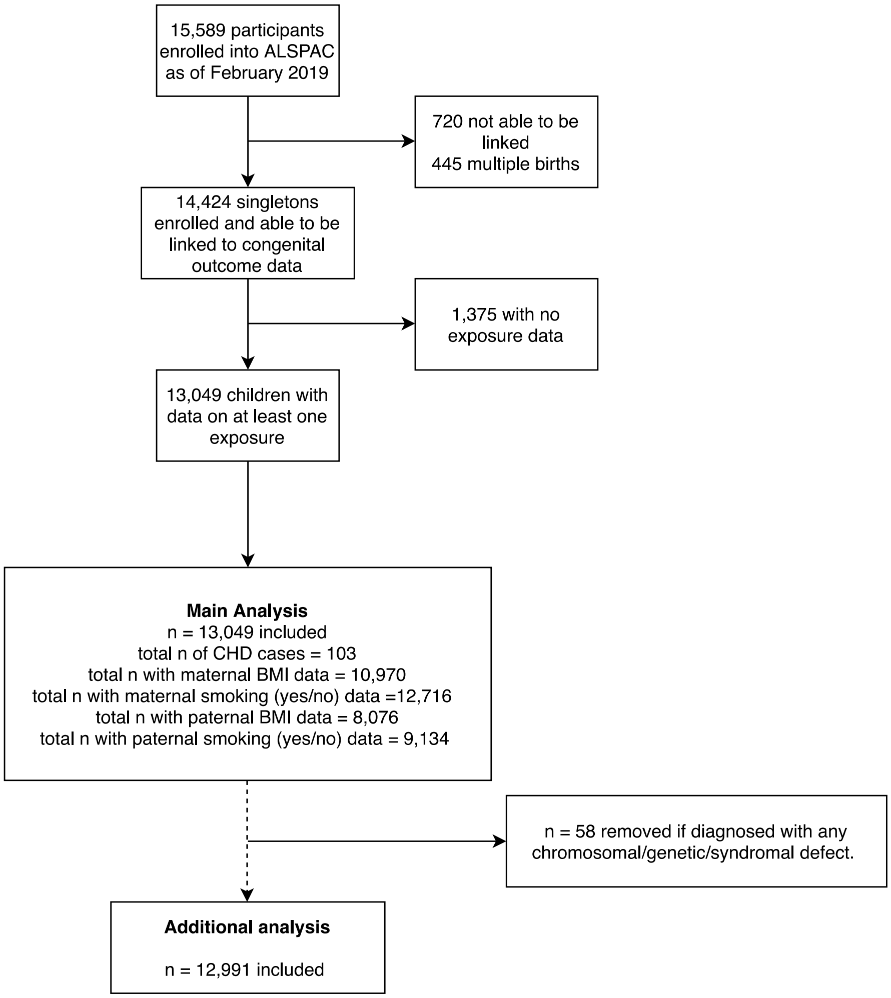

**Figure S2.** Study flow chart illustrating participant selection in the ALSPAC cohort.

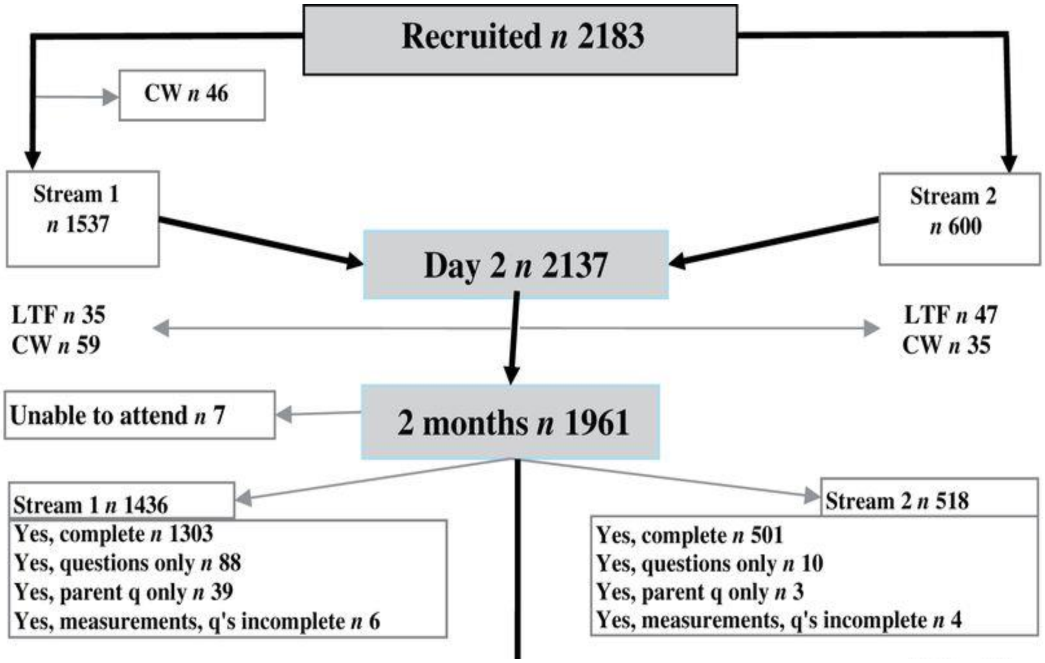

**Figure S3.** Study flow chart illustrating participant selection in the BASELINE cohort. We included 1436 participants in our study (Stream 1). Adapted from: https://doi.org/10.1093/ije/dyu157

**LifeCycle CHD analysis in the BiB cohort**

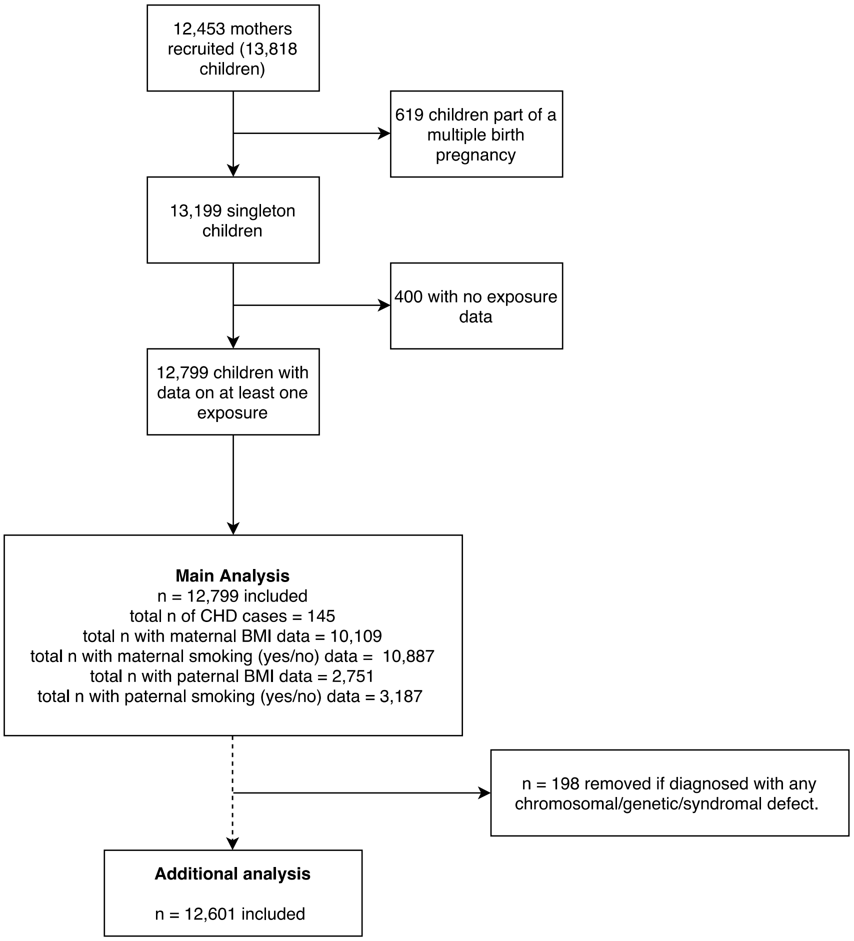

**Figure S4.** Study flow chart illustrating participant selection in the BiB cohort.

#### LifeCycle CHD analysis in the DNBC

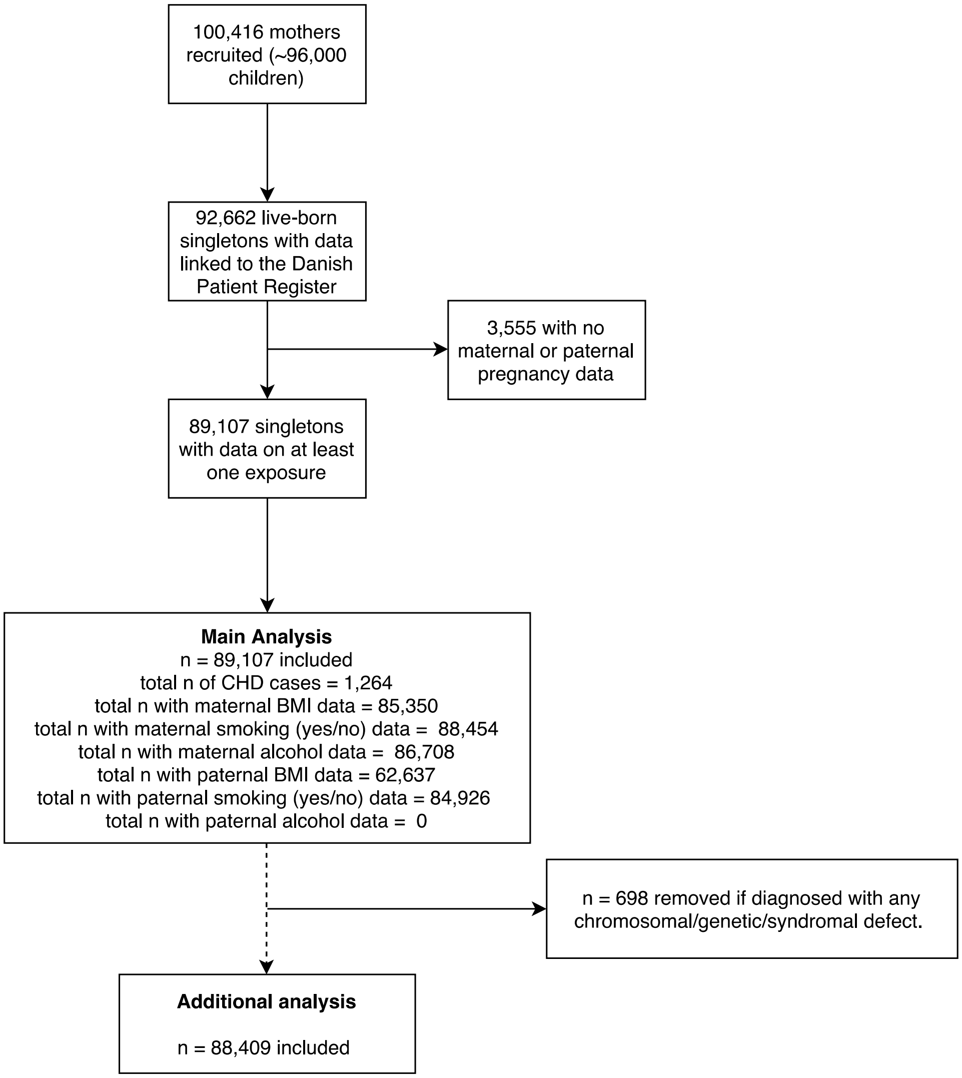

**Figure S5.** Study flow chart illustrating participant selection in the DNBC cohort.

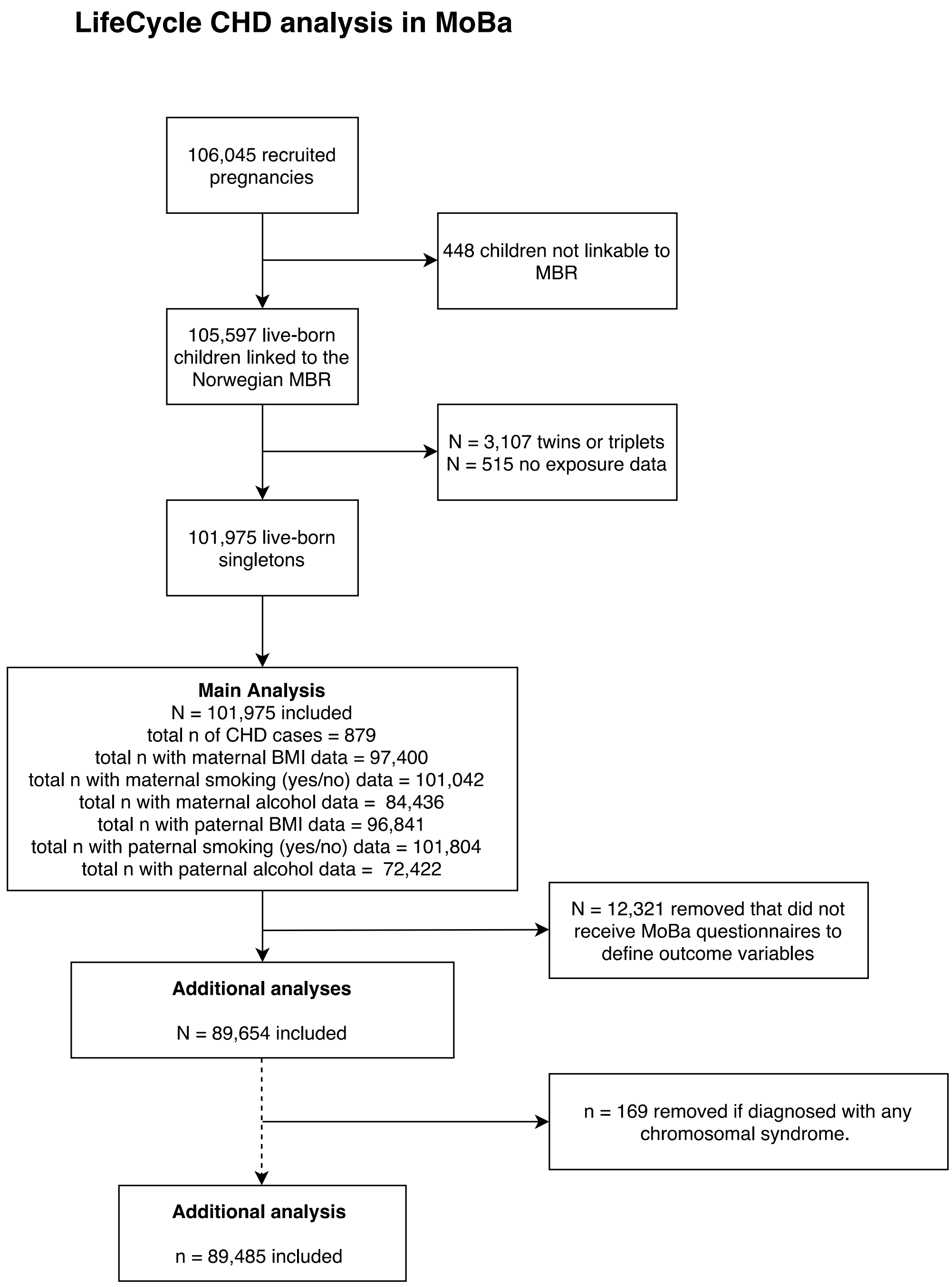

**Figure S6.** Study flow chart illustrating participant selection in the MoBa cohort. MBR = Medical birth registry.

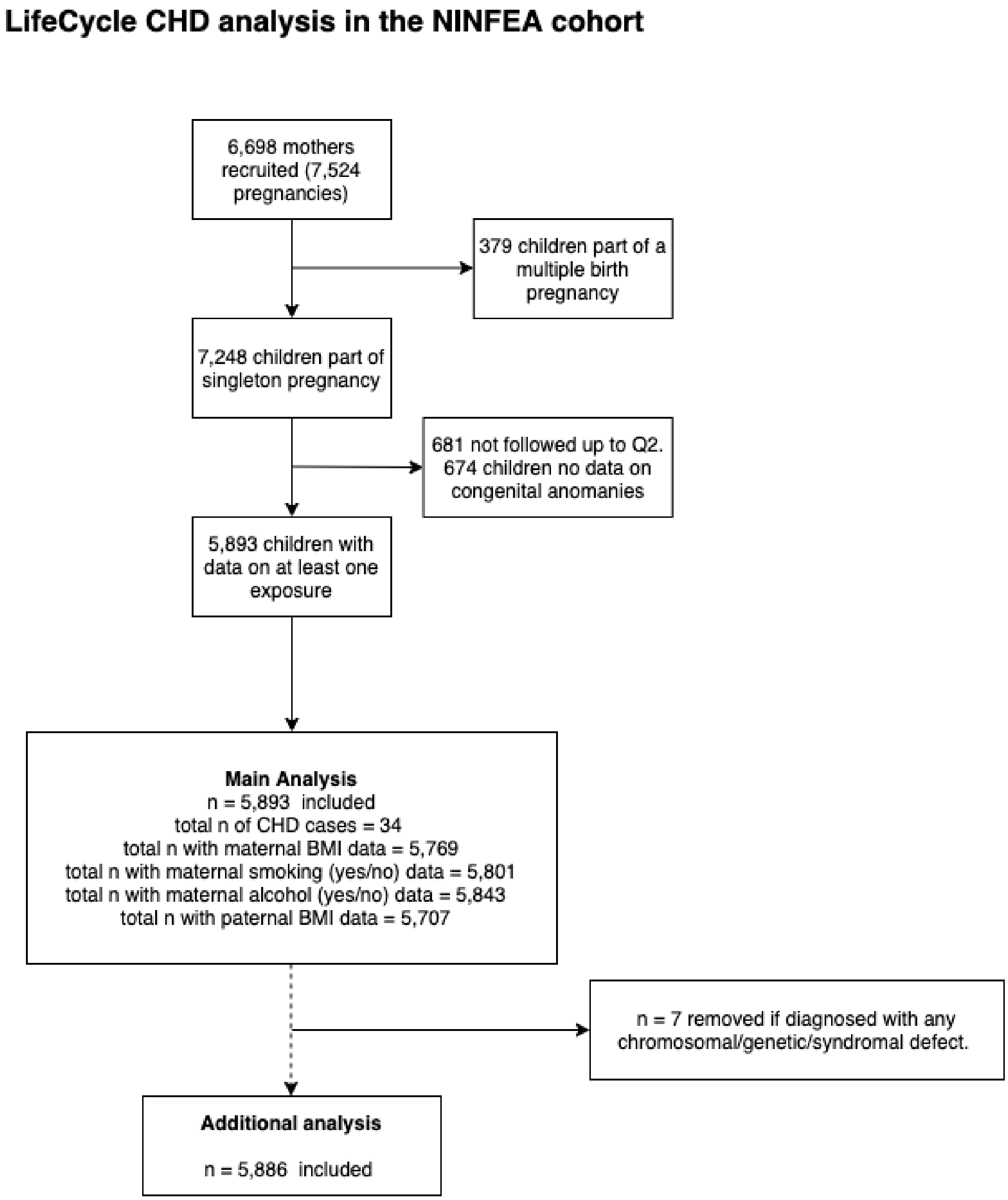

**Figure S7.** Study flow chart illustrating participant selection in the NINFEA cohort.

#### ***Missing data***

**Table S4.** Summary of missing data in each cohort.

| | ABCD<br>*N* = 8,131 | ALSPAC<br>*N* = 13,049 | BASELINE<br>*N* = 1,436 | BiB<br>*N* = 12,799 | DNBC<br>*N* = 89,107 | MoBa<br>*N* = 101,975 | NINFEA<br>*N* = 5,893 |
|---|---|---|---|---|---|---|---|
| Country | Netherlands | UK | RoI | UK | Denmark | Norway | Italy |
| Recruitment period | 2003-2004 | 1991-1992 | 2008-2011 | 2007-2011 | 1996-2002 | 1999-2008 | 2005-2016 |
| ***Maternal (n missing (%))*** | | | | | | | |
| Age, years | 0 | 2062 (15.8) | 0 | 0 | 0 | 181 (0.2) | 1 (0.0) |
| BMI, kg/m$^2$ | 789 (9.7) | 2079 (15.9) | 0 | 2690 (21.0) | 3757 (4.2) | 4575 (4.5) | 124 (2.1) |
| Preg smoking yes/no | 14 (0.2) | 333 (2.6) | 0 | 1912 (14.9) | 2367 (2.7) | 933 (0.9) | 92 (1.6) |
| Preg smoking heaviness | - | 2350 (18.0) | - | 1912 (14.9) | 1184 (1.3) | 390 (0.4) | 72 (1.2) |
| Preg alcohol yes/no | 6 (0.1) | 427 (3.3) | 43 (3.0) | - | 2399 (2.7) | 19617 (19.2) | 50 (0.9) |
| Preg alcohol heaviness | - | 6548 (50.2) | - | - | 758 (0.9) | 17539 (17.2) | 79 (1.3) |
| Parity | 0 | 502 (3.8) | 0 | 470 (3.7) | 0 | 1805 (1.8) | 272 (4.6) |
| Education | 83 (1.0) | 1152 (8.8) | 9 (0.6) | 2750 (21.5) | 8451 (9.5) | 6963 (6.8) | 46 (0.8) |
| Ethnicity | 14 (0.2) | - | 0 | 1906 (14.9) | - | - | - |
| Folic acid supp | 98 (1.2) | 424 (3.2) | - | - | 6510 (7.3) | 1805 (1.8) | 148 (2.5) |
| ***Paternal (n missing (%))*** | | | | | | | |
| Age, years | 4378 (53.8) | 5488 (42.1) | 321 (22.4) | 9439 (73.7) | 1371 (1.5) | 521 (0.5) | 2506 (42.5) |
| BMI, kg/m$^2$ | 4542 (55.9) | 4973 (38.1) | 321 (22.4) | 10074 (78.7) | 26470 (29.7) | 5134 (5.0) | 186 (3.2) |
| Smoking | - | 3915 (30.0) | 323 (22.5) | 9612 (75.1) | 4181 (4.7) | 171 (0.2) | - |
| Alcohol | - | 4844 (37.1) | - | - | - | 29553 (28.9) | - |
| Education | 5873 (72.2) | 1620 (12.4) | 0 | 4676 (36.5) | 10690 (12.0) | 5372 (5.3) | 138 (2.3) |
| Ethnicity | 197 (2.4) | - | 321 (22.4) | 9625 (75.2) | - | - | - |
| Offspring sex | 203 (2.5) | 0 | 0 | 0 | 0 | 196 (0.2) | 1 (0.0) |

#### ***Sensitivity analysis: complete-case analyses***

**Table S5.** Comparison between maximal numbers from **main analyses presented in the manuscript (black, top rows)** and **complete case models (red, bottom rows).** Results are odds ratios (95% CIs) of any offspring CHD per unit difference in BMI.

| Model | *ABCD* | *ALSPAC* | *BASELINE* | *BiB* | *DNBC* | *MoBa* | *NINFEA* | *Meta-analysis results* |
|---|---|---|---|---|---|---|---|---|
| ***Maternal BMI unadjusted*** | 1.02 (0.94, 1.09)<br>N = 7,342 | 1.05 (1.00, 1.09)<br>N = 10,970 | 1.07 (0.92, 1.20)<br>N = 1,436 | 1.01 (0.97, 1.04)<br>N = 10,109 | 1.02 (1.01, 1.03)<br>N = 85,350 | 0.99 (0.98, 1.01)<br>N = 97,400 | 0.93 (0.83, 1.03)<br>N = 5,769 | **1.01 (1.00, 1.02)**<br>**N = 218,376** |
| | 1.07 (0.95, 1.16)<br>N = 3,415 | 1.01 (0.93, 1.08)<br>N = 6,452 | 1.06 (0.87, 1.23)<br>N = 1,078 | 0.99 (0.89, 1.10)<br>N = 1,753 | 1.02 (1.00, 1.03)<br>N = 55,564 | 0.99 (0.97, 1.01)<br>N = 73,637 | 0.93 (0.83, 1.04)<br>N = 5,393 | **1.01 (0.99, 1.02)**<br>**N = 147,292** |
| ***Maternal BMI confounder adjusted*** | 1.04 (0.95, 1.11)<br>N = 7,103 | 1.05 (0.99, 1.10)<br>N = 9,179 | 1.08 (0.93, 1.21)<br>N = 1,386 | 1.02 (0.98, 1.05)<br>N = 7,279 | 1.02 (1.00, 1.03)<br>N = 78,180 | 0.99 (0.97, 1.01)<br>N = 75,448 | 0.94 (0.84, 1.05)<br>N = 5,476 | **1.01 (1.00, 1.02)**<br>**N = 184,051** |
| | 1.05 (0.93, 1.15)<br>N = 3,415 | 1.01 (0.94, 1.08)<br>N = 6,452 | 1.06 (0.87, 1.23)<br>N = 1,078 | 0.98 (0.87, 1.09)<br>N = 1,753 | 1.01 (1.00, 1.03)<br>N = 55,564 | 0.99 (0.97, 1.01)<br>N = 73,637 | 0.95 (0.85, 1.06)<br>N = 5,393 | **1.01 (0.99, 1.02)**<br>**N = 147,292** |
| ***Maternal BMI confounder and other parent BMI adjusted*** | 1.05 (0.93, 1.15)<br>N = 3,415 | 1.02 (0.94, 1.10)<br>N = 6,452 | 1.05 (0.85, 1.23)<br>N = 1,078 | 0.99 (0.88, 1.09)<br>N = 1,753 | 1.01 (1.00, 1.03)<br>N = 55,564 | 0.99 (0.97, 1.01)<br>N = 73,637 | 0.94 (0.84, 1.06)<br>N = 5,393 | **1.00 (0.99, 1.02)**<br>**N = 147,292** |
| | 1.05 (0.93, 1.15)<br>N = 3,415 | 1.02 (0.94, 1.10)<br>N = 6,452 | 1.05 (0.85, 1.23)<br>N = 1,078 | 0.99 (0.88, 1.09)<br>N = 1,753 | 1.01 (1.00, 1.03)<br>N = 55,564 | 0.99 (0.97, 1.01)<br>N = 73,637 | 0.94 (0.84, 1.06)<br>N = 5,393 | **1.00 (0.99, 1.02)**<br>**N = 147,292** |
| ***Paternal BMI unadjusted*** | 0.99 (0.84, 1.08)<br>N = 3,589 | 0.99 (0.91, 1.06)<br>N = 8,076 | 1.07 (0.86, 1.21)<br>N = 1,115 | 1.03 (0.94, 1.12)<br>N = 2,706 | 1.02 (1.00, 1.04)<br>N = 62,637 | 0.99 (0.97, 1.01)<br>N = 96,841 | 1.02 (0.92, 1.13)<br>N = 5,707 | **1.01 (0.99, 1.02)**<br>**N = 180,690** |
| | 1.04 (0.88, 1.11)<br>N = 1,732 | 0.97 (0.86, 1.07)<br>N = 5,044 | 1.07 (0.86, 1.21)<br>N = 1,113 | 1.01 (0.89, 1.13)<br>N = 1,572 | 1.02 (1.00, 1.04)<br>N = 53,922 | 0.99 (0.96, 1.01)<br>N = 67,071 | 0.96 (0.81, 1.13)<br>N = 3,166 | **1.01 (0.99, 1.03)**<br>**N = 133,620** |
| ***Paternal BMI confounder adjusted*** | 1.03 (0.84, 1.10)<br>N = 1,800 | 0.96 (0.86, 1.06)<br>N = 5,550 | 1.06 (0.86, 1.21)<br>N = 1,113 | 1.04 (0.93, 1.14)<br>N = 2,085 | 1.02 (1.00, 1.05)<br>N = 54,710 | 1.00 (0.97, 1.02)<br>N = 68,623 | 1.03 (0.89, 1.19)<br>N = 3,294 | **1.01 (1.00, 1.03)**<br>**N = 137,175** |
| | 1.03 (0.84, 1.10)<br>N = 1,732 | 0.97 (0.86, 1.08)<br>N = 5,044 | 1.06 (0.86, 1.21)<br>N = 1,113 | 1.04 (0.92, 1.16)<br>N = 1,572 | 1.02 (1.00, 1.04)<br>N = 53,922 | 1.00 (0.97, 1.02)<br>N = 67,071 | 0.96 (0.81, 1.14)<br>N = 3,166 | **1.01 (1.00, 1.03)**<br>**N = 133,620** |
| ***Paternal BMI confounder and other parent BMI adjusted*** | 1.03 (0.85, 1.10)<br>N = 1,732 | 0.97 (0.86, 1.08)<br>N = 5,044 | 1.05 (0.84, 1.21)<br>N = 1,113 | 1.04 (0.92, 1.15)<br>N = 1,572 | 1.02 (1.00, 1.04)<br>N = 53,922 | 1.00 (0.97, 1.02)<br>N = 67,071 | 0.99 (0.83, 1.18)<br>N = 3,166 | **1.01 (0.99, 1.03)**<br>**N = 133,620** |
| | 1.03 (0.85, 1.11)<br>N = 1,732 | 0.97 (0.86, 1.08)<br>N = 5,044 | 1.05 (0.84, 1.21)<br>N = 1,113 | 1.04 (0.92, 1.15)<br>N = 1,572 | 1.02 (1.00, 1.04)<br>N = 53,922 | 1.00 (0.97, 1.02)<br>N = 67,071 | (0.99, 0.83, 1.18)<br>N = 3,166 | **1.01 (0.99, 1.03)**<br>**N = 133,620** |

Covariates used for each study in fully adjusted models (mutually adjusted models the same as fully adjusted but with additional adjustment for the other parent's BMI and parity in paternal models);
ABCD: Maternal: offspring sex, age, education, parity, ethnicity, smoking, alcohol. Paternal: offspring sex, age, education, ethnicity.
ALSPAC: Maternal: offspring sex, age, education, parity, smoking, alcohol. Paternal: offspring sex, age, education, smoking, alcohol.
BASELINE: Maternal: offspring sex, age, education, smoking, alcohol. Paternal: offspring sex, age, smoking.
BiB: Maternal: offspring sex, age, education, parity, ethnicity, smoking. Paternal: offspring sex, age, education, ethnicity, smoking.
DNBC: Maternal: offspring sex, age, education, parity, smoking, alcohol. Paternal: offspring sex, age, education, smoking.
MoBa: Maternal: offspring sex, age, education, parity, smoking, alcohol. Paternal: offspring sex, age, education, smoking, alcohol.
NINFEA: Maternal: offspring sex, age, education, parity, smoking, alcohol. Paternal: offspring sex, age, education.

**Table S6.** Comparison between **maximal numbers (black, top rows)** and **complete case models (red, bottom rows)**. Results are odds ratios (95% CIs) of any offspring CHD for a BMI category in comparison to normal BMI. Categories: underweight (BMI <18.5 kg/m$^2$), normal weight (BMI 18.5 to <25 kg/m$^2$), overweight (BMI 25 to <30 kg/m$^2$) and obese (BMI ≥30 kg/m$^2$).

| Exposure | *ALSPAC* | *BiB* | *DNBC* | *MoBa* | *Meta-analysis results* |
|---|---|---|---|---|---|
| ***Maternal underweight unadjusted*** | 0.69 (0.26, 1.48)<br>N = 10,970 | 0.67 (0.17, 0.89)<br>N = 10,109 | 1.36 (1.05, 1.73)<br>N = 85,350 | 1.03 (0.70, 1.52)<br>N = 97,400 | **1.19 (0.97, 1.46)<br>N = 203,829** |
| | 0.63 (0.15, 1.78)<br>N = 6,452 | NA | 1.35 (0.95, 1.86)<br>N = 55,564 | 1.06 (0.66, 1.71)<br>N = 73,637 | **1.21 (0.92, 1.57)<br>N = 135,653** |
| ***Maternal underweight confounder adjusted*** | 0.63 (0.19, 1.57)<br>N = 9,179 | 0.64 (0.10, 2.11)<br>N = 7,360 | 1.33 (1.01, 1.71)<br>N = 79,288 | 1.06 (0.66, 1.71)<br>N = 75,448 | **1.20 (0.96, 1.50)<br>N = 171,275** |
| | 0.68 (0.16, 1.93)<br>N = 6,452 | NA | 1.34 (0.94, 1.84)<br>N = 55,564 | 1.08 (0.67, 1.74)<br>N = 73,637 | **1.21 (0.93, 1.58)<br>N = 135,653** |
| ***Maternal underweight confounder and other parent BMI adjusted*** | 0.65 (0.15, 1.84)<br>N = 6,452 | NA | 1.35 (0.95, 1.86)<br>N = 55,564 | 1.07 (0.67, 1.73)<br>N = 73,637 | **1.21 (0.93, 1.58)<br>N = 135,653** |
| | 0.65 (0.15, 1.84)<br>N = 6,452 | NA | 1.35 (0.95, 1.86)<br>N = 55,564 | 1.07 (0.67, 1.73)<br>N = 73,637 | **1.21 (0.93, 1.58)<br>N = 135,653** |
| ***Maternal overweight unadjusted*** | 1.23 (0.64, 2.20)<br>N = 10,970 | 1.35 (0.87, 2.08)<br>N = 10,109 | 1.24 (1.07, 1.42)<br>N = 85,350 | 1.01 (0.85, 1.20)<br>1.02 N = 97,400 | **1.15 (1.04, 1.28)<br>N = 203,829** |
| | 0.71 (0.21, 1.82)<br>N = 6,452 | 1.46 (0.41, 5.29)<br>N = 1,753 | 1.28 (1.07, 1.53)<br>N = 55,564 | 1.04 (0.86, 1.27)<br>N = 73,637 | **1.16 (1.02, 1.32)<br>N = 137,406** |
| ***Maternal overweight confounder adjusted*** | 0.85 (0.35, 1.80)<br>N = 9,179 | 1.34 (0.80, 2.22)<br>N = 7,360 | 1.23 (1.06, 1.42)<br>N = 79,288 | 1.06 (0.87, 1.29)<br>N = 75,448 | **1.17 (1.04, 1.31)<br>N = 171,275** |
| | 0.72 (0.21, 1.87)<br>N = 6,452 | 1.45 (0.39, 5.37)<br>N = 1,753 | 1.26 (1.05, 1.51)<br>N = 55,564 | 1.04 (0.85, 1.27)<br>N = 73,637 | **1.15 (1.01, 1.31)<br>N = 137,406** |
| ***Maternal overweight confounder and other parent BMI adjusted*** | 0.77 (0.23, 1.99)<br>N = 6,452 | 1.46 (0.39, 5.42)<br>N = 1,753 | 1.24 (1.04, 1.49)<br>N = 55,564 | 1.05 (0.86, 1.29)<br>N = 73,637 | **1.15 (1.01, 1.31)<br>N = 137,406** |
| | 0.77 (0.23, 1.99)<br>N = 6,452 | 1.46 (0.39, 5.42)<br>N = 1,753 | 1.24 (1.04, 1.49)<br>N = 55,564 | 1.05 (0.86, 1.29)<br>N = 73,637 | **1.15 (1.01, 1.31)<br>N = 137,406** |
| ***Maternal obesity unadjusted*** | 1.99 (0.95, 3.78)<br>N = 10,970 | 1.05 (0.62, 1.74)<br>N = 10,109 | 1.30 (1.06, 1.57)<br>N = 85,350 | 1.07 (0.85, 1.35)<br>N = 97,400 | **1.21 (1.05, 1.39)<br>N = 203,829** |
| | 1.56 (0.46, 4.00)<br>N = 6,452 | 0.84 (0.12, 3.93)<br>N = 1,753 | 1.16 (0.88, 1.51)<br>N = 55,564 | 1.10 (0.83, 1.44)<br>N = 73,637 | **1.14 (0.94, 1.37)<br>N = 137,406** |
| ***Maternal obesity confounder adjusted*** | 2.16 (0.93, 4.43)<br>N = 9,179 | 1.20 (0.66, 2.11)<br>N = 7,360 | 1.21 (0.97, 1.49)<br>N = 79,288 | 1.09 (0.83, 1.43)<br>N = 75,448 | **1.19 (1.02, 1.40)<br>N = 171,275** |
| | 1.72 (0.50, 4.49)<br>N = 6,452 | 0.67 (0.10, 3.33)<br>N = 1,753 | 1.14 (0.86, 1.48)<br>N = 55,564 | 1.09 (0.83, 1.44)<br>N = 73,637 | **1.12 (0.93, 1.36)<br>N = 137,406** |
| ***Maternal obesity confounder and other parent BMI adjusted*** | 1.88 (0.55, 4.93)<br>N = 6,452 | 0.70 (0.09, 3.44)<br>N = 1,753 | 1.10 (0.83, 1.43)<br>N = 55,564 | 1.12 (0.85, 1.49)<br>N = 73,637 | **1.12 (0.93, 1.36)<br>N = 137,406** |
| | 1.88 (0.55, 4.93)<br>N = 6,452 | 0.70 (0.09, 3.44)<br>N = 1,753 | 1.10 (0.83, 1.43)<br>N = 55,564 | 1.12 (0.85, 1.49)<br>N = 73,637 | **1.12 (0.93, 1.36)<br>N = 137,406** |
| ***Paternal underweight unadjusted*** | NA | NA | 0.59 (0.10, 1.84)<br>N = 62,637 | 1.97 (0.73, 5.31)<br>N = 96,841 | **1.31 (0.58, 2.95)<br>N = 159,478** |

| | | | | | |
|---|---|---|---|---|---|
| | NA | NA | 0.38 (0.02, 1.71)<br>N = 53,922 | 0.81 (0.11, 5.80)<br>N = 67,071 | **0.56 (0.14, 2.24)<br>N = 120,993** |
| ***Paternal underweight confounder adjusted*** | NA | NA | 0.36 (0.02, 1.63)<br>N = 54,710 | 0.82 (0.11, 5.87)<br>N = 68,623 | **0.54 (0.13, 2.19)<br>N = 123,333** |
| | NA | NA | 0.37 (0.02, 1.67)<br>N = 53,922 | 0.85 (0.12, 6.09)<br>N = 67,071 | **0.56 (0.14, 2.26)<br>N = 120,993** |
| ***Paternal underweight confounder and other parent BMI adjusted*** | NA | NA | 0.36 (0.02, 1.65)<br>N = 53,922 | 0.85 (0.12, 6.08)<br>N = 67,071 | **0.55 (0.14, 2.24)<br>N = 120,993** |
| | NA | NA | 0.36 (0.02, 1.64)<br>N = 53,922 | 0.85 (0.12, 6.08)<br>N = 67,071 | **0.55 (0.14, 2.24)<br>N = 120,993** |
| ***Paternal overweight unadjusted*** | 0.90 (0.53, 1.49)<br>N – 8,076 | 0.60 (0.18, 1.88)<br>N = 2,725 | 1.10 (0.95, 1.27)<br>N = 62,637 | 1.02 (0.88, 1.18)<br>N = 96,841 | **1.05 (0.95, 1.16)<br>N = 159,478** |
| | 0.73 (0.32, 1.54)<br>N = 5,044 | 0.53 (0.11, 2.17)<br>N = 1,572 | 1.18 (1.01, 1.38)<br>N = 53,922 | 1.03 (0.86, 1.23)<br>N = 67,071 | **1.10 (0.98, 1.23)<br>N = 127,609** |
| ***Paternal overweight confounder adjusted*** | 1.07 (0.37, 3.20)<br>N = 5,550 | 0.67 (0.17, 2.39)<br>N = 2,085 | 1.20 (0.95, 1.53)<br>N = 54,710 | 1.08 (0.90, 1.28)<br>N = 68,623 | **1.11 (0.97, 1.28)<br>N = 130,968** |
| | 1.11 (0.33, 3.78)<br>N = 5,044 | 0.66 (0.13, 2.76)<br>N = 1,572 | 1.22 (0.97, 1.56)<br>N = 53,922 | 1.05 (0.88, 1.25)<br>N = 67,071 | **1.10 (0.96, 1.27)<br>N = 127,609** |
| ***Paternal overweight confounder and other parent BMI adjusted*** | 1.10 (0.33, 3.73)<br>N = 5,044 | 0.67 (0.13, 2.82)<br>N = 1,572 | 1.22 (0.96, 1.56)<br>N = 53,922 | 1.05 (0.88, 1.26)<br>N = 67,071 | **1.10 (0.96, 1.27)<br>N = 127,609** |
| | 1.10 (0.33, 3.73)<br>N = 5,044 | 0.67 (0.13, 2.82)<br>N = 1,572 | 1.22 (0.96, 1.56)<br>N = 53,922 | 1.05 (0.88, 1.26)<br>N = 67,071 | **1.10 (0.96, 1.27)<br>N = 127,609** |
| ***Paternal obesity unadjusted*** | 1.33 (0.54, 2.81)<br>N – 8,076 | 1.65 (0.56, 4.83)<br>N = 2,725 | 1.31 (1.00, 1.67)<br>N = 62,637 | 1.00 (0.79, 1.37)<br>N = 96,841 | **1.15 (0.97, 1.37)<br>N = 159,478** |
| | 1.12 (0.26, 3.31)<br>N = 5,044 | 1.40 (0.34, 5.31)<br>N = 1,572 | 1.35 (1.01, 1.76)<br>N = 53,922 | 0.95 (0.71, 1.25)<br>N = 67,071 | **1.15 (0.95, 1.40)<br>N = 127,609** |
| ***Paternal obesity confounder adjusted*** | 2.03 (0.19, 18.64)<br>N = 5,550 | 1.79 (0.50, 6.16)<br>N = 2,085 | 1.48 (0.89, 2.48)<br>N = 54,710 | 1.02 (0.76, 1.37)<br>N = 68,623 | **1.15 (0.90, 1.47)<br>N = 130,968** |
| | 2.96 (0.24, 33.50)<br>N = 5,044 | 1.93 (0.46, 7.70)<br>N = 1,572 | 1.47 (0.88, 2.49)<br>N = 53,922 | 1.02 (0.76, 1.37)<br>N = 67,071 | **1.15 (0.89, 1.48)<br>N = 127,609** |
| ***Paternal obesity confounder and other parent BMI adjusted*** | 2.99 (0.25, 33.86)<br>N = 5,044 | 1.96 (0.47, 7.78)<br>N = 1,572 | 1.46 (0.87, 2.46)<br>N = 53,922 | 1.03 (0.76, 1.39)<br>N = 67,071 | **1.16 (0.90, 1.50)<br>N = 127,609** |
| | 2.99 (0.25, 33.86)<br>N = 5,044 | 1.96 (0.47, 7.78)<br>N = 1,572 | 1.46 (0.87, 2.46)<br>N = 53,922 | 1.03 (0.76, 1.39)<br>N = 67,071 | **1.16 (0.90, 1.50)<br>N = 127,609** |

Covariates used for each study in fully adjusted models (mutually adjusted models the same as fully adjusted but with additional adjustment for the other parent's BMI and parity in paternal models);
ABCD: Maternal: offspring sex, age, education, parity, ethnicity, smoking, alcohol. Paternal: offspring sex, age, education, ethnicity.
ALSPAC: Maternal: offspring sex, age, education, parity, smoking, alcohol. Paternal: offspring sex, age, education, smoking, alcohol.
BASELINE: Maternal: offspring sex, age, education, smoking, alcohol. Paternal: offspring sex, age, smoking.
BiB: Maternal: offspring sex, age, education, parity, ethnicity, smoking. Paternal: offspring sex, age, education, ethnicity, smoking.
DNBC: Maternal: offspring sex, age, education, parity, smoking, alcohol. Paternal: offspring sex, age, education, smoking.
MoBa: Maternal: offspring sex, age, education, parity, smoking, alcohol. Paternal: offspring sex, age, education, smoking, alcohol.
NINFEA: Maternal: offspring sex, age, education, parity, smoking, alcohol. Paternal: offspring sex, age, education.

**Table S7.** Comparison **between maximal numbers (black, top rows)** and **complete case models (red, bottom rows)**. Results are odds ratios (95% CIs) of any offspring CHD for smoking during pregnancy.

| Model | *ABCD* | *ALSPAC* | *BiB* | *DNBC* | *MoBa* | *NINFEA* | *Meta-analysis results* |
|---|---|---|---|---|---|---|---|
| ***Maternal smoking unadjusted*** | 2.06 (0.77, 4.65)<br>N = 8,117 | 1.23 (0.78, 1.87)<br>N = 12,716 | 0.89 (0.53, 1.42)<br>N = 10,887 | 1.11 (0.98, 1.26)<br>N = 86,740 | 1.07 (0.86, 1.34)<br>N = 101,042 | 0.77 (0.18, 3.21)<br>N = 5,801 | **1.11 (1.00, 1.23)<br>N = 225,303** |
| | 2.04 (0.76, 4.62)<br>N = 7,824 | 1.40 (0.71, 2.56)<br>N = 7,626 | 1.62 (0.52, 4.20)<br>N = 2,624 | 1.10 (0.96, 1.26)<br>N = 78,229 | 1.03 (0.78, 1.37)<br>N = 77,266 | 0.79 (0.19, 3.29)<br>N = 5,527 | **1.11 (0.99, 1.25)<br>N = 179,096** |
| ***Maternal smoking confounder adjusted*** | 2.02 (0.73, (4.77)<br>N = 7,824 | 1.22 (0.69, 2.06)<br>N = 10,217 | 0.93 (0.50, 1.60)<br>N = 9,646 | 1.05 (0.91, 1.20)<br>N = 80,571 | 1.02 (0.77, 1.36)<br>N = 77,311 | 0.92 (0.22, 3.96)<br>N = 5,527 | **1.06 (0.94, 1.18)<br>N = 191,096** |
| | 2.02 (0.73, (4.77)<br>N = 7,824 | 1.31 (0.65, 2.46)<br>N = 7,626 | 2.09 (0.64, 5.84)<br>N = 2,624 | 1.07 (0.93, 1.23)<br>N = 78,229 | 1.02 (0.77, 1.37)<br>N = 77,266 | 0.92 (0.22, 3.96)<br>N = 5,527 | **1.09 (0.97, 1.23)<br>N = 179,096** |
| ***Maternal smoking confounder and other parent smoking adjusted*** | - | 1.27 (0.61, 2.50)<br>N = 7,626 | 1.77 (0.51, 5.36)<br>N = 2,624 | 1.11 (0.96, 1.28)<br>N = 79,000 | 1.05 (0.78, 1.41)<br>N = 77,266 | - | **1.11 (0.97, 1.25)<br>N = 166,516** |
| | - | 1.27 (0.61, 2.50)<br>N = 7,626 | 1.77 (0.51, 5.36)<br>N = 2,624 | 1.13 (0.98, 1.30)<br>N = 78,229 | 1.05 (0.78, 1.41)<br>N = 77,266 | - | **1.12 (0.99, 1.28)<br>N = 165,745** |
| ***Paternal smoking unadjusted*** | - | 1.29 (0.79, 2.10)<br>N = 9,134 | 1.20 (0.50, 2.66)<br>N = 3,187 | 0.95 (0.84, 1.08)<br>N = 84,926 | 0.96 (0.82, 1.11)<br>N = 101,804 | - | **0.97 (0.88, 1.06)<br>N = 198,421** |
| | - | 1.28 (0.68, 2.35)<br>N = 6,182 | 1.46 (0.54, 3.73)<br>N = 2,373 | 0.96 (0.84, 1.10)<br>N = 77,477 | 1.00 (0.83, 1.20)<br>N = 70,018 | - | **0.99 (0.89, 1.10)<br>N = 156,050** |
| ***Paternal smoking confounder adjusted*** | - | 1.17 (0.61, 2.19)<br>N = 6,308 | 1.43 (0.51, 3.76)<br>N = 2,424 | 0.95 (0.83, 1.08)<br>N = 77,526 | 1.05 (0.87, 1.26)<br>N = 70,766 | - | **0.99 (0.89, 1.10)<br>N = 157,024** |
| | - | 1.23 (0.64, 2.30)<br>N = 6,182 | 1.51 (0.53, 4.06)<br>N = 2,373 | 0.95 (0.83, 1.08)<br>N = 77,477 | 1.05 (0.87, 1.27)<br>N = 70,018 | - | **0.99 (0.89, 1.10)<br>N = 156,050** |
| ***Paternal smoking confounder and other parent BMI adjusted*** | - | 1.14 (0.56, 2.23)<br>N = 6,182 | 1.18 (0.38, 3.41)<br>N = 2,373 | 0.90 (0.79, 1.04)<br>N = 77,499 | 1.04 (0.85, 1.26)<br>N = 70,018 | - | **0.96 (0.85, 1.07)<br>N = 156,072** |
| | - | 1.14 (0.64, 2.30)<br>N = 6,182 | 1.18 (0.38, 3.41)<br>N = 2,373 | 0.91 (0.79, 1.04)<br>N = 77,477 | 1.04 (0.85, 1.26)<br>N = 70,018 | - | **0.96 (0.86, 1.07)<br>N = 156,050** |

Covariates used for each study in fully adjusted models (mutually adjusted models the same as fully adjusted but with additional adjustment for the other parent's smoking);
ABCD: Maternal: offspring sex, age, education, parity, ethnicity, alcohol.
ALSPAC: Maternal: offspring sex, age, education, parity, alcohol. Paternal: offspring sex, age, education, alcohol.
BiB: Maternal: offspring sex, age, education, parity, ethnicity. Paternal: offspring sex, age, education, ethnicity.
DNBC: Maternal: offspring sex, age, education, parity, alcohol. Paternal: offspring sex, age, education.
MoBa: Maternal: offspring sex, age, education, parity, alcohol. Paternal: offspring sex, age, education, alcohol.
NINFEA: Maternal: offspring sex, age, education, parity, alcohol.

**Table S8.** Comparison between **maximal numbers (black, top rows)** and **complete case models (red, bottom rows)**. Results are odds ratios (95% CIs) of any offspring CHD for alcohol intake during pregnancy in comparison to non-drinkers.

| Model | *ABCD* | *ALSPAC* | *DNBC* | *MoBa* | *NINFEA* | *Meta-analysis results* |
|---|---|---|---|---|---|---|
| ***Maternal alcohol (yes/no) unadjusted*** | 1.38 (0.61, 2.85)<br>N = 8,125 | 1.20 (0.81, 1.80)<br>N = 12,622 | 1.00 (0.89, 1.12)<br>N = 86,708 | 1.04 (0.88, 1.23)<br>N = 82,358 | 1.20 (0.57, 2.51)<br>N = 5,843 | **1.03 (0.94, 1.12)<br>N = 195,656** |
| | 1.36 (0.60, 2.81)<br>N = 7,824 | 1.18 (0.56, 2.55)<br>N = 4,585 | 1.00 (0.89, 1.13)<br>N = 79,648 | 1.06 (0.86, 1.31)<br>N = 51,006 | 1.19 (0.57, 2.49)<br>N = 5,527 | **1.03 (0.93, 1.14)<br>N = 148,590** |
| ***Maternal alcohol (yes/no) confounder adjusted*** | 1.17 (0.50, 2.56)<br>N = 7,824 | 1.24 (0.78, 2.01)<br>N = 10,217 | 1.01 (0.90, 1.14)<br>N = 80,571 | 1.03 (0.86, 1.23)<br>N = 77,311 | 1.18 (0.56, 2.49)<br>N = 5,527 | **1.03 (0.94, 1.13)<br>N = 181,450** |
| | 1.17 (0.50, 2.56)<br>N = 7,824 | 1.20 (0.56, 2.63)<br>N = 4,585 | 1.01 (0.89, 1.14)<br>N = 79,648 | 1.06 (0.85, 1.31)<br>N = 51,066 | 1.18 (0.56, 2.49)<br>N = 5,527 | **1.03 (0.93, 1.14)<br>N = 148,590** |
| ***Maternal light drinking unadjusted*** | - | 0.93 (0.52, 1.67)<br>N = 6,501 | 0.92 (0.82, 1.03)<br>N = 88,349 | 1.10 (0.88, 1.36)<br>N = 84,436 | - | **0.96 (0.87, 1.06)<br>N = 179,286** |
| | - | 1.27 (0.58, 2.93)<br>N = 4,585 | 0.93 (0.82, 1.05)<br>N = 79,648 | 1.24 (0.94, 1.63)<br>N = 51,006 | - | **0.98 (0.88, 1.09)<br>N = 135,239** |
| ***Maternal light drinking confounder adjusted*** | - | 0.92 (0.48, 1.78)<br>N = 5,797 | 0.95 (0.85, 1.08)<br>N = 80,214 | 1.13 (0.90, 1.41)<br>N = 79,695 | - | **0.99 (0.89, 1.10)<br>N = 165,706** |
| | - | 1.35 (0.61, 3.14)<br>N = 4,585 | 0.94 (0.83, 1.06)<br>N = 79,648 | 1.22 (0.92, 1.61)<br>N = 51,006 | - | **0.99 (0.88, 1.10)<br>N = 135,239** |
| ***Maternal light drinking confounder and other parent alcohol adjusted*** | - | 1.40 (0.62, 3.27)<br>N = 4,585 | - | 1.13 (0.87, 1.47)<br>N = 59,571 | - | **1.15 (0.90, 1.48)<br>N = 64,156** |
| | - | 1.40 (0.62, 3.27)<br>N = 4,585 | - | 1.19 (0.90, 1.57)<br>N = 51,006 | - | **1.21 (0.93, 1.57)<br>N = 55,591** |
| ***Maternal mod/heavy drinking unadjusted*** | - | 0.67 (0.22, 1.65)<br>N = 6,501 | 1.14 (0.87, 1.48)<br>N = 88,349 | 1.85 (0.92, 3.73)<br>N = 84,436 | - | **1.17 (0.92, 1.49)<br>N = 179,286** |
| | - | 0.92 (0.21, 3.01)<br>N = 4,585 | 1.19 (0.89, 1.56)<br>N = 79,648 | 1.77 (0.66, 4.78)<br>N = 51,006 | - | **1.21 (0.93, 1.58)<br>N = 135,239** |
| ***Maternal mod/heavy drinking confounder adjusted*** | - | 0.64 (0.18, 1.75)<br>N = 5,797 | 1.21 (0.90, 1.58)<br>N = 80,214 | 1.47 (0.65, 3.32)<br>N = 79,695 | - | **1.19 (0.92, 1.54)<br>N = 165,706** |
| | - | 0.89 (0.20, 2.98)<br>N = 4,585 | 1.19 (0.89, 1.57)<br>N = 79,648 | 1.73 (0.64, 4.69)<br>N = 51,006 | - | **1.21 (0.93, 1.58)<br>N = 135,239** |
| ***Maternal mod/heavy drinking confounder and other parent alcohol adjusted*** | - | 0.94 (2.06, 3.19)<br>N = 4,585 | - | 1.31 (0.48, 3.56)<br>N = 59,571 | - | **1.16 (0.52, 2.58)<br>N = 64,156** |
| | - | 0.94 (2.06, 3.19)<br>N = 4,585 | - | 1.57 (0.58, 4.27)<br>N = 51,006 | - | **1.30 (0.59, 2.89)<br>N = 55,591** |
| ***Paternal light drinking unadjusted*** | - | 0.90 (0.36, 3.02)<br>N = 8,205 | - | 0.90 (0.61, 1.32)<br>N = 72,422 | - | **0.90 (0.63, 1.29)<br>N = 80,627** |
| | - | 1.90 (0.39, 34.09)<br>N = 5,228 | - | 1.01 (0.62, 1.65)<br>N = 58,847 | - | **1.05 (0.65, 1.68)<br>N = 64,075** |
| ***Paternal light drinking confounder adjusted*** | - | 2.11 (0.44, 37.99)<br>N = 5,346 | - | 0.86 (0.58, 1.28)<br>N = 70,766 | - | **0.89 (0.60, 1.31)<br>N = 76,112** |
| | - | 2.04 (0.42, 36.80)<br>N = 5,228 | - | 0.97 (0.60, 1.58)<br>N = 58,847 | - | **1.01 (0.63, 1.63)<br>N = 64,075** |

| | | | | | | |
|---|---|---|---|---|---|---|
| ***Paternal light drinking confounder and other parent alcohol adjusted*** | | 1.77 (0.36, 32.20)<br>N = 5,316 | - | 0.97 (0.63, 1.62)<br>N = 58,847 | | **1.01 (0.63, 1.62)<br>N = 64,163** |
| | - | 1.74 (0.35, 31.60)<br>N = 5,228 | - | 0.97 (0.60, 1.59)<br>N = 58,847 | - | **1.01 (0.62, 1.62)<br>N = 64,075** |
| ***Paternal mod/heavy drinking unadjusted*** | - | 0.86 (0.34, 2.93)<br>N = 8,205 | - | 1.11 (0.73, 1.70)<br>N = 72,422 | - | **1.08 (0.73, 1.59)<br>N = 80,627** |
| | - | 1.83 (0.37, 33.05)<br>N = 5,228 | - | 1.28 (0.76, 2,17)<br>N = 58,847 | - | **1.31 (0.79, 2.18)<br>N = 64,075** |
| ***Paternal mod/heavy drinking confounder adjusted*** | - | 2.00 (0.40, 36.05)<br>N = 5,346 | - | 1.07 (0.69, 1.66)<br>N = 70,766 | - | **1.10 (0.72, 1.69)<br>N = 76,112** |
| | - | 1.94 (0.39, 35.05)<br>N = 5,228 | - | 1.20 (0.71, 2.04)<br>N = 58,847 | - | **1.24 (0.74, 2.07)<br>N = 64,075** |
| ***Paternal mod/heavy drinking confounder and other parent alcohol adjusted*** | - | 1.72 (0.34, 31.20)<br>N = 5,316 | - | 1.21 (0.71, 2.05)<br>N = 58,847 | - | **1.23 (0.74, 2.06)<br>N = 64,163** |
| | - | 1.70 (0.34, 30.83)<br>N = 5,228 | - | 1.21 (0.71, 2.05)<br>N = 58,847 | - | **1.23 (0.74, 2.06)<br>N = 64,075** |

Covariates used for each study in fully adjusted models (mutually adjusted models the same as fully adjusted but with additional adjustment for the other parent's alcohol intake);
ABCD: Maternal: offspring sex, age, education, parity, ethnicity, smoking.
ALSPAC: Maternal: offspring sex, age, education, parity, smoking. Paternal: offspring sex, age, education, smoking.
DNBC: Maternal: offspring sex, age, education, parity, smoking.
MoBa: Maternal: offspring sex, age, education, parity, smoking. Paternal: offspring sex, age, education, smoking.
NINFEA: Maternal: offspring sex, age, education, parity, ethnicity, smoking.

#### ***Supplementary results (BMI)***

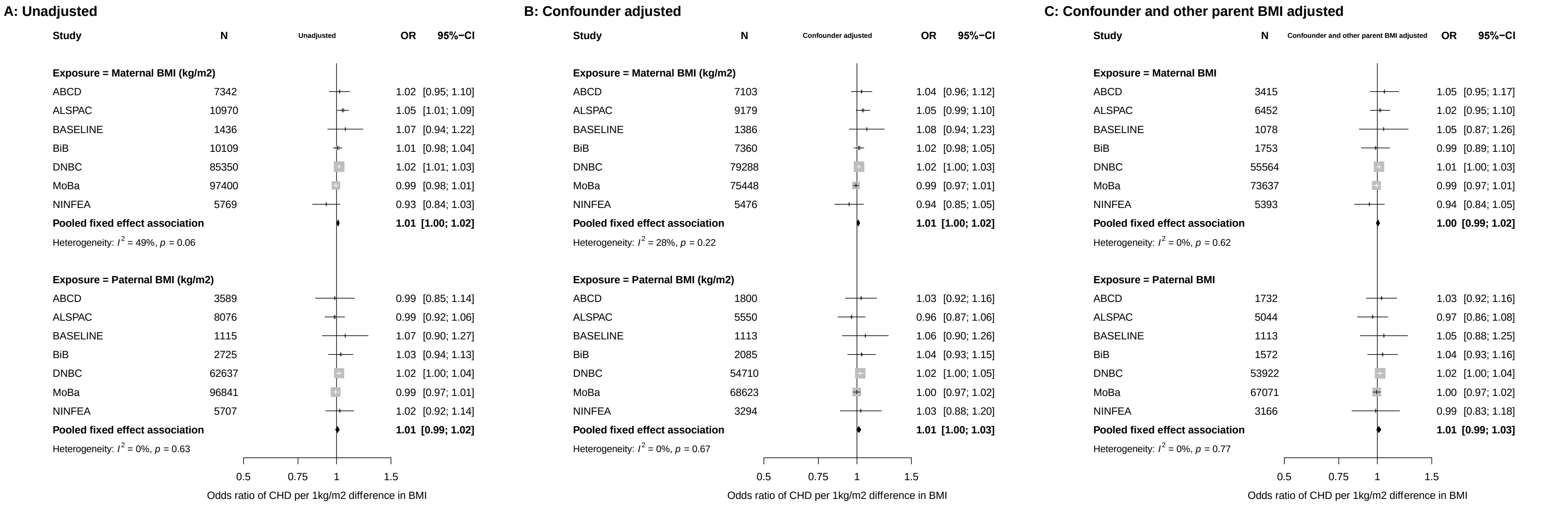

**Figure S8.** Main analysis associations between parental BMI as a continuous measurement in kg/m$^2$ (maternal top, paternal bottom) and offspring congenital heart disease. Panel A results are unadjusted, panel B results are fully adjusted for all confounders and panel C results are adjusted for all confounders as well as other parent's BMI. Confounders: **ABCD:** parental age, education, parity, ethnicity, smoking, alcohol, offspring sex; ***ALSPAC:*** parental age, education, parity, smoking, alcohol, offspring sex; ***BASELINE:*** parental age, education, smoking, alcohol, offspring sex ***BiB***: parental age, education, parity, ethnicity, smoking, offspring sex; ***DNBC:***, parental age, education, parity, smoking, alcohol, offspring sex; **MoBa:** parental age, education, parity, smoking, alcohol, offspring sex; **NINFEA:** parental age, education, parity, smoking, alcohol, offspring sex.

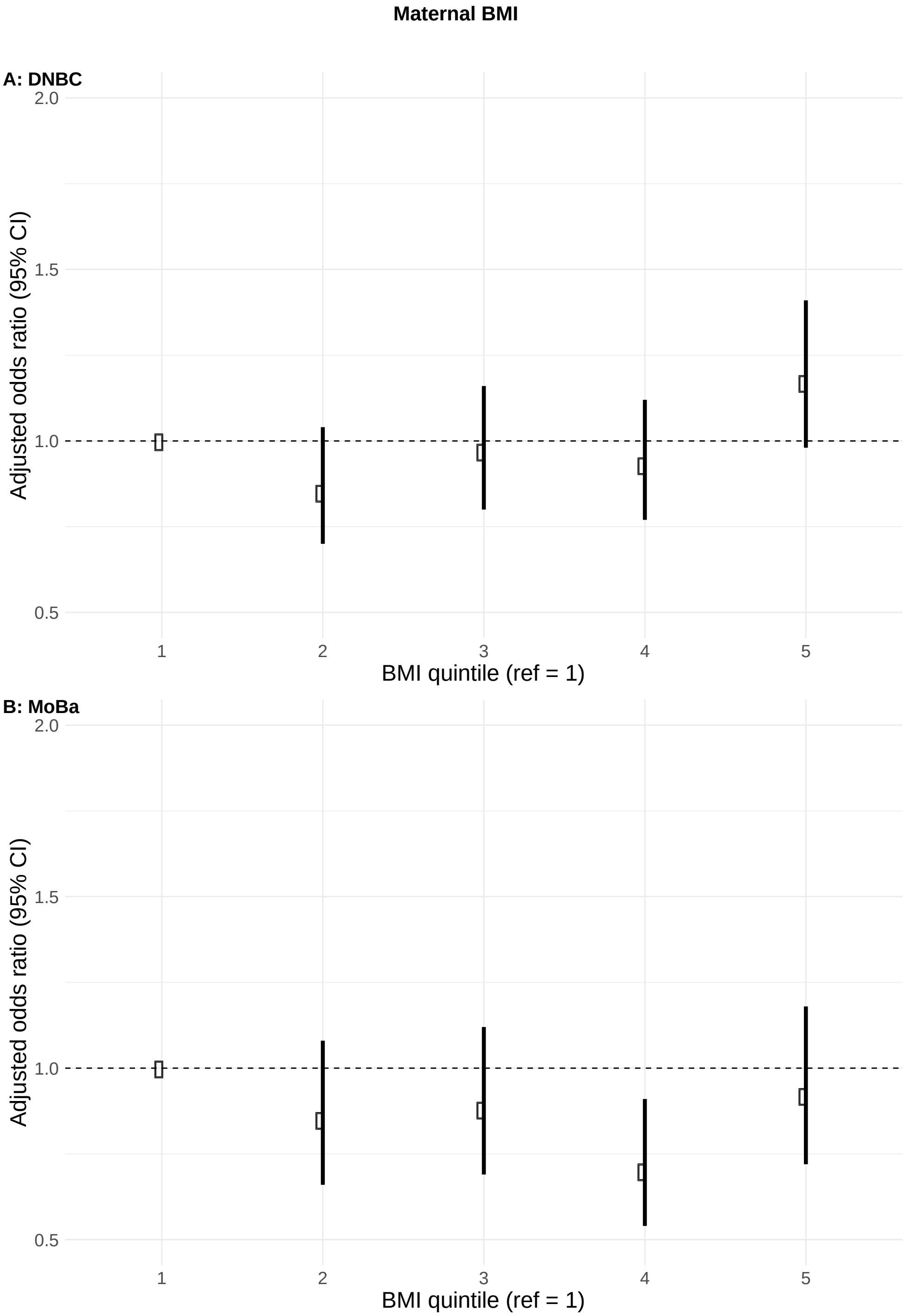

**Figure S9.** Confounder adjusted associations between maternal BMI split into fifths and offspring CHDs in the DNBC (A) and MoBa (B). Results are odds ratios and 95% CIs for maternal BMI quintile and offspring CHD in comparison to BMI quintile 1.

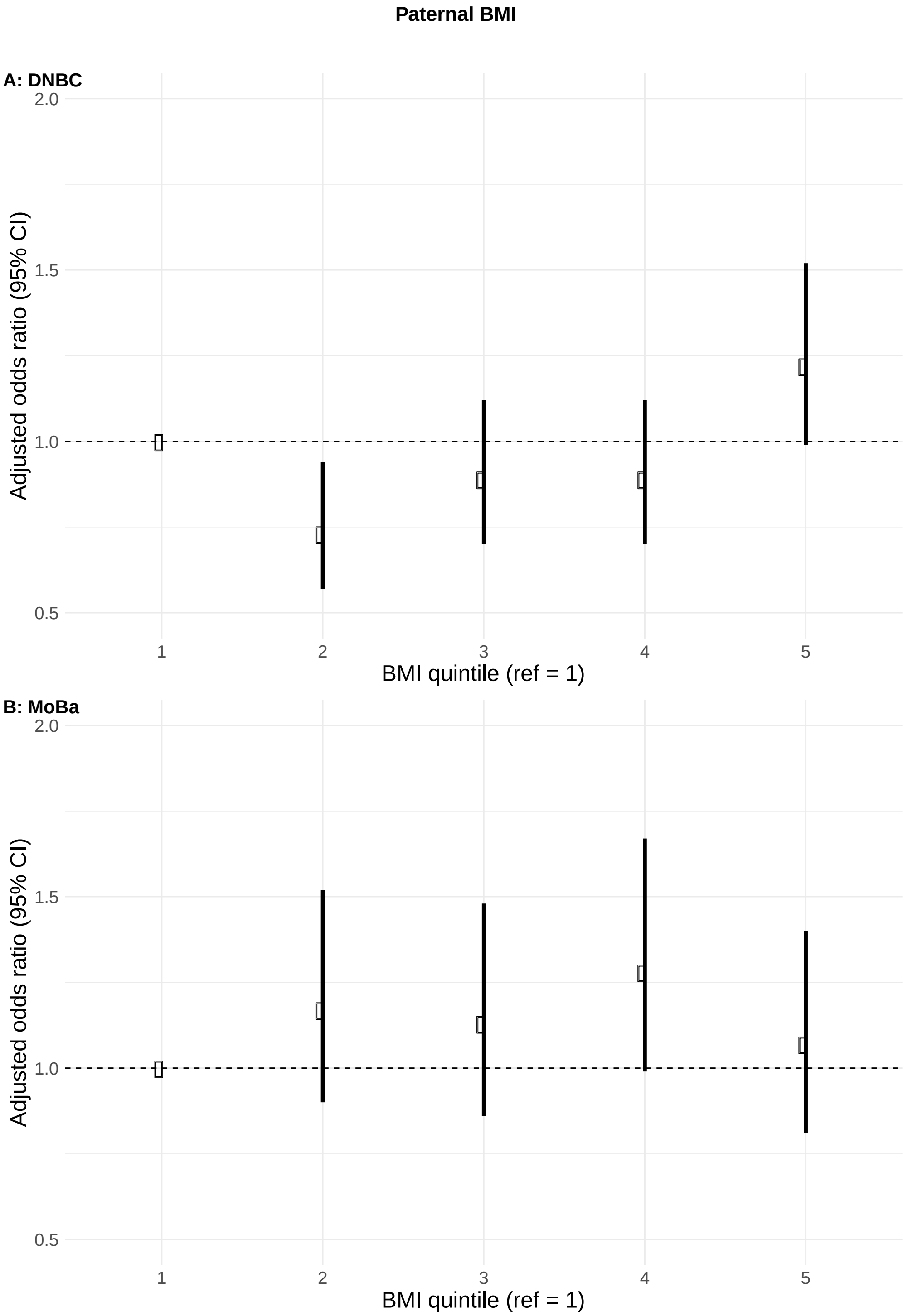

**Figure S10.** Confounder adjusted associations between paternal BMI split into fifths and offspring CHDs in the DNBC (A) and MoBa (B). Results are odds ratios and 95% CIs for paternal BMI quintile and offspring CHD in comparison to BMI quintile 1.

***BMI analyses using World Health Organization (WHO) categories***

In this section, we present meta-analysis results from the WHO BMI analyses. All Odds ratios should be interpreted as an increase/decrease odds of CHD for a maternal/paternal BMI category in comparison to normal weight. The BMI categories are: underweight (BMI <18.5 kg/m$^2$), normal weight (BMI 18.5 to <25 kg/m$^2$, reference range), overweight (BMI 25 to <30 kg/m$^2$) and obesity (BMI ≥30 kg/m$^2$).

We present fully adjusted (adjusted for all confounders) and mutually adjusted (adjusted for all confounders plus other parents' exposure) models. ALSPAC, BiB, DNBC and MoBa contributed to these analyses. Covariates adjusted for by each study are:

- ALSPAC - parental age, education, parity, smoking, alcohol, offspring sex
- BiB - parental age, education, parity, ethnicity, smoking, offspring sex
- DNBC – parental age, education, parity, smoking, alcohol, offspring sex
- MoBa - parental age, education, parity, smoking, alcohol, offspring sex

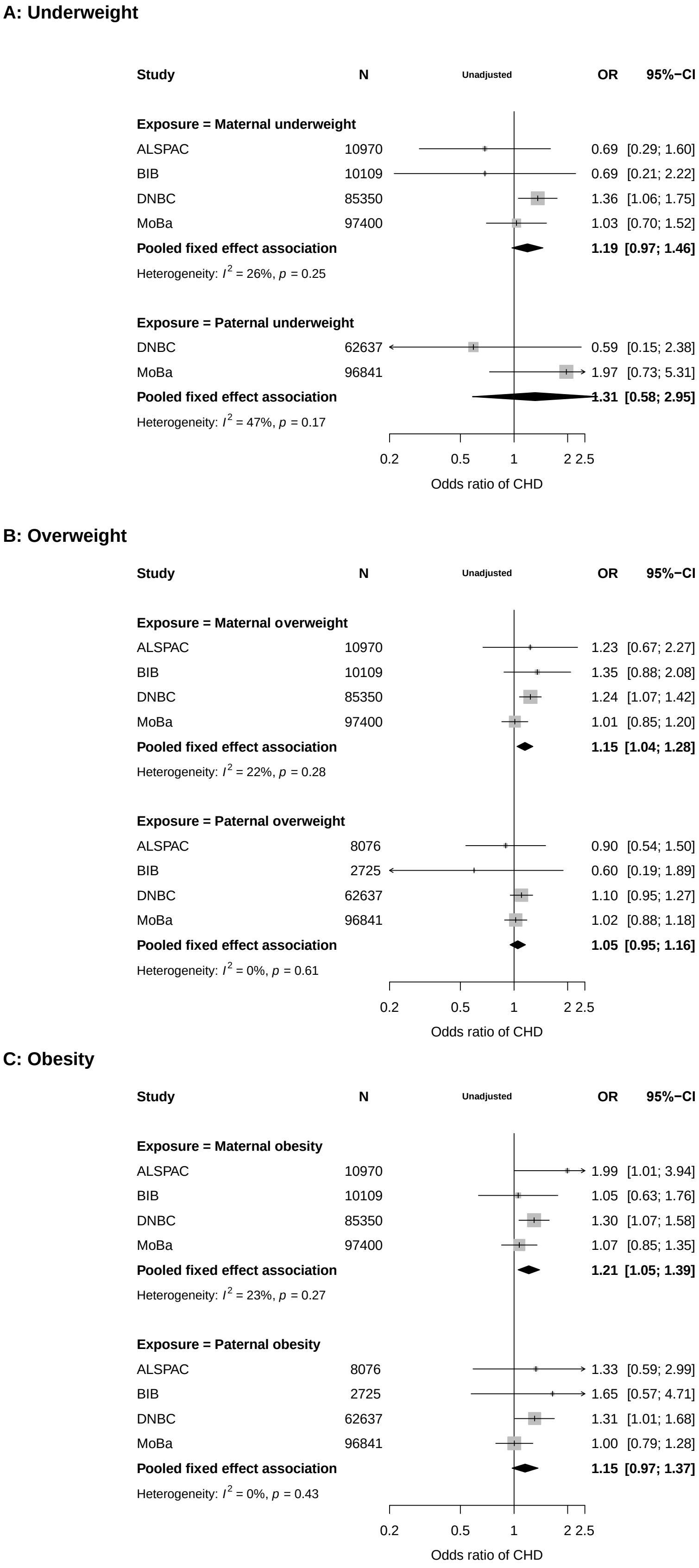

**Figure S11.** Meta-analysis results for unadjusted BMI categories using World Health Organization cut-offs with normal BMI as the reference. Outcome = any CHD in the offspring.

**A: Underweight**

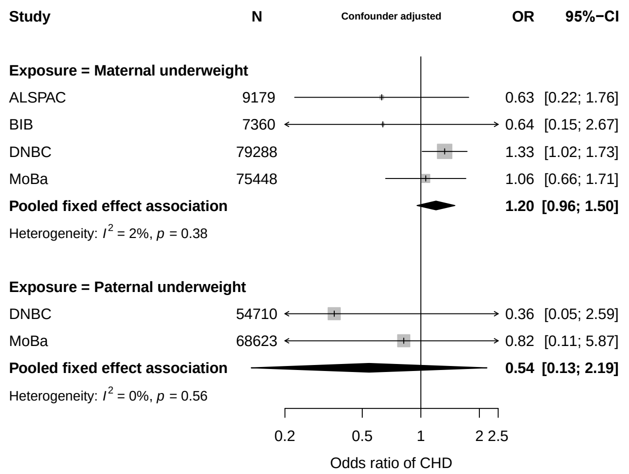

**B: Overweight**

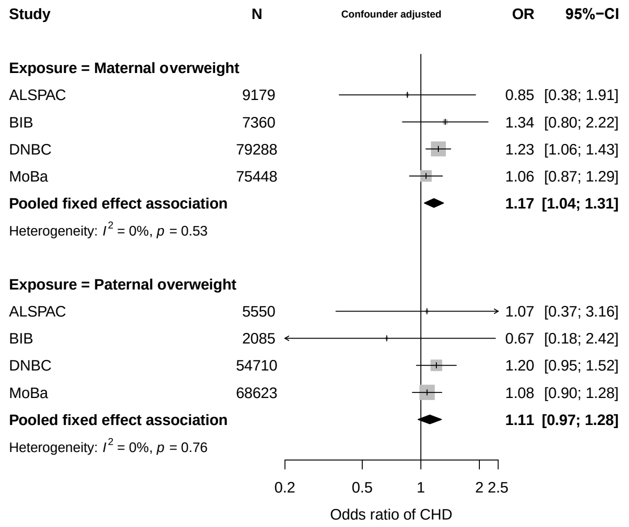

**C: Obesity**

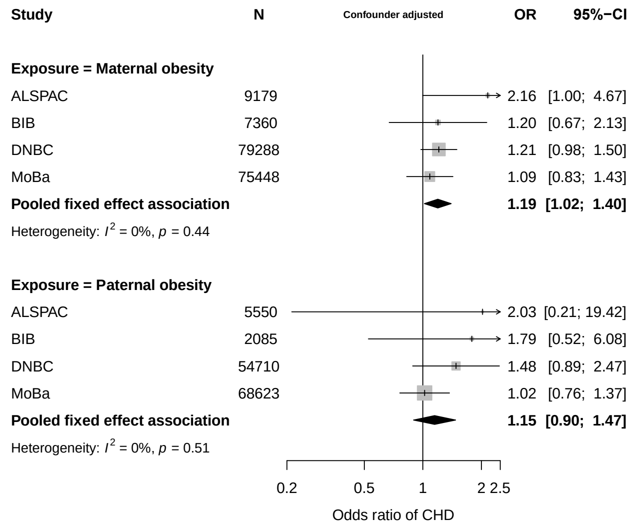

**Figure S12.** Meta-analysis results for confounder adjusted BMI categories using World Health Organization cut-offs with normal BMI as the reference. Outcome = any CHD in the offspring.

**A: Underweight**

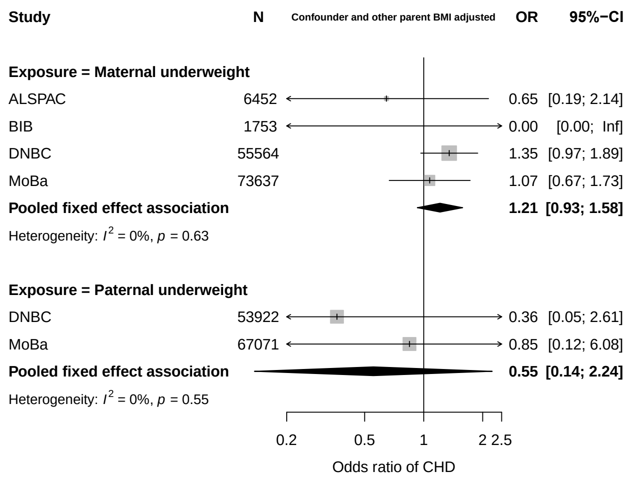

**B: Overweight**

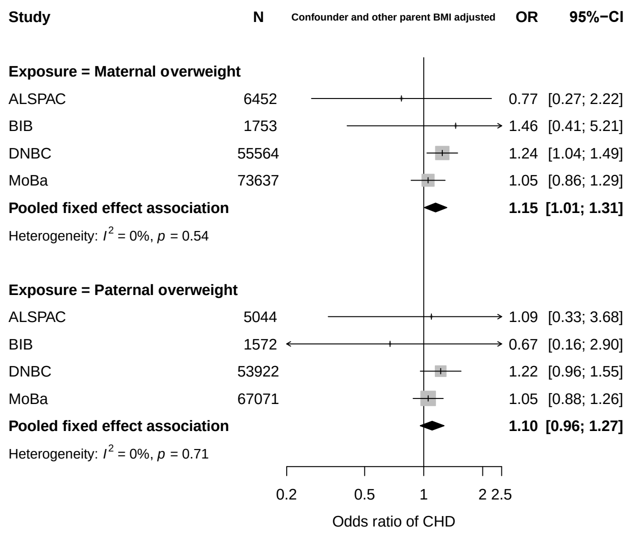

**C: Obesity**

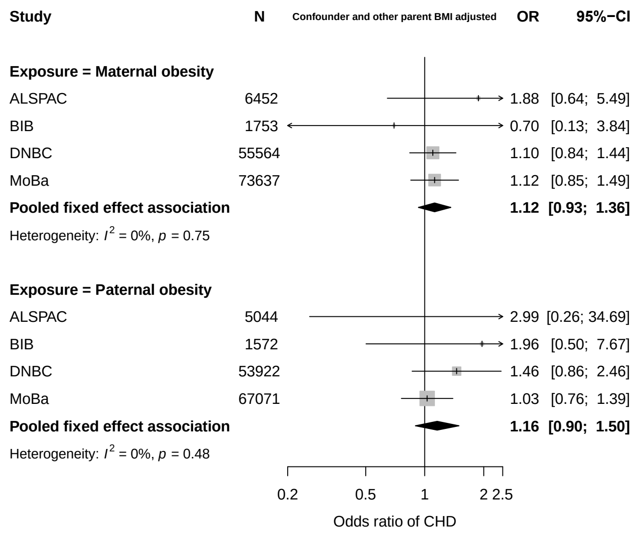

**Figure S13.** Meta-analysis results for confounder and other parent BMI adjusted BMI categories using World Health Organization cut-offs with normal BMI as the reference. Outcome = any CHD in the offspring.

***BMI supplementary results additional and CHD severity analyses***

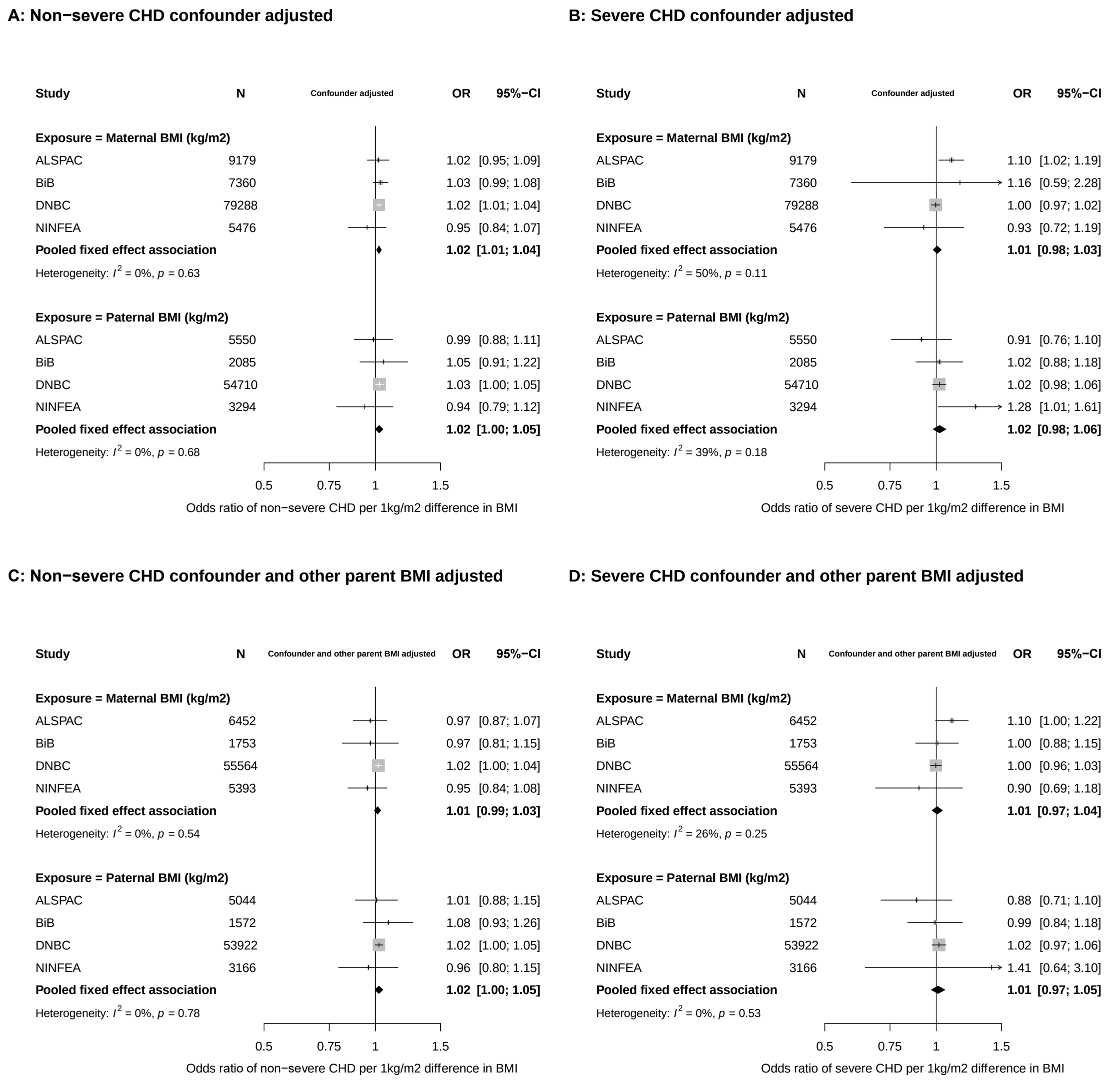

**Figure S14.** Linear associations (**top (A&B): confounder adjusted, bottom (C&D): confounder and other parent BMI adjusted)** between parental BMI and offspring non-severe congenital heart disease **(left)** and severe congenital heart disease **(right)**. Definitions for CHD subtypes can be found in Table S2.

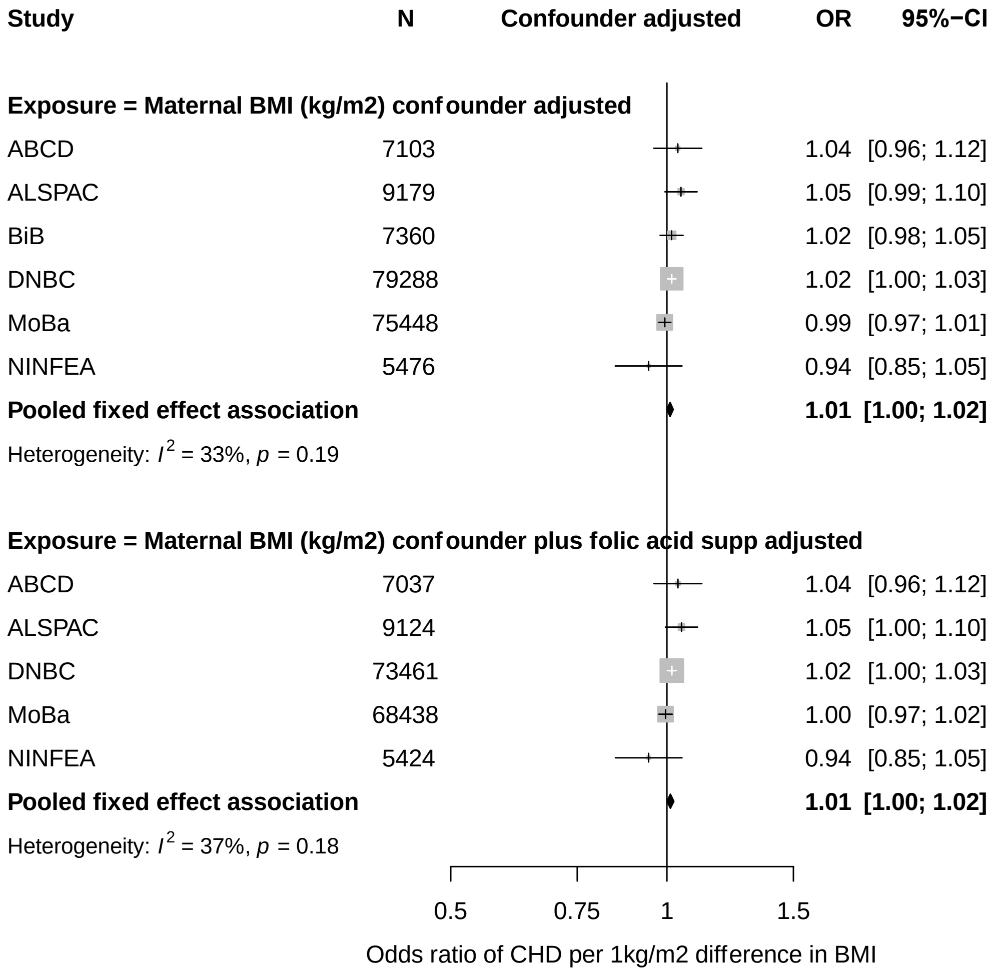

**Figure S15.** Linear associations between maternal BMI and offspring congenital heart disease. Results are fully adjusted for all confounders (top) and all confounders plus additional adjustment for folic acid supplementation during weeks 0-12 of pregnancy (bottom).

**Mean BMI results with chromosomal/genetic defects removed from study population**

**A: Additional analysis confounder adjusted**

**B: Additional analysis confounder and other parent BMI adjusted**

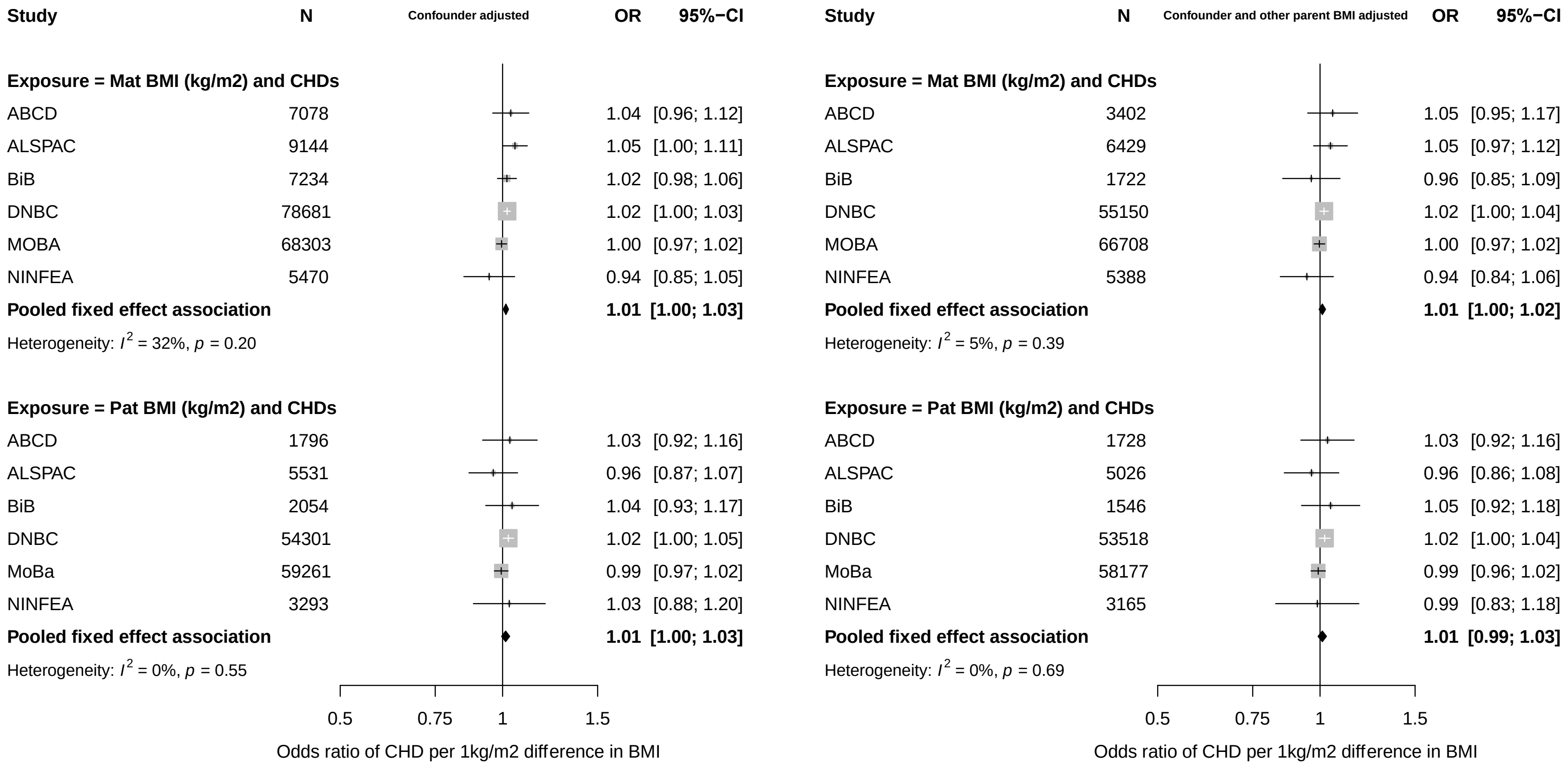

**Figure S16.** Additional analysis: linear associations between parental BMI and offspring congenital heart disease with chromosomal/genetic defects removed from the study population. **A** is adjusted for all confounders, and **B** is adjusted for all confounders and the other parent's BMI. The rationale here is to see if estimates differ when we remove offspring from the population with an anomaly associated with a pre-specified cause such as a genetic, chromosomal or teratogenic aberration. ICD codes used to remove these cases from the population can be found in Table S3. For comparison the pooled associations from main analyses (without removal of genetic/chromo disorders) were: 1.01 (1.00, 1.02) & 1.01 (0.99, 1.02) for maternal (top graphs, left and right respectively) and 1.01 (1.00, 1.03) & 1.01 (0.99, 1.03) for paternal (bottom graphs left and right respectively).

**Table S9.** Meta-analysis results from 4 cohorts (ALSPAC, BiB, DNBC, MoBa) for associations between BMI categories and CHDs. Results reported as odds ratios for CHD/CA/chromosomal for parental underweight, overweight or obesity in comparison to parental normal weight.

| ***Model*** | ***Main analysis*** <br> ***Outcome = CHD*** | ***Additional analysis*** <br> ***Outcome = CHD with chromo/gen defects removed from study population*** |
|---|---|---|
| ***Confounder adjusted*** | **M**-Underweight: 1.20 (0.96, 1.50) <br> **P**-Underweight: 0.54 (0.13, 2.19) | **M**- Underweight: 1.16 (0.90, 1.48) <br> **P**-Underweight: 0.67 (0.16, 2.70) |
| | **M**-Overweight: 1.17 (1.04, 1.31) <br> **P**-Overweight: 1.11 (0.97, 1.28) | **M**-Overweight: 1.20 (1.06, 1.35) <br> **P**-Overweight: 1.09 (0.94, 1.27) |
| | **M**-Obesity: 1.19 (1.02, 1.40) <br> **P**-Obesity: 1.15 (0.90, 1.47) | **M**-Obesity: 1.21 (1.02, 1.44) <br> **P**-Obesity: 1.19 (0.91, 1.58) |
| ***Confounder and other parent BMI adjusted*** | **M**-Underweight: 1.21 (0.93, 1.58) <br> **P**-Underweight: 0.55 (0.14, 2.24) | **M**-Underweight: 1.22 (0.90, 1.63) <br> **P**-Underweight: 0.67 (0.17, 2.72) |
| | **M**-Overweight: 1.15 (1.01, 1.31) <br> **P**-Overweight: 1.10 (0.96, 1.27) | **M**-Overweight: 1.20 (1.04, 1.38) <br> **P**-Overweight: 1.08 (0.93, 1.27) |
| | **M**-Obesity: 1.12 (0.93, 1.36) <br> **P**-Obesity: 1.16 (0.90, 1.50) | **M**-Obesity: 1.15 (0.93, 1.42) <br> **P**-Obesity: 1.20 (0.90, 1.59) |

**M** = maternal
**P** = paternal

### ***Supplementary results (Smoking)***

##### ***Smoking supplementary results from main analyses and additional analyses***

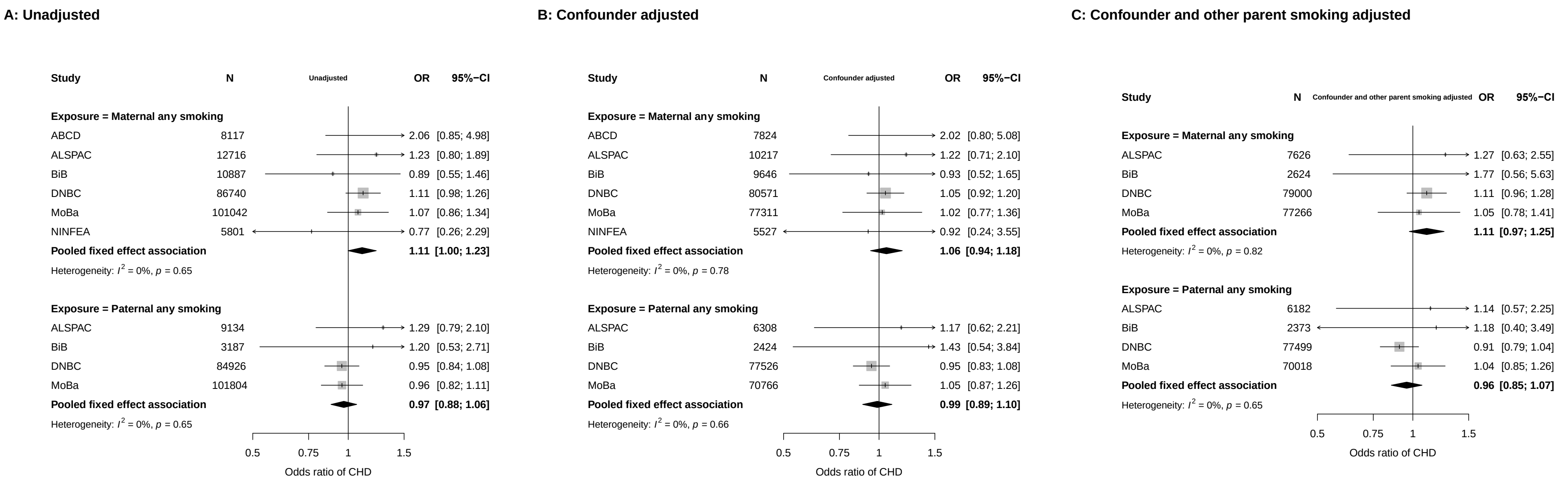

**Figure S17.** Main analysis associations between parental smoking (maternal top, paternal bottom) and offspring congenital heart disease. Panel A results are unadjusted, B results are fully adjusted for all confounders and C results are adjusted for all confounders as well as other parent's smoking. Confounders: **ABCD:** parental age, education, parity, ethnicity, alcohol, offspring sex; ***ALSPAC:*** parental age, education, parity, alcohol, offspring sex; ***BiB***: parental age, education, parity, ethnicity, offspring sex; ***DNBC:***, parental age, education, parity, alcohol, offspring sex; **MoBa:** parental age, education, parity, alcohol, offspring sex; **NINFEA:** parental age, education, parity, alcohol, offspring sex.

**Smoking (yes/no) results with chromosomal/genetic defects removed from study population**

**A: Additional analysis confounder adjusted**

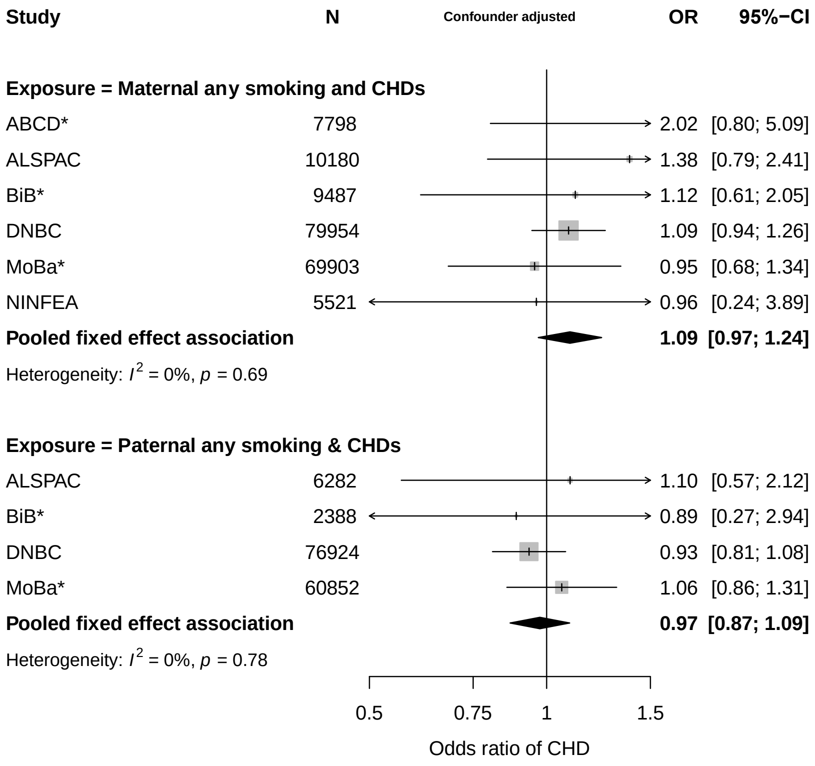

**B: Additional analysis confounder and other parent smoking adjusted**

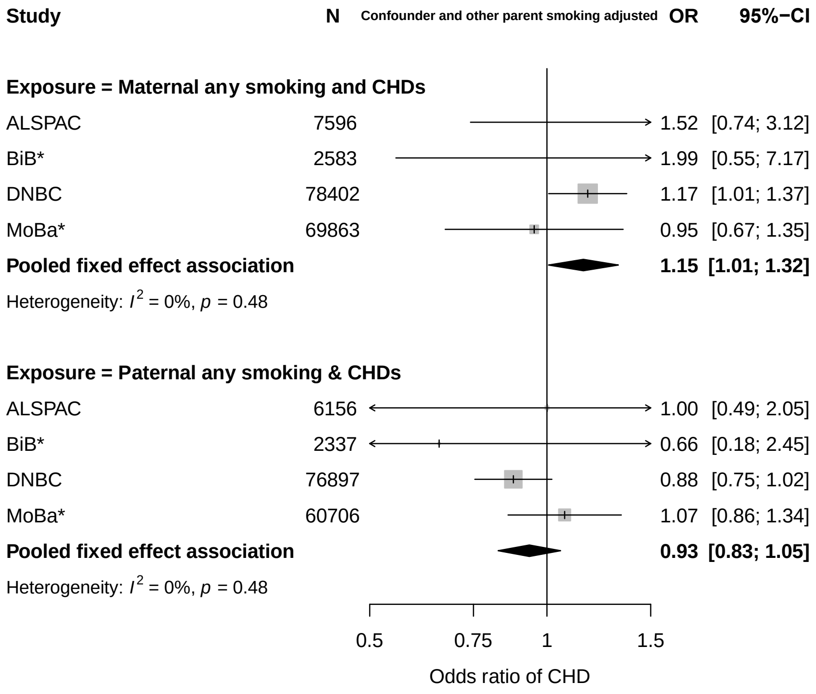

**Figure S18.** Additional analysis: Associations between parental smoking and offspring congenital heart disease with chromosomal/genetic defects removed from the study population. **A** is adjusted for all confounders, and **B** is adjusted for all confounders and the other parent's smoking. The rationale here is to see if estimates differ when we remove offspring from the population with an anomaly associated with a pre-specified cause such as a genetic, chromosomal or teratogenic aberration. ICD codes used to remove these cases from the population can be found in Table S3. For comparison the pooled associations from main analyses (without removal of genetic/chromo disorders) were: 1.06 (0.94, 1.18) & 1.11 (0.97, 1.25) for maternal (top graphs, left and right respectively) and 0.99 (0.89, 1.0) & 0.96 (0.85, 1.07) for paternal (bottom graphs left and right respectively).

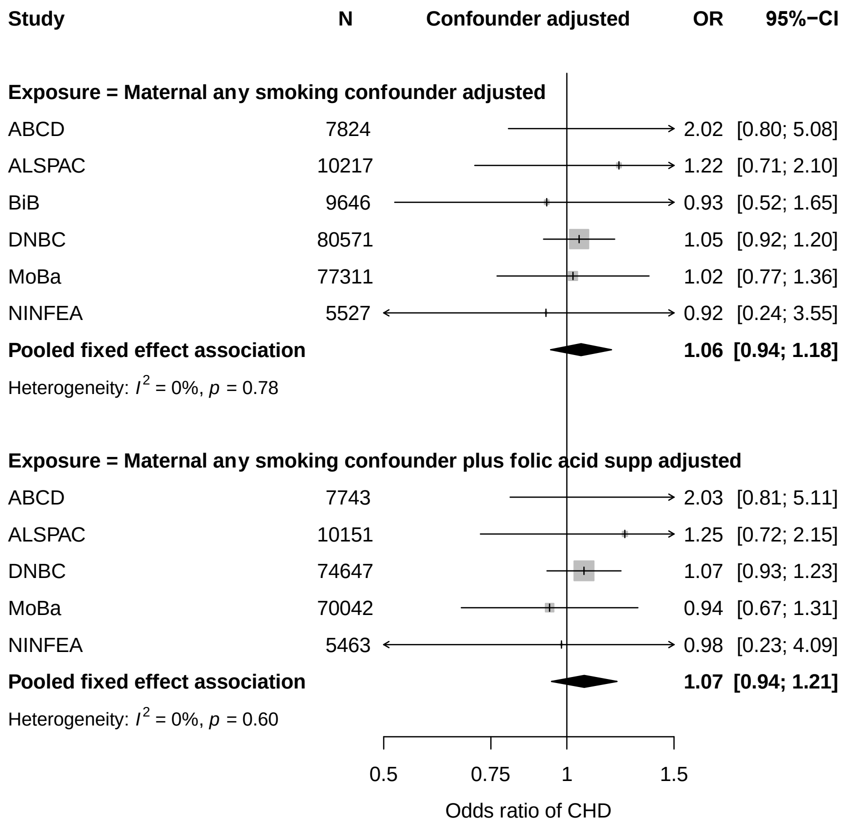

**Figure S19.** Associations between maternal smoking and offspring congenital heart disease. Results are fully adjusted for all confounders (top) and all confounders plus additional adjustment for folic acid supplementation during weeks 0-12 of pregnancy (bottom).

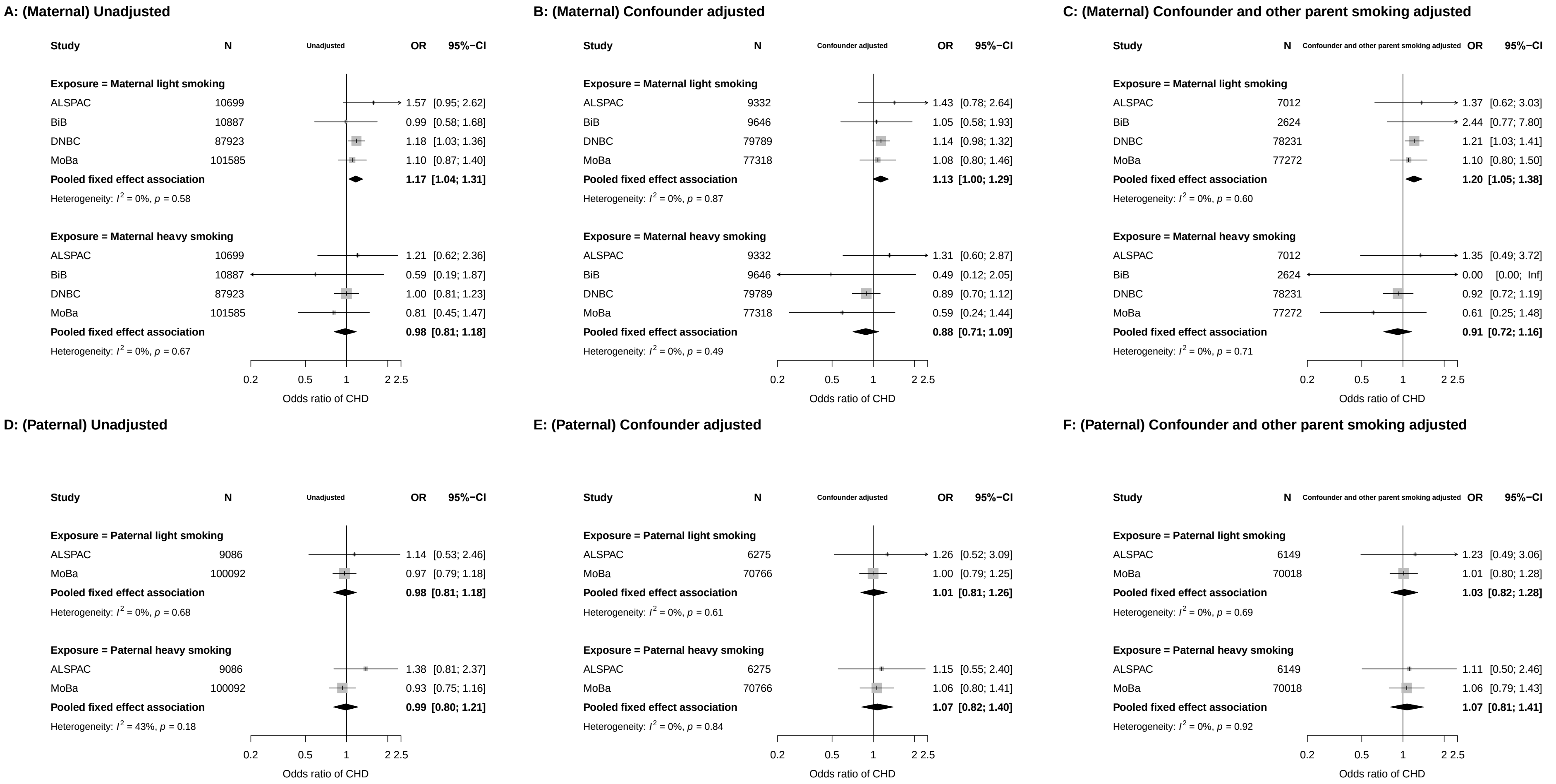

**Figure S20.** Associations between parental smoking heaviness **(top (A, B & C): maternal, bottom (D, E & F): paternal)** and offspring congenital heart disease. Results are unadjusted (left), adjusted for all confounders (middle) as well as all confounders and other parents smoking (right). Smoking categorized as none (non-smoker), light (< 10 cigarettes smoked per day during pregnancy) and heavy (≥ 10 cigarettes per day). Results presented as odds ratios and 95% confidence intervals for offspring CHD in comparison to non-smokers.

#### ***Supplementary results (Alcohol)***

#### A: Unadjusted

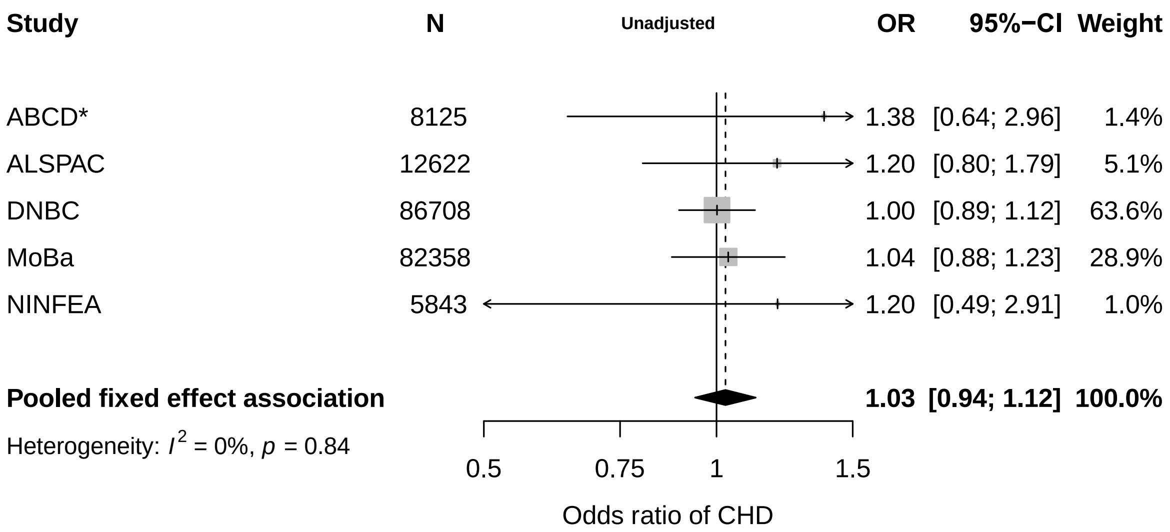

#### B: Confounder adjusted

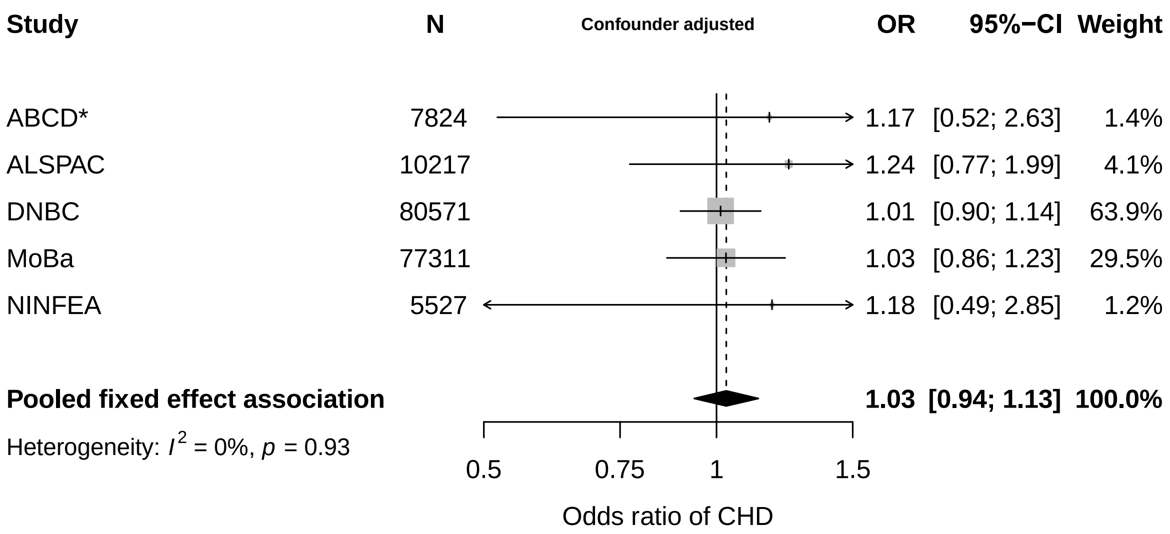

**Figure S21.** Associations between maternal alcohol consumption in the first trimester and offspring congenital heart disease. ABCD did not have trimester-specific data, therefore analyses presented for ABCD are any alcohol consumption during pregnancy. Results are adjusted for all confounders. Confounders: **ABCD:** parental age, education, parity, ethnicity, smoking, offspring sex; ***ALSPAC:*** parental age, education, parity, smoking, offspring sex; ***DNBC:*** parental age, education, parity, smoking, offspring sex; **MoBa:** parental age, education, parity, smoking, offspring sex; **NINFEA:** parental age, education, parity, smoking, offspring sex.

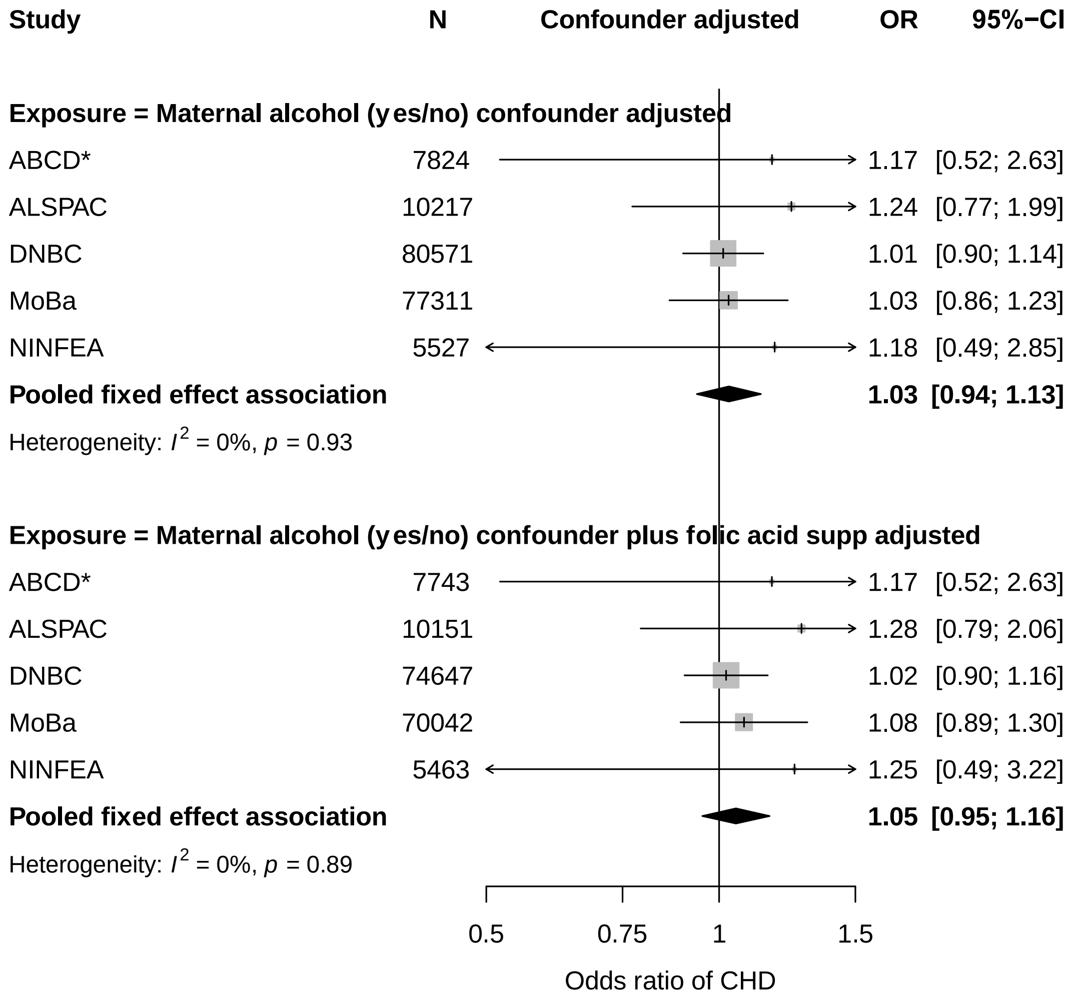

**Figure S22.** Associations between maternal drinking during the first trimester and offspring congenital heart disease. Results are adjusted for all confounders (top) and all confounders plus additional adjustment for folic acid supplementation during weeks 0-12 of pregnancy (bottom). Results are for first trimester drinking or any drinking during pregnancy where trimester data were not available (denoted by *).

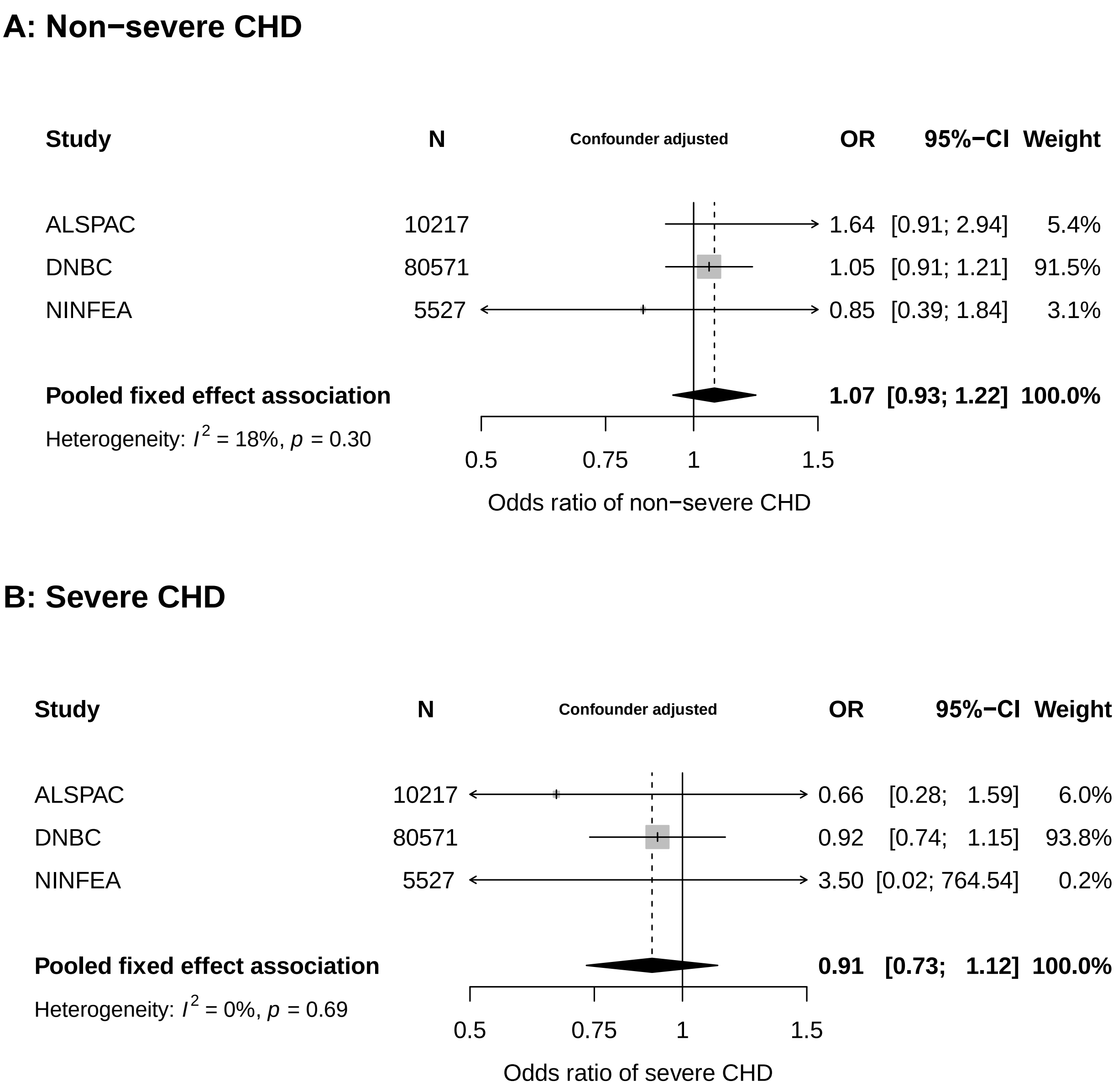

**Figure S23.** Fully adjusted associations between maternal alcohol consumption during the first trimester and offspring non-severe congenital heart disease **(A)** and severe congenital heart disease **(B).** Definitions for CHD subtypes can be found in Table S2

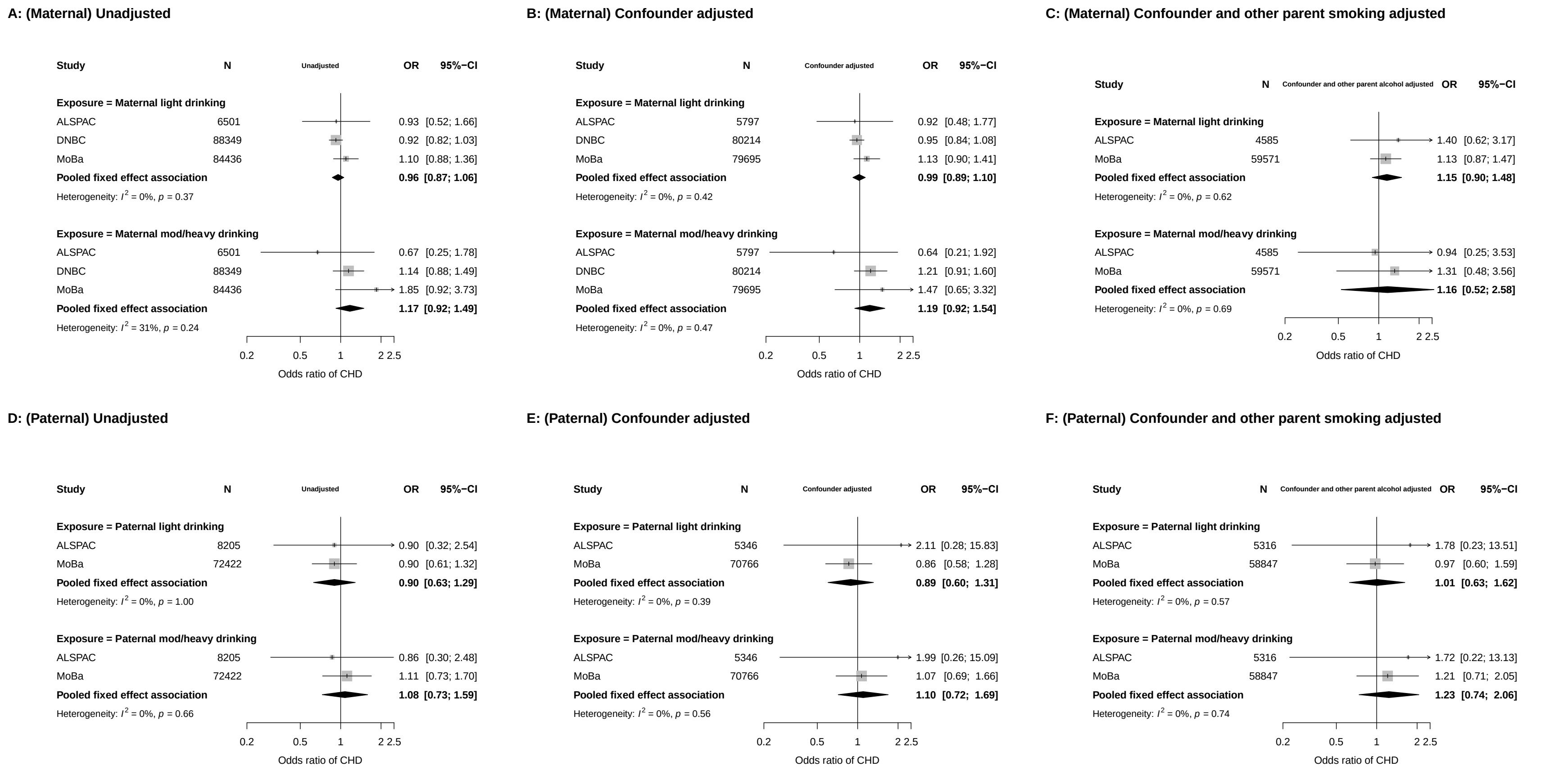

**Figure S24.** Associations **(top (A, B & C): maternal, bottom (D, E & F): paternal)** between parental alcohol intake and offspring congenital heart disease. Results are unadjusted (left), adjusted for all confounders (middle) as well as all confounders and other parents smoking (right). Maternal alcohol intake categorized as none (non-drinker), light (< 3 units per week during pregnancy) and moderate/heavy (≥ 3 units per week during pregnancy). Paternal alcohol intake categorized as none (non-drinker), light (< 7 units per week during pregnancy) and moderate/heavy (≥ 7 units per week during pregnancy). Results presented as odds ratios and 95% confidence intervals for offspring CHD in comparison to non-drinkers.

**Table S10.** Meta-analysis results for associations between alcohol intake and CHDs. Results reported as odds ratios and 95% confidence intervals for CHD/chromosomal defects in comparison to non-drinkers.

| *Model* | *Main analysis*<br>*Outcome = CHD* | *Additional analysis*<br>*Outcome = CHD with chromo/gen removed from study population* |
|---|---|---|
| ***Confounder adjusted*** | **M** – y/n: 1.03 (0.93, 1.13) | **M** – y/n: 1.04 (0.94, 1.15) |
| | **M** – light: 0.99 (0.89, 1.10)<br>**P** – light: 0.89 (0.60, 1.31) | **M** – light: 0.95 (0.85, 1.07)<br>**P** – light: 1.13 (0.69, 1.87) |
| | **M** – mod/heavy: 1.19 (0.92, 1.54)<br>**P** – mod/heavy: 1.10 (0.72, 1.69) | **M** – mod/heavy: 1.24 (0.94, 1.63)<br>**P** – mod/heavy: 1.36 (0.79, 2.34) |
| ***Confounder and other parent BMI adjusted*** | **M** – light: 1.15 (0.90, 1.48)<br>**P** – light: 1.01 (0.63, 1.62) | **M** – 1.17 (0.88, 1.55)<br>**P** – light: 1.21 (0.68, 2.16) |
| | **M** – mod/heavy: 1.16 (0.52, 2.58)<br>**P** – mod/heavy: 1.23 (0.74, 2.06) | **M** – mod/heavy: 1.20 (0.52, 3.17)<br>**P** – mod/heavy: 1.52 (0.82, 2.80) |

**M** = maternal
**P** = paternal
y/n = alcohol as a binary variable, yes or no.
Estimates from yes/no analyses derived from 5 cohorts (ABCD, ALSPAC, DNBC, MoBa, NINFEA).
Estimates from maternal light and mod/heavy drinking analyses derived from ALSPAC, DNBC and MoBa in fully adjusted results, but only ALSPAC and MoBa in paternal and mutually adjusted results.

#### Text S6. Acknowledgements

We are grateful to all participating hospitals, obstetric clinics, and general practitioners for their assistance in implementing the ABCD study and thank all of the women who participated for their cooperation.

We are grateful to all the families who took part in the ALSPAC study, the midwives for their help in recruiting them, and the whole ALSPAC team, which includes interviewers, computer and laboratory technicians, clerical workers, research scientists, volunteers, managers, receptionists and nurses.

The authors thank the families for their continued support and the Cork BASELINE Birth Cohort Study research team.

BiB is only possible because of the enthusiasm and commitment of the children and parents. We are grateful to all the participants, practitioners, and researchers who have made BiB happen.

The Danish National Birth Cohort was established with a significant grant from the Danish National Research Foundation. Additional support was obtained from the Danish Regional Committees, the Pharmacy Foundation, the Egmont Foundation, the March of Dimes Birth Defects Foundation, the Health Foundation and other minor grants. The DNBC Biobank has been supported by the Novo Nordisk Foundation and the Lundbeck Foundation.

We are grateful to all the participating families in Norway who take part in the ongoing Norwegian Mother, Father and Child cohort study.

The authors thank all families participating in the NINFEA cohort.
